# Effects of exogenous oxytocin on human brain function are regulated by oxytocin gene expression: a meta-analysis of 20 years of oxytocin neuroimaging and transcriptomic analyses

**DOI:** 10.1101/2025.08.24.25334303

**Authors:** Junjie Wang, Xianyang Gan, Mengfan Han, Wenyi Dong, Jingxian He, Kun Fu, Mercy C. Bore, Ting Xu, Benjamin Klugah-Brown, Stefania Ferraro, Benjamin Becker

## Abstract

Over the past two decades, numerous pharmaco-imaging studies have explored the neural basis of oxytocin (OT) on human cognition and behavior, yet findings are heterogenous and the links between the macroscopic neurofunctional effects and microscopic organization of the brain remain unclear. Here, we systematically explored the general neuromodulatory effects of OT across behavioral domains and populations by combining neuroimaging meta-analysis, meta-analytic connectivity modeling, behavioral decoding, and transcriptomic analysis. The primary meta-analysis based on data from 75 experiments and n = 2,247 participants revealed consistent domain-general neural effects of OT on the left thalamus, pallidum, caudate, and insula. Meta-analytic connectivity analyses revealed that these regions constituted a functionally integrated thalamus-striatum-insula circuit, directly influenced by OT, embedded within a larger secondary network, and associated with reward, motivation, and emotion, potentially mediating the complex effects of OT. Transcriptomic analyses revealed that the expression of three core OT pathway genes (CD38, OXT, and OXTR) is enriched in these subcortical regions and associated with the observed neural effects. Moreover, OT’s effects were linked with the distribution of acetylcholinergic, dopaminergic, opioidergic, and vasopressinergic gene expressions, potentially reflecting functionally relevant interactions with these systems. Together, these findings delineate a robust effect of OT on human brain function that can help to inform effective pharmacotherapeutic targets, and highlight a core circuitry via which OT exerts its complex regulatory role.

**Highlights:** Meta-analytic evidence for robust effects of exogenous OT on brain activity

A thalamus-striatum-insula circuitry showed domain-general OT effects

OT effects mediated by OT pathway gene expression (OXT, OXTR, CD38)

OT effects interact with multiple neurotransmitter systems

Decoding links OT targets to reward, emotion, social behavior

## 1. Introduction

Oxytocin (OT) is a highly conserved hypothalamic neuropeptide (Gimpl & Fahrenholz, 2001; Jurek & Neumann, 2018; Menon & Neumann, 2023; Meyer-Lindenberg et al., 2011). While initially known for its reproductive function, such as uterine contraction and lactation (Hermesch et al., 2024), accumulating evidence has indicated an essential role of OT in regulating social and affective domains (Kendrick et al., 2018; Quintana et al., 2021). The synthesis of OT and the development of OT-containing nasal sprays (Gronroos et al., 1962; Luhman, 1963; Vigneaud et al., 1953) together with its combination with experimental paradigms and functional neuroimaging facilitated the exploration of the behavioral and neural effects of OT in humans (Kirsch et al., 2005; Kosfeld et al., 2005; Meyer-Lindenberg et al., 2011; Tully et al., 2023). Several studies reported that exogenous OT regulates reward, affective and social domains and associated neural systems in humans (Baumgartner et al., 2008; Chen et al., 2020b; Hu et al., 2015; Lan et al., 2023; Ma et al., 2018; Xu et al., 2019). These findings have motivated translational studies aiming to explore the therapeutic potential of OT in disorders characterized by dysfunctions in these domains (Feifel et al., 2010; Giovanna et al., 2020; Le et al., 2022; Mellentin et al., 2023; Zhao et al., 2024). However, the rapidly expanding literature examining the effects of OT on human brain function simultaneously revealed considerable heterogeneity in both location and direction of the reported effects. Furthermore, these neurofunctional effects have been shown to depend on both, personal and contextual factors, e.g., sex or the social versus nonsocial contexts (e.g., Bartz et al., 2011; Fu et al., 2025; Ma et al., 2018; Yao et al., 2018b).

While the neural and behavioral effects of OT show considerable variability (Spetter et al., 2018), it is neurobiologically plausible that a set of core systems mediates the effects across task domains and individual differences, thereby gating its influence on overarching behavioral domains. Converging evidence suggests the presence of a distributed, core functional network in the human brain that is consistently recruited across task domains and enables flexible switching of both low- and high-order cognitive functions in humans by dynamically guiding and configuring neural processing (Cole et al., 2014; Krienen et al., 2014; Shine et al., 2019; Tononi & Edelman, 1998). Supporting this notion, a recent pharmacological-imaging meta-analysis demonstrated such a key functional architecture and its neurotransmitter modulation via delta-9-tetrahydrocannabinol, which exerts neuromodulatory effects in brain regions central to numerous cognitive and affective processes (Gunasekera et al., 2022). In line with this framework, it is conceivable that exogenous OT exerts its complex behavioral effects via two complimentary mechanisms: (1) direct modulation of core brain systems that are biologically primed for OT sensitivity due to their microscopic architecture, particularly a higher expression of OT sensitive receptors and related pathway components (Gimpl & Fahrenholz, 2001; Quintana et al., 2019), and (2) indirect modulation of communication between these core systems and a broader set of brain systems, thereby enabling complex regulatory effects that operate both in interaction with and across different task contexts (e.g., Bethlehem et al., 2013; Jiang et al., 2021; Wu et al., 2022; Xiao et al., 2024; Zhao et al., 2019).

To address these questions, previous studies have begun to quantitatively summarize the convergence of the modulatory effects of OT across a variety of tasks, within specific task types, and within specific populations, by means of coordinate-based neuroimaging meta-analysis (Grace et al., 2018; Rocchetti et al., 2014; Wang et al., 2017; Wigton et al., 2015). These studies capitalized on the increasing number of pharmacological functional magnetic resonance imaging (fMRI) studies administering OT and the development of neuroimaging meta-analyses allowing a robust quantification of congruent neural effects across study populations and paradigms (Eickhoff et al., 2012; Klugah-Brown et al., 2021; McTeague et al., 2020). Two early meta-analyses reported that OT enhanced activity of the left anterior insula across fMRI studies targeting emotional processes (Rocchetti et al., 2014; Wigton et al., 2015). Subsequently, an updated meta-analysis further revealed general OT effect across all studies, i.e., OT enhanced activation in the bilateral amygdala and left caudate, and attenuated activation in bilateral amygdala (extending to globus pallidus) simultaneously (Wang et al., 2017). In consideration of the potential bias introduced by the incorporation of non-whole-brain results in Wang’s (2017) study, a subsequent meta-analysis followed up the results and reported no overall consistent OT effect across behavioral domains, but an OT enhancing effect in the superior temporal gyrus during emotional processes (Grace et al., 2018). Since the last meta-analyses, the pool of suitable studies has nearly doubled and the methodological quality of both, the original OT studies (Quintana et al., 2021) and the meta-analytic procedures (Eickhoff et al., 2017) has increased as well. We therefore capitalized on the expanded database and recent methodological advances in the field to facilitate identification of robust and unbiased neural effects of OT, which is vital for establishing overarching biological frameworks and advancing therapeutic applications.

Furthermore, methodological progress has allowed to explore how microscopic properties of the brain influence neuroimaging phenotypes (Long et al., 2024), including behavioral performance (Liu et al., 2024), mental disorders (Li et al., 2024; Luo et al., 2023), and effects of psychopharmacological agents (Fan et al., 2025; Gunasekera et al., 2022; Xu et al., 2024). As a macroscopic phenotype, the neural effects of OT administration may also be shaped by microscopic biological processes (Chen et al., 2020a), in particular the biological architecture of the OT system. Three genes in the OT pathway, oxytocin/neurophysin I prepropeptide (OXT), oxytocin receptor (OXTR), and CD38 molecule (CD38) play pivotal roles in the synthesis, storage, release, and binding of OT (Feldman et al., 2016; Gimpl & Fahrenholz, 2001; Jin et al., 2007; Jurek & Neumann, 2018) and consequently may mediate the neural effects of exogenous OT. Initial evidence suggested that CD38 and OXTR are more densely expressed in subcortical and olfactory regions, respectively (Quintana et al., 2019), relative to the average brain-wide expression level, with the former being robustly activated in prior pharmacological-imaging studies of OT administration (Grace et al., 2018), and the latter being recognized as a key node in the pathway of OT administration into the brain (Quintana et al., 2016). A subsequent study correlated OT-related neuroimaging phenotypes with gene expression and found higher OXTR expression in subcortical regions sensitive to OT administration (Habets et al., 2021). However, the first study only offered indirect evidence, while the second used Allen Human Brain Atlas (AHBA) anatomical structures differing from standard neuroimaging segmentations, making precise alignment and functional annotation challenging (Arnatkevic_iūtė et al., 2019). Additionally, the high co-expression of OT pathway genes with dopaminergic and acetylcholinergic genes (Quintana et al., 2019) and accumulating recent evidence indicating that the neurobehavioral effects of OT are critically mediated by monoamine neurotransmitters (e.g., Jansen et al., 2023; Lan et al., 2022; Liu et al., 2021) suggest that OT may exert its regulatory role via various neurotransmitter systems (Dal Monte et al., 2017; Landgraf & Neumann, 2004; Sauer et al., 2013; Zhang et al., 2024b).

Finally, accumulating evidence has indicated that OT’s effects on a core set of localized brain areas lead to changes in associated brain functional networks, in both, mesoscopic animal models (Froemke & Young, 2021; Valtcheva et al., 2023) and macroscopic human pharmacological imaging approaches (Seeley et al., 2018; Xin et al., 2018; Yao et al., 2018a; Zhao et al., 2016). While the original studies allow to explore context and population-specific OT networks, recent progress in meta-analytic modelling of connectivity and co-activation patterns allows to determine highly generalizable network connectivity patterns of brain systems across task paradigms and populations (Langner & Camilleri, 2021). The investigation of such a functional network characterization may allow to identify unique, task-unspecific, generalized functional networks that are directly or indirectly modulated by OT and mediate the complex effects of OT.

Against this background, we aimed to re-examine robust neurofunctional effects of OT administration on brain function in humans by a series of pre-registered updated and comprehensive neuroimaging meta-analyses employing a considerable larger database of original studies and a more robust meta-analysis methodology (Eickhoff et al., 2017), neuroimaging transcriptomics allowing to link macroscopic imaging phenotypes with the microscopic architecture of the OT system, as well as meta-analytic connectivity and co-activation modelling (Langner & Camilleri, 2021).

Primary aims were as follow. (1) Determining general neural effects of OT across tasks and populations (by pooled meta-analyses) and examining the effects in both intensity (disregarding the direction of OT effect) and direction frameworks (enhancing and attenuating effects of OT; Müller et al., 2017). Based on previous studies, we hypothesized convergent effects in the basal ganglia (Hikosaka et al., 2014; Nelson & Kreitzer, 2014; Utter & Basso, 2008), thalamus (Rikhye et al., 2018; Sherman, 2016; Shine et al., 2023), and insula (Gasquoine, 2014; Namkung et al., 2017). The main meta-analytic results were followed up by subgroup meta-analyses examining OT effects separately for the domains of social, emotional, and reward processing, separately for female and male, and separately for healthy populations and populations with brain-related diseases/disorders.

Next, (2) we aimed to link the core regions showing OT effects on the macroscopic level with the underlying microscopic organization of the OT signaling system by means of transcriptome-neuroimaging analyses (Arnatkevic_iūtė et al., 2019). We first determined whether the three OT pathway genes (OXT, OXTR, CD38) were densely expressed in brain regions that converged significantly in the updated meta-analysis and whether the spatial distribution of their expression levels in the human brain was linearly associated with the neural effects of OT. Based on an increasing number of studies reporting that OT exerts its effects in interaction with other neurotransmitter systems, we next explored associations with gene expression profiles of dopaminergic, acetylcholinergic, opioidergic, and vasopressinergic systems. We hypothesized that OT pathway genes were densely expressed in subcortical regions, and that the spatial distribution of the neural effects of OT was significantly correlated with OT pathway gene expressions and co-predicted by expressions of OT pathway genes as well as the other neurotransmitter-associated genes (Quintana et al., 2019; Rokicki et al., 2022).

Then, (3) we employed meta-analytic connectivity modeling (MACM; Laird et al., 2013), to identify regions that interact with the core regions of OT effects as identified by the primary meta-analyses. We proposed that a core network directly affected by OT would exist and exhibit co-activation with larger scale brain networks to facilitate the complex effects of OT brain-wide activation pattern centered on the core network.

Finally, two exploratory analyses were further performed to validate and strengthen findings above. Recent advances in behavioral decoding techniques enable us to directly link OT-induced neural changes to behavior, and we therefore performed an exploratory analysis to determine whether OT effect is associated with behavioral dimensions such as reward, emotion, or social salience. Also, spatial correlations between brain effects of OT revealed by meta-analysis and in-vivo molecular imaging phenotypes of candidate neurotransmitters were computed to support potential transcriptomic-neuroimaging associations.

## 2. Method

### 2.1. Literature search

The present meta-analysis was preregistered on the Open Science Framework (https://osf.io/g9xsb) and adhered to Preferred Reporting Items for Systematic reviews and Meta-Analyses (PRISMA) statement (Page et al., 2021) and recommendations for neuroimaging meta-analyses (M ü ller et al., 2018). The study focused on whole-brain activation results from fMRI studies administering exogenous OT published between December 2005, the publication date of the first fMRI study administering intranasal OT (Kirsch et al., 2005), and January 2024. The literature was retrieved with the following search terms: “BOLD” OR “blood oxygen level-dependent” OR “neuroimaging” OR “neural imaging” OR “brain imaging” OR “pharmaco-imaging” OR “functional imaging” OR “MRI” OR “fMRI” OR “magnetic resonance imaging” AND “oxytocin” OR “syntocinon” OR “pitocin”, on the following five databases: Web of Science, Scopus, Ebsco, PubMed, and Science Direct. The specific search criteria and codes were available in Supplementary Material I. Citing and cited articles of the four previous meta-analyses on OT were searched for further relevant articles. Google Scholar was additionally screened. As shown in the PRISMA Flowchart (Fig. 1) a total of 6,904 records were retrieved, of which the full text of 183 studies was downloaded and assessed for eligibility.

**Fig. 1.**
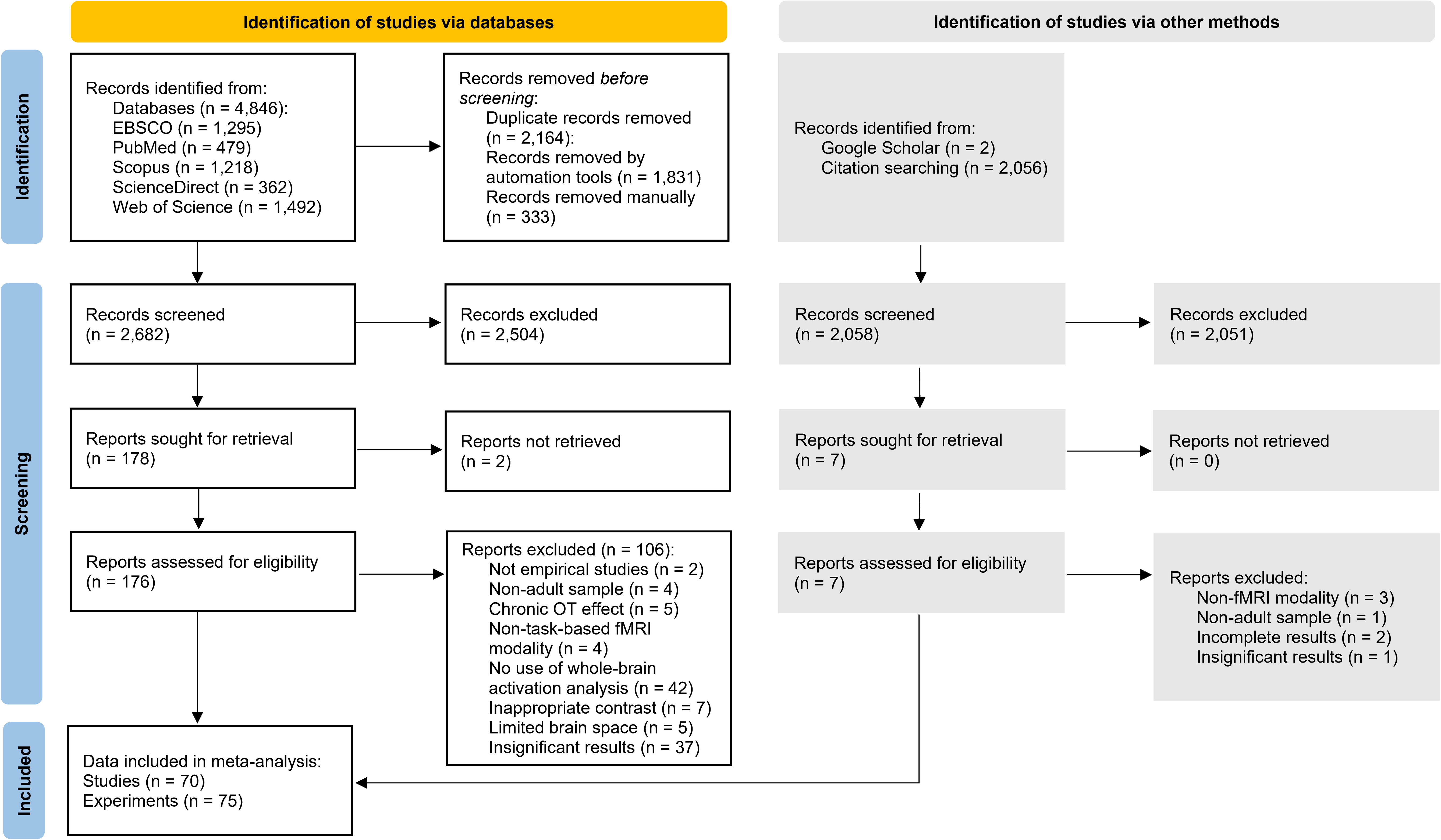
Flowchart illustrating the literature retrieval, exclusion and inclusion process according to PRISMA statement.

### 2.2. Eligibility criteria

Inclusion criteria for the studies encompassed (1) sample size > 5 participants, (2) originally published in a peer-reviewed journal in English, (3) administering exogenous single-dose OT (excluding multi-dose OT studies), (4) employing task-based fMRI rather than resting-state fMRI or other neuroimaging techniques, (5) reporting significant whole-brain activation results for main or interaction effect based on interpretable placebo-controlled contrasts, (e.g., “Oxytocin > Placebo” and “Placebo > Oxytocin”, excluding e.g., “Oxytocin vs Vasopressin”), (6) and reporting peak foci of activation in either Talairach (TAL) or Montreal Neurological Institute (MNI) space. Of note, we contacted corresponding authors of two studies reporting significant whole-brain results without specific brain coordinates to reduce the possibility of a biased dataset (Chen et al., 2016; Liu et al., 2019a), and corresponding coordinates provided within one month were included in the final analysis

(from Chen et al., 2016).

### 2.3. Data selection and extraction

The following data were extracted for each study. (1) Index information of the study: authors, article title, name of publication, year of publication. (2) Sample characteristics: sample size, disorder status, gender, and age. (3) Administration information: dosage, administration route, administration design, and time interval between administration and task-based fMRI scanning. (4) Task characteristics: name of the task, type of main stimulus used in the task, main sensory modality, behavioral domain of the task, task contrasts included, and whether multiple contrasts were included. (5) Coordinate information: position (x, y, z) and intensity value (T/F/Z/P) of each coordinate, number of foci and intensity of foci (mean, maxima, and minima), coordinate space, source of the coordinates, and whether the coordinates were corrected for multiple comparisons. Additionally, for nine studies with key information unclear (e.g., number of subjects, or whether the coordinates were based on whole-brain analysis), we further contacted the corresponding authors of these publications to complete accurate records of the information and data analyses (Baumgartner et al., 2008; Cavalli et al., 2017; Eckstein et al., 2016; Eckstein et al., 2014; Hu et al., 2015; Ide et al., 2018; Kanat et al., 2015; Pincus et al., 2010; Tully et al., 2023). To warrant unbiased results, not only the full text assessment was independently carried out by two reviewers (J. W., W. D.), but also the extraction of coordinates was independently performed by two reviewers (J. W., J. H.). Any disagreement between the two reviewers was resolved by discussion and a final decision by a third independent reviewer (B. B.). Other information was recorded by J. W. and validated by another independent reviewer (M. H.).

### 2.4. Activation likelihood estimation

Coordinate-based meta-analyses were implemented in GingerALE 3.0.2 (http://www.brainmap.org/ale/) to determine consistency in reported brain regions showing differences in activation between the oxytocin group/condition vs the placebo group/condition (Eickhoff et al., 2012). Prior to the formal analysis, the coordinates in TAL space were transformed to MNI space using the Lancaster transform algorithm embedded in GingerALE (Laird et al., 2010), and the coordinates from different contrasts from the same dataset were combined into one single experiment to minimize within-group effects (Turkeltaub et al., 2012). The ALE algorithm quantitatively evaluated spatial convergence of reported neural activations beyond chance-level clustering within gray matter (Eickhoff et al., 2009), and generally followed three key stages. (1) Activation map generation: the activation probabilities were created for each study by assigning value “1” to peak coordinates, lower values determined by an isotropic Gaussian kernel with width inversely dependent on each sample size to neighboring voxels, and value “0” to non-activated voxels, which were further used for generating the modeled activation maps (MA) for each study via combining the probabilities in all activation foci for each voxel (Eickhoff et al., 2012); (2) Integration of activation probabilities: the MA maps for each experiment were combined on a voxel level to obtain an ALE map, that is, the activation likelihood for each voxel across experiments; (3) Statistical inference: an uncorrected voxel-level threshold of p < 0.001 was adopted in identifying significant convergence of voxels across experiments, then ALE identified significant and spatially contiguous voxels based on the p-value of each voxel to organize clusters of voxels, from which significantly activated brain regions were further obtained by a cluster-level family-wise error (FWE) correction at a threshold of p < 0.05 and a null distribution from 5,000 permutation tests (Eickhoff et al., 2016; Gan et al., 2022). This approach has demonstrated a good balance between Type I error control and power (Eickhoff et al., 2016). Finally, the anatomical label of each peak coordinate in each cluster was extracted from the Anatomic Automatic Labeling atlas (AAL3; Rolls et al., 2020). For the coordinate that could not be successfully identified to an anatomical label according to AAL3, the label identified in the Harvard-Oxford atlas or the original label in the ALE was used as an alternative.

### 2.5. General and sub-group meta-analyses

In line with the main aims of the present project, the impact of OT on brain function was (1) assessed across all behavioral domains to determine brain systems generally modulated by OT, including the determination of regions that showed a robust increase, decrease or non-directional effect of OT; (2) assessed separately for the domains of social, emotional, and reward processing, and (3) assessed separately for female, male, healthy populations and populations with brain-related diseases.

In line with prior work (Grace et al., 2018; Wang et al., 2017), we initially explored the enhancing effect (OT > PL) and attenuating effect (OT < PL) of OT separately to improve the interpretation of the results. In contrast to previous studies, we additionally merged the coordinates of enhancing and attenuating effects of OT to detect regions generally involved in the effects of OT irrespective of direction (OT > PL & OT < PL; similar approach see also Feng et al., 2021; Müller et al., 2017). This approach additionally allowed to increase power by capitalizing on the entire set of available studies, and fully considered individual and context specific effects of OT, e.g., the direction of OT effect on the same brain region varied depending on gender and task valence (Domes et al., 2010; Lan et al., 2024). In addition, given the domain-specific effects of OT in social, emotional, and reward processing, the task contrasts in the above studies were categorized into social processing subgroup, emotional processing subgroup, positive valence system subgroup, and negative valence system subgroup, respectively. All emotional task contrasts that were involved in the recognition or perception of emotions or that could explicitly elicit any sort of emotional experience, as well as other task contrasts capable of bringing about emotional consequences implicitly, were incorporated into the emotional processing subgroup (Ma, 2015). The classification of the social processing subgroup and the two valence subgroups was implemented according to the Research Domain Criteria (RDoC) Projects (https://www.nimh.nih.gov/research/research-funded-by-nimh/rdoc/constructs/rdoc-matrix) released by the National Institute of Mental Health (Insel et al., 2010).

Also, in an effort to investigate the population-specific neural effects of OT administration, the studies were further categorized into separate male, female, healthy population, and patient groups based on gender and whether or not the disease was present reported in each study. The patient subgroup consisted primarily of both clinical populations with Asperger’s Disorder (n = 1), Autism Spectrum Disorder (n = 4), Depression Disorder (n = 1), Frontotemporal Dementia (n = 1), Generalized Social Anxiety Disorder (n = 1), and Schizophrenia Spectrum Disorder (n = 2), and sub-clinical populations with Social Drinking (n = 2) as well as Clinical High Risk for Psychosis (n = 1). The categorization of all subgroups was conducted independently by two researchers (J. W., K. F.), and any discrepancy that arose between the two was discussed with, and ultimately resolved by, a third researcher (B. B.). The described above meta-analyses for the non-directional effect, enhancing effect, and attenuating effect were replicated in each subgroup. Of note, in spite of insufficient power to detect smaller effects and results may be driven by a single experiment in ALE meta-analysis including fewer than 17 experiments (Eickhoff et al., 2016), we still exploratively conducted meta-analysis in subgroups with fewer than 17 but greater than 10 experiments. However, findings from inadequate sample size should be construed with caution and remain to be further verified.

### 2.6. Transcriptomic-neuroimaging analysis

The gene expression data were downloaded from the official website of AHBA (https://www.brain-map.org), and then preprocessed with reference to the standard processing pipeline and processing flow in the pioneering transcriptomic study of OT pathway genes (Arnatkevic_iūtė et al., 2019; Hawrylycz et al., 2012; Quintana et al., 2019). The main procedure included probe-to-gene re-annotation, gene filter and selection, probe selection, and mapping samples to brain regions (see details in “Method” section in Supplementary Material I).

After preprocessing of the gene expression data, we performed a series of stepwise analyses to explain the microscopic basis of the macro-neurological effects of OT. First, we identified the two clusters activated in the general and non-directional effect of OT as ROIs, and then further extracted the average expression values in the two regions for all candidate genes we included in each donor. One sample t-tests across donors were then performed to assess which genes in these two regions exhibited higher expression values compared to the average brain-wide expression values. The differences between the average expression values of each brain region in the AAL atlas and the whole brain have been well explored (Quintana et al., 2019), but we still measured which brain regions parceled on the Brainnetome atlas showed significantly higher or lower gene expression than the whole-brain average expression, as different atlases would yield distinct results and atlas-based results could be useful validation of the above mentioned ROI-based findings. Secondly, Pearson spatial correlations between the unthresholded z-map representing the general and non-directional effect of OT and the expression maps of candidate genes were calculated, whose significance was corrected by FDR with a threshold of 0.05. Third, the partial least squares (PLS) regression analysis was used to examine multivariate associations between the general and non-directional OT effect observed in meta-analysis and gene expressions (Geladi & Kowalski, 1986), with the gene expression matrix (20 genes × 123 regions) as the predictor variable and OT effect brain matrix (1 × 123 regions) as the response variable. A list of PLS components was obtained by iterative computation, with ordering based on the maximized capacity to explain the covariance matrix of the independent and dependent variables. Typically, the first component in the PLS (PLS 1) collectively reflected the most significant covariance characteristics between gene expression patterns and neural effects (Abdi & Williams, 2013). To verify whether PLS 1 captured the multivariate relationship between gene expressions and OT effect, we also computed the Pearson’s correlation between the gene expression weights in PLS 1 and the brain matrix for OT effect (Xie et al., 2022; Xu et al., 2024). Finally, 10,000 instances of bootstrapping for PLS weight of each gene were calculated to build a sampling distribution representing the overall sample. We transformed the PLS weight of each gene into a z-scored value via dividing the weight by the standard deviation of bootstrapped sample set, and sorted all the z-scored weights to identify the genes with highest positive and negative weights.

### 2.7. MACM analysis

We employed MACM to determine which larger scale brain systems the regions showing robust OT effects communicate with. MACM was a method used for performing second-order meta-analyses on studies reporting activation in region of interest (ROI) retrieved from large neuroimaging databases (Robinson et al., 2012), to test the co-activation patterns associated with brain regions revealed in neuroimaging meta-analyses in a data-driven and highly generalizable fashion (Laird et al., 2013; Langner & Camilleri, 2021). MACM-based FC was generally developed as a post hoc extension of neuroimaging meta-analysis results (Fascher et al., 2024; Gan et al., 2022; Reimann et al., 2023), as it provides a network-level functional characterization by identifying which other brain regions the meta-analytically identified regions were functionally interconnected with, and by testing how these networks align with higher-order large-scale functional networks. Accumulating evidence has further demonstrated its reliability and potential as an alternative to resting-state FC and anatomical connectivity through the demonstration of its similarity to them (Cauda et al., 2011; Eickhoff et al., 2010).

Consistent with a previous study (Gan et al., 2022), we identify both OT-constrained and non-OT-constrained FC patterns, as they both might be able to locate OT-regulated core network and sub-network in a mutually complementary way. Therefore, we used two analytical strategies, MACM-A and MACM-B. MACM-A identified which brain regions were connected with the four peak coordinates attributed to the two clusters (left thalamus: -4, -16, 6; left pallidum: -12, -2, -2; left caudate: -10,8,0; left insula: -36, 22, 4) revealed in the meta-analysis of general non-directional effect of OT under the OT effect framework. Remarkably, previous studies have routinely chosen to add further filters in Sleuth (https://www.brainmap.org/sleuth/) to obtain studies under the task topic of interest (Fascher et al., 2024; Gan et al., 2022). However, the insufficient number of studies regarding the OT effect in Sleuth (n = 2) limited us to retrieving data only in the initial meta-analytic database we created to report the OT effect (70 studies, 75 experiments, 900 foci, 151 contrasts, and 2,247 subjects). As successfully implemented in a previous meta-analysis (Maliske et al., 2023), the Euclidean distances between each seed coordinate and all coordinates was calculated, then a study would be included in the MACM-A analysis when it included at least one coordinate whose Euclidean distance from the seed coordinate was less than 20 mm. After meticulous screening, datasets included in the MACM-A analysis for the 4 coordinates were as follows: 17 experiments, 450 foci, and 559 subjects for the left thalamus; 19 experiments, 396 foci, and 632 subjects for the left pallidum; 22 experiments, 469 foci, and 696 subjects for the left caudate; 20 experiments, 476 foci, and 750 subjects for the left insula, whose experiments were sufficient in number for detecting robust convergence (see Supplementary Material VI for detailed information; Eickhoff et al., 2016). For each seed coordinate, a secondary ALE meta-analysis was performed to identify its co-activation patterns under the influence of the OT after organizing the coordinates in all eligible experiments into a set of coordinates similar to those organized in the meta-analysis.

MACM-B fully referred to the standard workflow of MACM analysis (Langner & Camilleri, 2021). The details of MACM-B could be found in “Method” section in Supplementary Material I. The co-activation pattern under the OT-constrained framework (MACM-A) could be defined as a core functional network modulated by OT, whereas that under the non-OT-constrained framework (MACM-B) might be specified as an extended functional network indirectly regulated by OT awaiting further verification. The software MRICroGL 1.2 (https://www.nitrc.org/projects/mricrogl/) was used for visualization of all brain activations with the Colin’s MNI template (https://www.brainmap.org/ale/Colin27_T1_seg_MNI.nii.gz) as the reference.

### 2.8. Exploratory analysis

#### 2.8.1 Behavioral decoding of activation and co-activation patterns

To further explore the behavioral domains of the OT-modulated brain regions/networks, we utilized the Brain Annotation Toolbox (BAT, available at https://istbi.fudan.edu.cn/lnen/info/1173/1788.htm) for functional decoding (Liu et al., 2019b). Based on the Neurosynth database (https://www.neurosynth.org/), the toolbox extracted an initial set of over 3000 terms via text mining technique, then further selected 217 functional terms and their activation maps with unequivocal biological significance and categorized them into 28 broad behavioral domains to serve as a functional annotation dataset (Yarkoni et al., 2011). Function annotation analysis hypothesized that voxel activation patterns within specific brain regions would be significantly co-activated with their functionally relevant meta-analytic terms, and that function-specific cognitive characterizations could be systematically identified via the comparison of activation similarity between the target region and the random voxel set (Liu et al., 2019b). To characterize the effects of OT, we here applied the decoding approach to both, brain regions and co-activation networks determined in the corresponding meta-analyses (see similar approach in Gan et al., 2024). Finally, the p-value for each term was calculated and ranked in ascending order after 10,000 permutations, and the top 20 terms with the highest significance and p < 0.05 were extracted as the functional characterizations most strongly associated with the corresponding brain activation patterns (Gan et al., 2022).

#### 2.8.2 Correlation analysis with vivo molecular imaging phenotype of neurotransmitters

We adopted JuSpace 2.0 (https://github.com/juryxy/JuSpace) to further determine the Pearson’s spatial correlations between unthresholded and z-scored ALE brain map for general and non-directional effect of OT and vivo nuclear imaging-derived maps related with cholinergic, dopaminergic, and opioidergic systems (n = 17), which could provide further validation for findings from genetic analyses (Dukart et al., 2021). In addition to this, exploratory analyses were also performed to determine the links between other vivo nuclear imaging-derived molecular maps incorporated in JuSpace and the neural effects of OT administration. Specifically, we computed the Pearson’s spatial correlations between unthresholded and z-scored ALE brain map for general and non-directional effect of OT and neurotransmitter maps (n = 16) covering the aspartic, cannabinergic, GABAergic, glutamatergic, noradrenergic, and serotonergic neurotransmission, a map for cerebral metabolic rate of glucose (n = 1), as well as some vivo nuclear imaging-derived protein maps (n = 4), which were from previous vivo molecular imaging studies (Positron Emission Tomography, and Single Photon Emission Computed Tomography), collected and assembled by JuSpace (a detailed description and related citations of these vivo nuclear imaging-derived molecular maps can be found in Table S2). The spatial auto-correlation was corrected by calculating the partial correlation with the gray matter probability estimate and a null distribution of spatial correlation coefficients was obtained by 10,000 permutations for computing the p-value (Burt et al., 2020). Given that multiple correlation analyses were performed, the false discovery rate (FDR)-corrected threshold of q < 0.05 was used to correct for false positives from multiple comparisons (Benjamini & Hochberg, 1995).

## 3. Results

### 3.1. Overview of study characteristics

After screening out 113 ineligible studies (Supplementary Material II), 70 studies were ultimately included in the meta-analysis (Table S3; Supplementary Material III). The cohort included 46 studies utilizing exclusively male-only sample, 17 studies employing female-only sample, and 15 studies focusing on (sub-)clinical population. In addition, the sample size ranged from 8 to 196 participants, with mean age spanning 19.68 to 64.29 years old. For the intervention protocols, 58 studies administered a single dose of OT with 24 international units (IU), while few studies administered 25 IU (n = 2), 26 IU (n = 1), 40 IU (n = 5), 72 IU (n = 1), or several dosage regimens (n = 3). Most studies used intranasal administration (n = 68), while two studies utilized oral administration. The time interval between administration and scanning ranged from 10-90 mins (mean = 43.17 mins). Twenty-nine of these studies adopted between-subject design and the other 41 utilized within-subject/cross-over design. Experimental tasks predominantly employed visual stimuli and mainly targeted social, emotional, or reward-processing. The coordinates in 67 studies were reported in native MNI space, and coordinates in three studies were reported in TAL space, which were converted to MNI space before formal analysis. Reported foci per study varied substantially (range: 1-72), with 22 studies reporting uncorrected coordinates and 49 reporting coordinates implementing multiple comparison corrections. The Supplementary Material IV describes detailed information of studies included in each subgroup analysis.

The included studies were organized as follows according to the analytic aims. Non-directional effect: 70 studies, 75 experiments, 900 foci, 151 contrasts, and 2,247 subjects; enhancing effect: 42 studies, 45 experiments, 430 foci, 75 contrasts, and 1,235 subjects; attenuating effect: 29 studies, 31 experiments, 260 foci, 44 contrasts, and 787 subjects. Subgroup analyses replicated above analytical framework and detailed sample characteristics for subgroup analyses were comprehensively documented in Table S4.

### 3.2. ALE meta-analysis for OT effect

The general ALE meta-analysis encompassing all studies with OT administration revealed a non-directional effect of OT in two clusters, located in the left thalamus (extending to left pallidum and caudate) and insula (Table 1, Fig. 2A). However, the separate meta-analyses for the enhancing and attenuating effects of OT failed to reach significant results. The uncorrected results showed a trend: OT enhanced the activation of the left thalamus and right caudate, and decreased the activation of the left putamen and pallidum (Table S5).

**Fig. 2.**
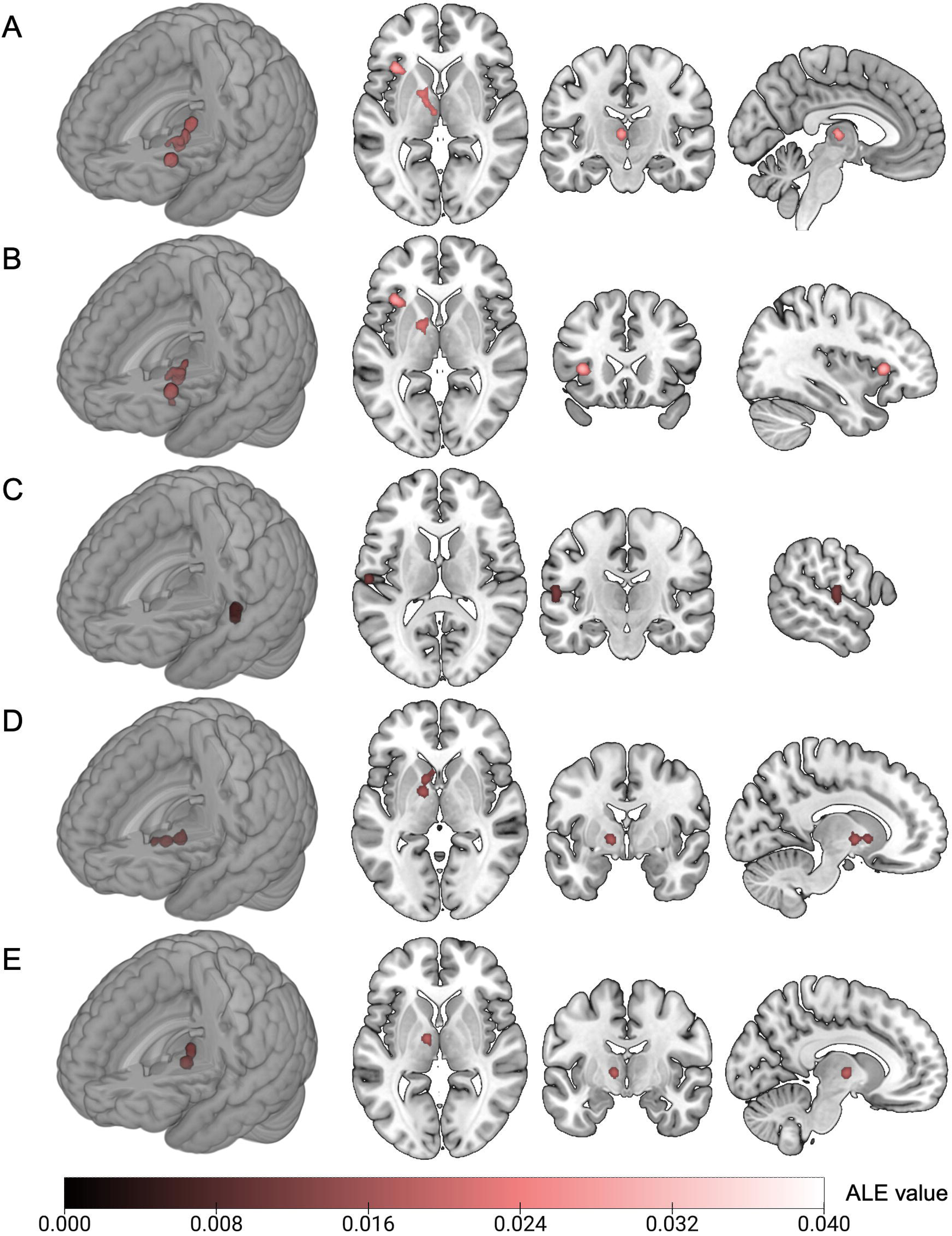
The brain regions affected by OT in the general and subgroup ALE meta-analyses. Panels A, B, and D show the non-directional neural effect of OT obtained from the ALE meta-analyses for full sample, health population subgroup, and positive valence subgroup, respectively. Panels C and E show the enhancing neural effect of OT obtained from the ALE meta-analyses for patient subgroup and social processing subgroup, respectively. The voxel-level threshold is set at p < 0.001, uncorrected; and the cluster-level threshold is set at p < 0.05, FWE-corrected.

**Table 1.**
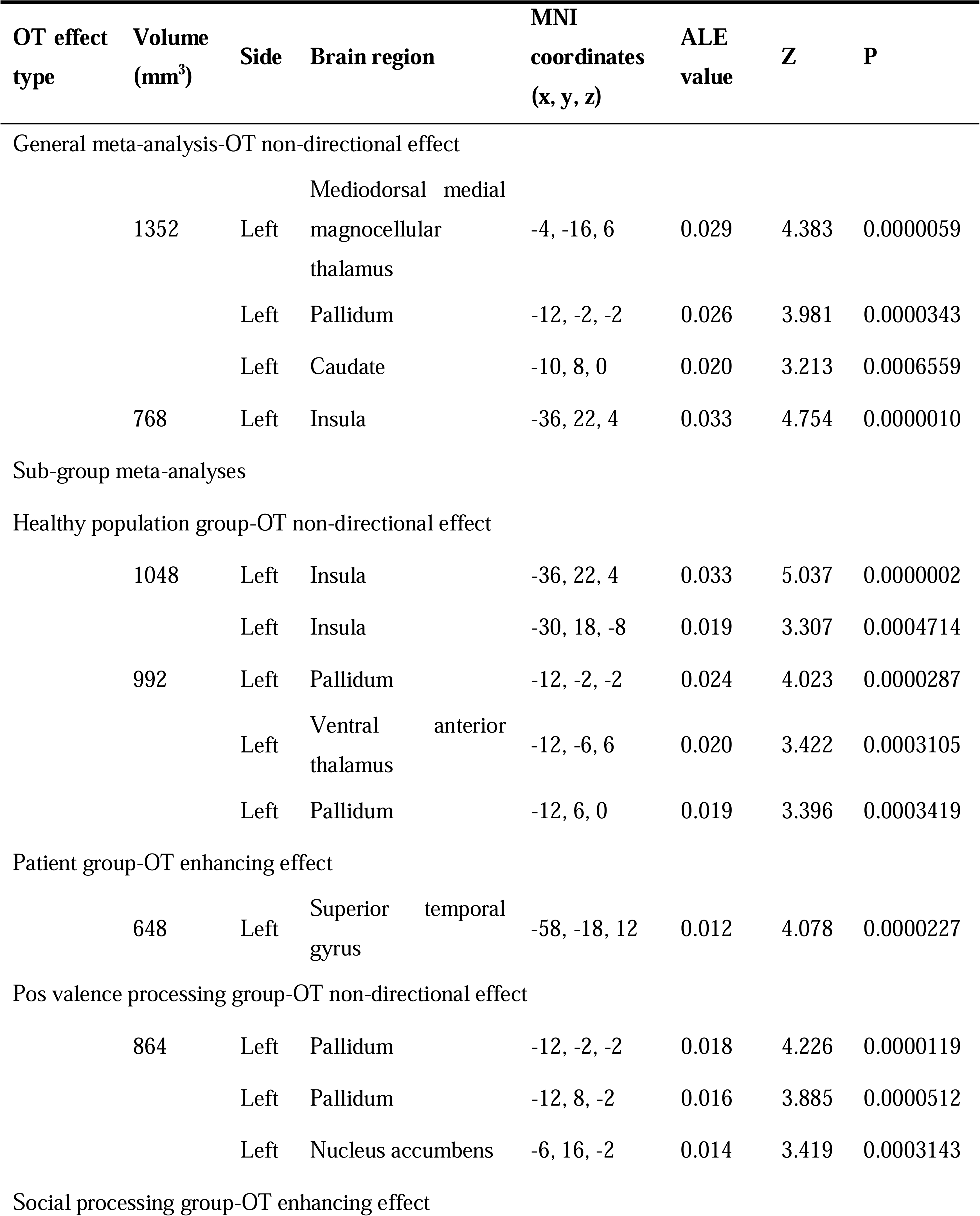
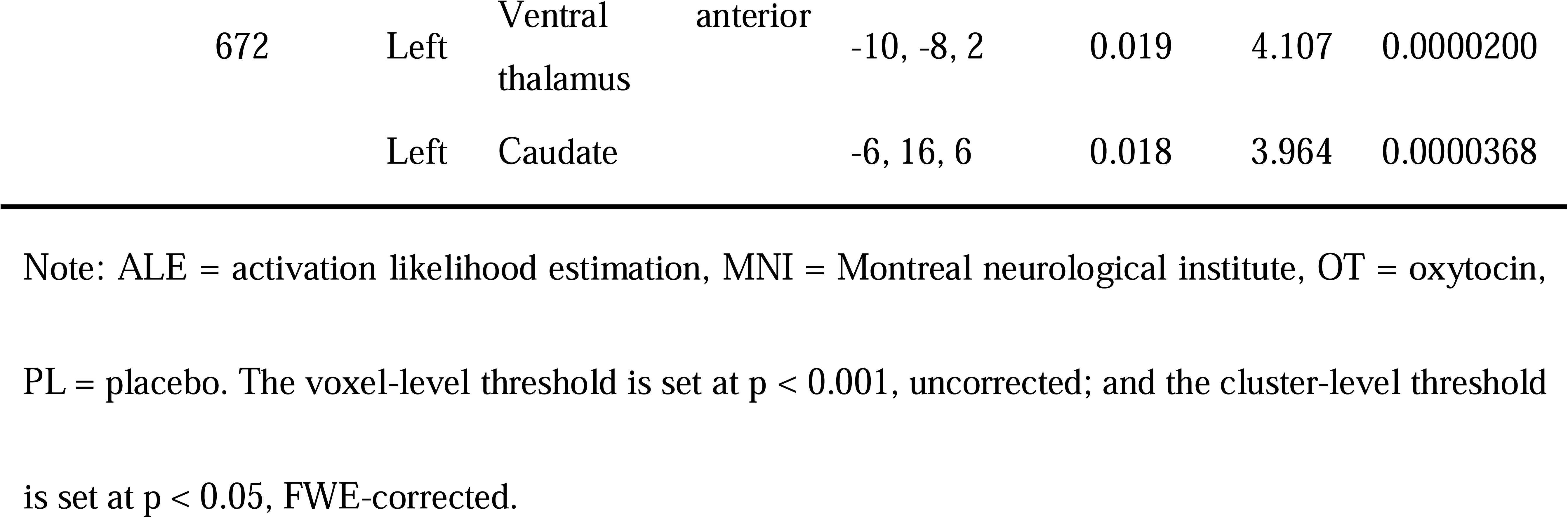
The brain regions affected by OT from the ALE meta-analyses.

The meta-analysis in the healthy population subgroup confirmed a robust non-directional effect of OT in the two clusters spanning the left pallidum (extending to thalamus) and insula (Table 1, Fig. 2B). For the patient subgroup meta-analyses, OT showed an enhancement to the brain activity in the left superior temporal gyrus (Table 1, Fig. 2C).

For the domain-specific analyses, OT exhibited a non-directional effect in the left pallidum (extending to the left nucleus accumbens) during positive valence processing (Table 1, Fig. 2D). OT enhanced the brain activity within the left thalamus (extending to left caudate) during social processing (Table 1, Fig. 2E). No significant clusters of convergence were found in any of other ALE meta-analyses.

### 3.3. Transcriptomic analysis for OT effect

For whole-brain gene expression analyses, CD38 was more intensively expressed within the left thalamus (Tha_8_1, medial pre-frontal thalamus; Tha_8_2, pre-motor thalamus; Tha_8_8, rostral temporal thalamus) and entire basal ganglia (BG_6_1, ventral caudate; BG_6_2, globus pallidus; BG_6_3, nucleus accumbens; BG_6_4, ventromedial putamen; BG_6_5, dorsal caudate; BG_6_6, dorsolateral putamen) compared with the average expression of the whole brain after FDR correction at 0.05 level (Fig. 3A, Supplementary Material V). Likewise, the expressions of OXT and OXTR within the thalamus and basal ganglia were significantly higher than the average expression of the whole brain, whereas such results failed to survive after FDR correction (p_uncorrected_ < 0.05; Fig. 3A, Supplementary Material V). In addition, some acetylcholinergic genes (CHRM1, CHRM2, CHRM3, and CHRM4), dopaminergic genes (DRD1, DRD2, DRD5, and COMT), and opioidergic genes (OPRK1 and OPRM1) showed significantly higher or lower expressions in the thalamus and basal ganglia than that across the whole brain (p_FDR_ < 0.05). Statistical information related to the differences between the expression of each gene in each brain region and corresponding mean expression across the whole-brain can be found in Supplementary Material V. We further measured the mRNA expression of selected OT pathway genes and other neurotransmitter genes in the ROIs, i.e., two clusters (peaking at left thalamus and insula) revealed by meta-analysis for general and non-directional effect of OT. In contrast with the whole brain, the cluster 1 (peaking at the thalamus and extending to basal ganglia) exhibited significantly higher mRNA expressions of all OT pathway genes (CD38 [Hedge’s g = 3.54, p_FDR_ < 0.001], OXT [Hedge’s g = 3.76, p_FDR_ < 0.001], and OXTR [Hedge’s g = 1.70, p_FDR_ = 0.008]), and significant differences in some acetylcholinergic genes (CHRM1 [Hedge’s g = - 4.45, p_FDR_ < 0.001], CHRM2 [Hedge’s g = -2.14, p_FDR_ = 0.003], CHRM3 [Hedge’s g = -3.51, p_FDR_ < 0.001], CHRM4 [Hedge’s g = 4.31, p_FDR_ < 0.001], and CHRM5 [Hedge’s g = 1.97, p_FDR_ = 0.005]), dopaminergic genes (DRD1 [Hedge’s g = -2.50, p_FDR_ = 0.002], DRD2 [Hedge’s g = 6.45, p_FDR_ < 0.001], and COMT [Hedge’s g = 2.44, p_FDR_ = 0.002]), and one opioidergic gene (OPRM1 [Hedge’s g = 1.67, p_FDR_ = 0.008]). And the mRNA expression of one opioidergic gene -OPRK1 in the insula was statistically significantly higher than whole-brain average (Hedge’s g = 2.09, p_FDR_ = 0.035). However, no significant difference was observed for other genes (Fig. 3B, Supplementary Material V).

**Fig. 3.**
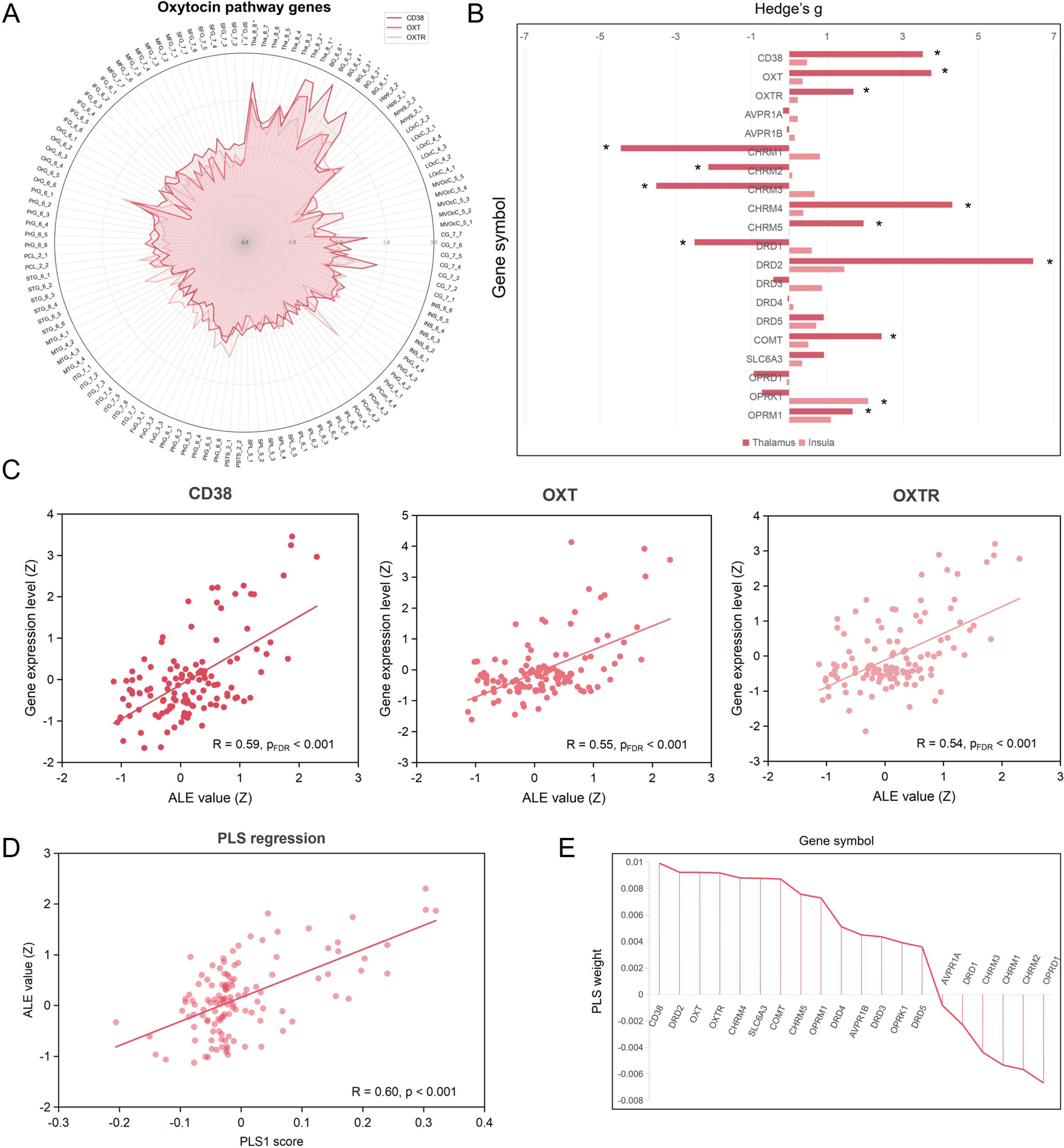
The gene expression profiles and transcriptomic correlates of neural effect of OT. Panel A shows the differences between the expression level of CD38, OXT, and OXTR in the 123 brain regions in the left hemisphere segmented by Brainnetome atlas and average expression level of the whole brain. The Hedge’s g is used for measurement of effect size. A brain region with significance symbol “*” signifies that the difference in gene expression of CD38 in that region compared to the average expression in the whole brain remains significant after FDR correction at 0.05 level. Panel B presents the differences between the expression level of oxytocinergic, vasopressinergic, acetylcholinergic, dopaminergic, and opioidergic genes in the thalamus and insula activated in the ALE meta-analysis for general and non-directional effect of OT and average expression level of the whole brain. “*”, significant difference between the brain region and whole brain after FDR correction at the 0.05 level. Panel C illustrates the Pearson spatial correlations between the ALE map derived from the meta-analysis for general and non-directional effect of OT and gene expression maps of CD38, OXT, and OXTR. Panel D displays the Pearson correlation between the ALE map and PLS scores of the component 1 in the PLS regression. Panel E shows the predictive weight for each gene in the PLS regression. Abbreviations for brain regions: Amyg = amygdala, BG = basal ganglia, CG = cingulate gyrus, FuG = fusiform gyrus, Hipp = hippocampus, IFG = inferior frontal gyrus, INS = insular gyrus, IPL = inferior parietal lobule, ITG = inferior temporal gyrus, LOcC = lateral occipital cortex, MFG = middle frontal gyrus, MTG = middle temporal gyrus, MVOcC = medioventral occipital cortex, OrG = orbital gyrus, PCL = paracentral lobule, Pcun = precuneus, PhG = parahippocampal gyrus, PoG = postcentral gyrus, PrG = precentral gyrus, pSTS = posterior superior temporal sulcus, STG = superior temporal gyrus, SFG = superior frontal gyrus, SPL = superior parietal lobule, Tha = thalamus; Abbreviations for genes: AVPR1A = arginine vasopressin receptor 1A, AVPR1B = arginine vasopressin receptor 1B, CD38 = CD38 molecule, CHRM1 = cholinergic receptor muscarinic 1, CHRM2 = cholinergic receptor muscarinic 2, CHRM3 = cholinergic receptor muscarinic 3, CHRM4 = cholinergic receptor muscarinic 4, CHRM5 = cholinergic receptor muscarinic 5, DRD1 = dopamine receptor D1, DRD2 = dopamine receptor D2, DRD3 = dopamine receptor D3, DRD4 = dopamine receptor D4, DRD5 = dopamine receptor D5, COMT = catechol-O-methyltransferase, SLC6A3 = solute carrier family 6 member 3, OPRD1 = opioid receptor delta 1, OPRK1 = opioid receptor kappa 1, OPRM1 = opioid receptor mu 1, OXT = oxytocin/neurophysin I prepropeptide, OXTR = oxytocin receptor; Other abbreviations: ALE = activation likelihood estimation, FDR = false discovery rate, PLS = partial least square.

In the correlation analyses between OT-related neuroimaging phenotype (Z-scored ALE value map for general and non-directional effect of OT) and gene expression maps, we found significant positive correlations between the spatial distribution of OT effect and that of mRNA expressions of CD38 (R = 0.59, p_FDR_ < 0.001), OXT (R = 0.55, p_FDR_ < 0.001), and OXTR (R = 0.54, p_FDR_ < 0.001; Fig. 3C). Besides, the neural effect of OT was significantly correlated with the mRNA expressions of vasopressinergic gene (AVP1RB [R = 0.27, p_FDR_ = 0.004]; Fig. S1), acetylcholinergic genes (CHRM1 [R = -0.32, p_FDR_ < 0.001], CHRM2 [R = - 0.34, p_FDR_ < 0.001], CHRM3 [R = -0.26, p_FDR_ = 0.004], CHRM4 [R = 0.52, p_FDR_ < 0.001], and CHRM5 [R = 0.45, p_FDR_ < 0.001]; Fig. S2), dopaminergic genes (DRD2 [R = 0.55, p_FDR_ < 0.001], DRD3 [R = 0.26, p_FDR_ = 0.005], DRD4 [R = 0.30, p_FDR_ < 0.001], DRD5 [R = 0.21, p_FDR_ = 0.02], COMT [R = 0.52, p_FDR_ < 0.001], and SLC6A3 [R = 0.52, p_FDR_ < 0.001]; Fig. S3), and opioidergic genes (OPRD1 [R = -0.40, p_FDR_ < 0.001], OPRK1 [R = 0.23, p_FDR_ = 0.011], and OPRM1 [R = 0.43, p_FDR_ < 0.001]; Fig. S4). Subsequently, multivariate neural associations between expression patterns of selected genes of interest and neural effect of OT were identified by PLS regression analysis. The first PLS component (PLS1) explained 36.12% of the variance in the neuromodulatory effect of OT, and PLS1 score was significantly correlated with ALE map of OT effect (R = 0.60, p < 0.001; Fig. 3D). In the prediction of the neural effects of OT, the OT pathway genes made the strongest positive contributions (CD38, OXT, OXTR; Z_mean_ = 8.44), followed by acetylcholinergic genes (CHRM4, CHRM5; Z_mean_ = 6.32), dopaminergic genes (DRD2, DRD3, DRD4, DRD5, COMT, SLC6A3; Z_mean_ = 4.45), opioid genes (OPRK1, OPRM1; Z_mean_ = 2.54), and one vasopressinergic gene (AVPR1B; Z_mean_ = 2.29); on the other hand, the opioid genes made the strongest negative contributions (OPRD1; Z_mean_ = -5.60), followed by acetylcholinergic genes (CHRM1, CHRM2, CHRM3; Z_mean_ = - 4.03), one dopaminergic gene (DRD1; Z_mean_ = -1.19), and one vasopressinergic gene (AVPR1A; Z_mean_ = -0.43; Fig. 3E).

### 3.4. MACM analysis for OT effect

The bilateral thalamus, left insula, left substantia nigra, and left ventral tegmental area showed significant convergence in the MACM-A analysis of left thalamus; the left pallidum, thalamus, caudate, putamen, amygdala, and insula were activated in the MACM-A analysis of left pallidum; the MACM-A analysis of left caudate revealed significant activations within left pallidum, caudate, amygdala, thalamus, putamen, insula, and nucleus accumbens; the co-activation patterns included left insula, left inferior frontal gyrus, right caudate, left substantia nigra, and left ventral tegmental area in the MACM-A analysis of left insula (Table 2; Fig. 4). Overall, the co-activation patterns of four ROIs conjointly revealed a functionally connected thalamus-striatum-insula pathway, serving as a core functional network architecture directly modulated by OT.

**Fig. 4.**
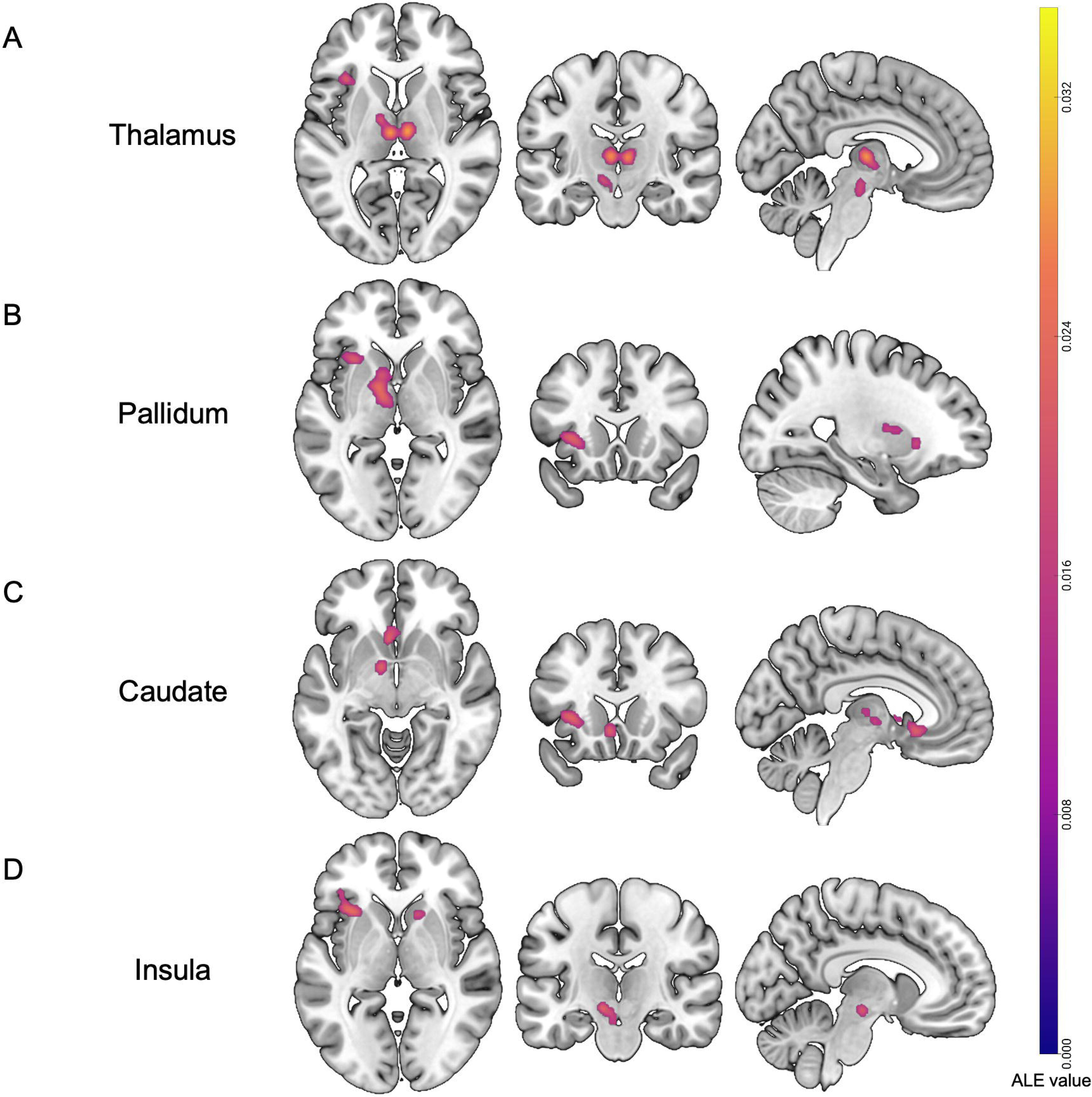
OT-affected co-activation networks revealed by MACM-A analyses. Panel A to D represent co-activation patterns of the left thalamus, pallidum, caudate, and insula, respectively. The voxel-level threshold is set at p < 0.001, uncorrected; and the cluster-level threshold is set at p < 0.05, FWE-corrected.

**Table 2.**
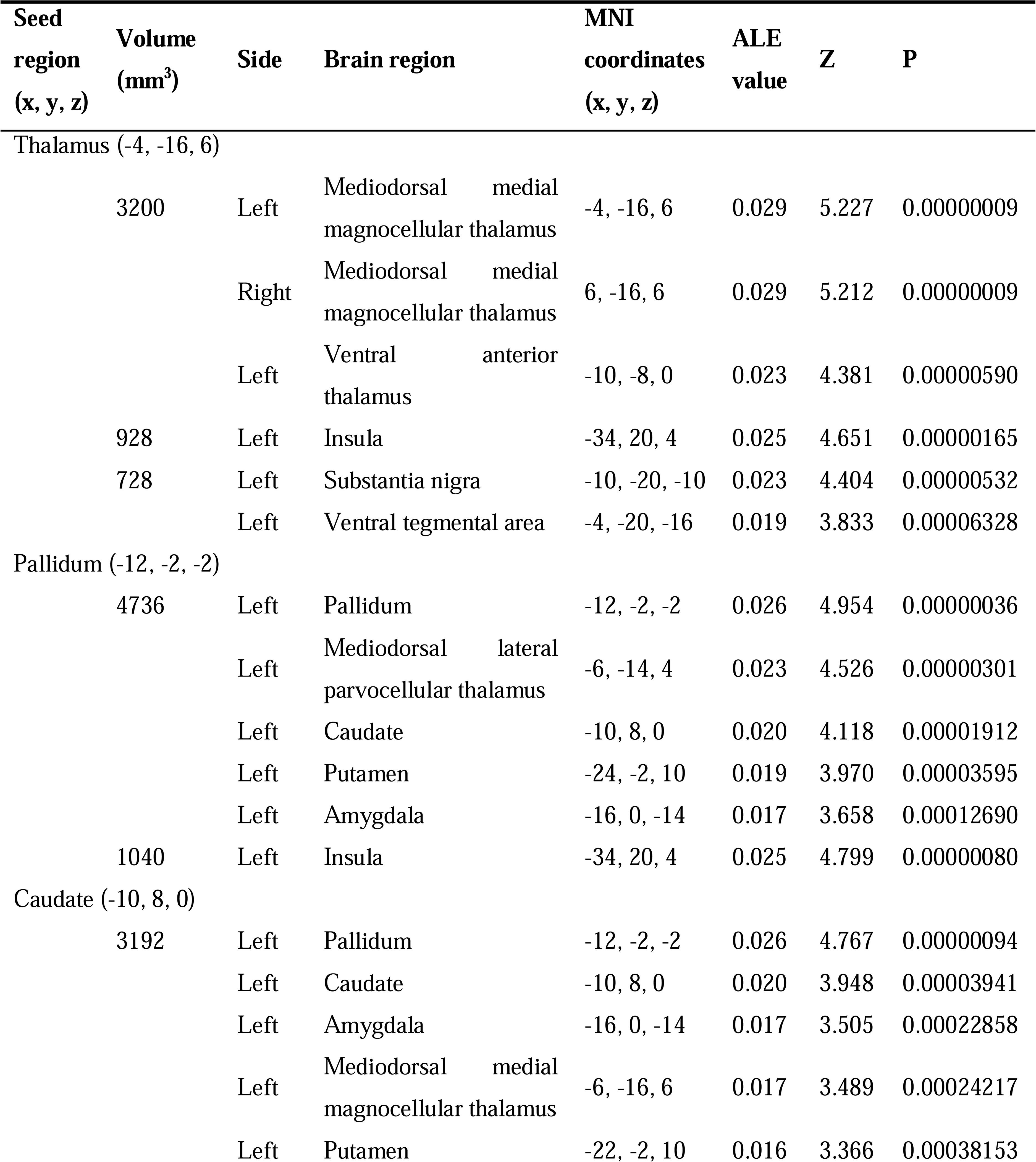
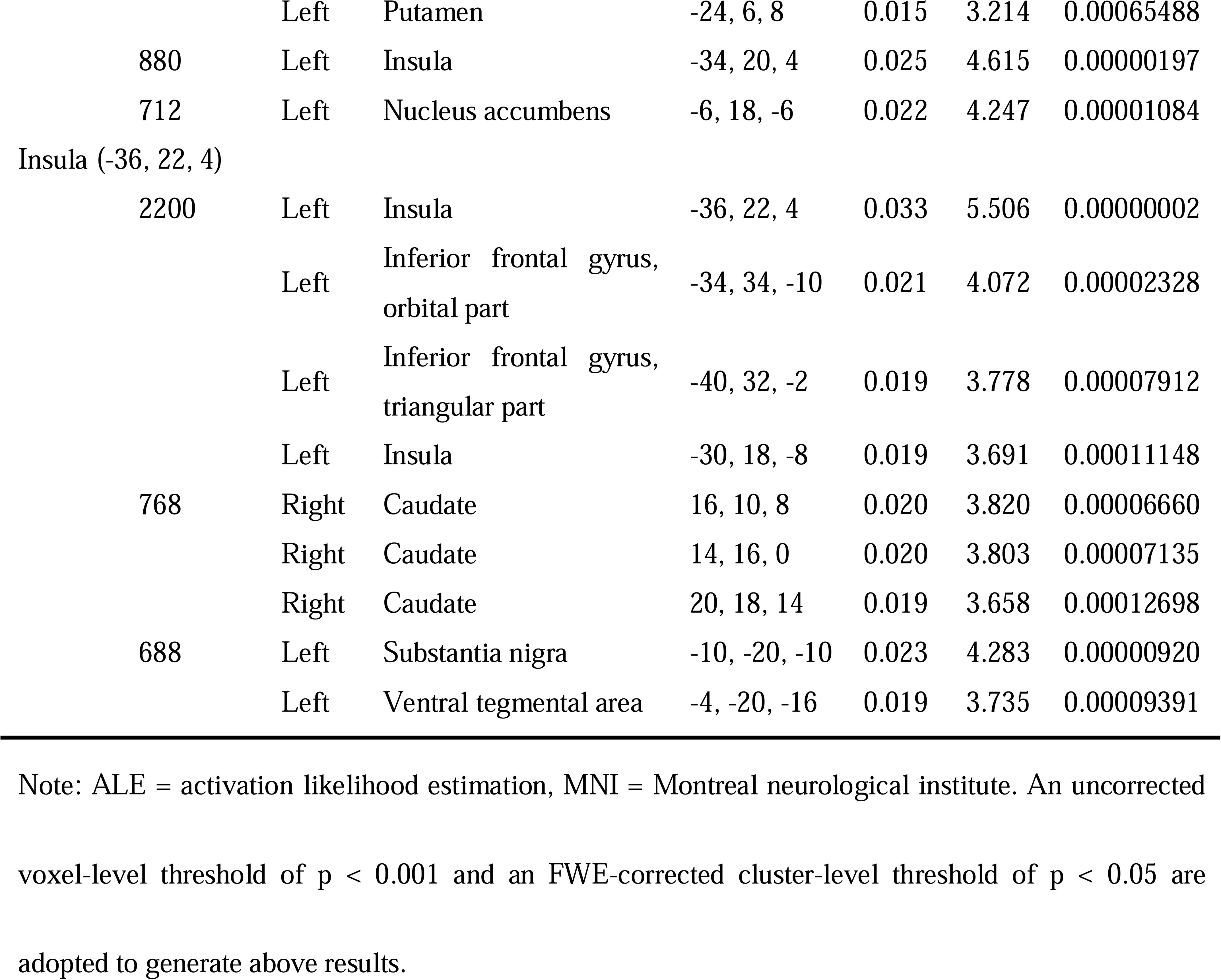
OT-affected co-activation networks revealed by MACM-A analyses.

| Seed region<br>(x, y, z) | Volume<br>(mm<sup>3</sup>) | Side | Brain region | MNI<br>coordinates<br>(x, y, z) | ALE<br>value | Z | P |
| --- | --- | --- | --- | --- | --- | --- | --- |
| Thalamus (-4, -16, 6) |  |  |  |  |  |  |  |
|  | 3200 | Left | Mediodorsal medial<br>magnocellular thalamus | -4, -16, 6 | 0.029 | 5.227 | 0.00000009 |
|  |  | Right | Mediodorsal medial<br>magnocellular thalamus | 6, -16, 6 | 0.029 | 5.212 | 0.00000009 |
|  |  | Left | Ventral anterior<br>thalamus | -10, -8, 0 | 0.023 | 4.381 | 0.00000590 |
|  | 928 | Left | Insula | -34, 20, 4 | 0.025 | 4.651 | 0.00000165 |
|  | 728 | Left | Substantia nigra | -10, -20, -10 | 0.023 | 4.404 | 0.00000532 |
|  |  | Left | Ventral tegmental area | -4, -20, -16 | 0.019 | 3.833 | 0.00006328 |
| Pallidum (-12, -2, -2) |  |  |  |  |  |  |  |
|  | 4736 | Left | Pallidum | -12, -2, -2 | 0.026 | 4.954 | 0.00000036 |
|  |  | Left | Mediodorsal lateral<br>parvocellular thalamus | -6, -14, 4 | 0.023 | 4.526 | 0.00000301 |
|  |  | Left | Caudate | -10, 8, 0 | 0.020 | 4.118 | 0.00001912 |
|  |  | Left | Putamen | -24, -2, 10 | 0.019 | 3.970 | 0.00003595 |
|  |  | Left | Amygdala | -16, 0, -14 | 0.017 | 3.658 | 0.00012690 |
|  | 1040 | Left | Insula | -34, 20, 4 | 0.025 | 4.799 | 0.00000080 |
| Caudate (-10, 8, 0) |  |  |  |  |  |  |  |
|  | 3192 | Left | Pallidum | -12, -2, -2 | 0.026 | 4.767 | 0.00000094 |
|  |  | Left | Caudate | -10, 8, 0 | 0.020 | 3.948 | 0.00003941 |
|  |  | Left | Amygdala | -16, 0, -14 | 0.017 | 3.505 | 0.00022858 |
|  |  | Left | Mediodorsal medial<br>magnocellular thalamus | -6, -16, 6 | 0.017 | 3.489 | 0.00024217 |
|  |  | Left | Putamen | -22, -2, 10 | 0.016 | 3.366 | 0.00038153 |

|  |  |  |  |  |  |  |
| --- | --- | --- | --- | --- | --- | --- |
|  | Left | Putamen | -24, 6, 8 | 0.015 | 3.214 | 0.00065488 |
| 880 | Left | Insula | -34, 20, 4 | 0.025 | 4.615 | 0.00000197 |
| 712 | Left | Nucleus accumbens | -6, 18, -6 | 0.022 | 4.247 | 0.00001084 |
| Insula (-36, 22, 4) |  |  |  |  |  |  |
| 2200 | Left | Insula | -36, 22, 4 | 0.033 | 5.506 | 0.00000002 |
|  | Left | Inferior frontal gyrus,<br>orbital part | -34, 34, -10 | 0.021 | 4.072 | 0.00002328 |
|  | Left | Inferior frontal gyrus,<br>triangular part | -40, 32, -2 | 0.019 | 3.778 | 0.00007912 |
|  | Left | Insula | -30, 18, -8 | 0.019 | 3.691 | 0.00011148 |
| 768 | Right | Caudate | 16, 10, 8 | 0.020 | 3.820 | 0.00006660 |
|  | Right | Caudate | 14, 16, 0 | 0.020 | 3.803 | 0.00007135 |
|  | Right | Caudate | 20, 18, 14 | 0.019 | 3.658 | 0.00012698 |
| 688 | Left | Substantia nigra | -10, -20, -10 | 0.023 | 4.283 | 0.00000920 |
|  | Left | Ventral tegmental area | -4, -20, -16 | 0.019 | 3.735 | 0.00009391 |

Note: ALE = activation likelihood estimation, MNI = Montreal neurological institute. An uncorrected voxel-level threshold of $p < 0.001$ and an FWE-corrected cluster-level threshold of $p < 0.05$ are adopted to generate above results.

In the MACM-B analysis, four ROIs evoked widespread co-activation patterns, including the thalamus-striatum-insula circuit uncovered in the MACM-A analysis. To be specific, the left thalamus was co-activated with right caudate, right putamen, right thalamus, right middle cingulate & paracingulate gyri, right supplementary motor area, left anterior cingulate & paracingulate gyri, left pallidum, left putamen, bilateral insula, left precentral gyrus, and left inferior frontal gyrus; the left pallidum was co-activated with right pallidum, right caudate, right thalamus, bilateral red nucleus, bilateral inferior frontal gyrus, left precentral gyrus, bilateral insula, right supplementary motor area, right middle cingulate & paracingulate gyri, left inferior parietal gyrus, right middle occipital gyrus, and right inferior occipital gyrus; the left caudate displayed co-activations with the left pallidum, right caudate, bilateral insula, bilateral thalamus, left precentral gyrus, right red nucleus, left substantia nigra, bilateral inferior frontal gyrus, right amygdala, right supplementary motor area, right middle cingulate & paracingulate gyri, right anterior cingulate & paracingulate gyri, bilateral inferior parietal gyrus, and bilateral superior parietal gyrus; the left insula showed co-activations with the right insula, bilateral inferior frontal gyrus, bilateral thalamus, bilateral middle frontal gyrus, bilateral putamen, bilateral precentral gyrus, right pallidum, right superior frontal gyrus, right red nucleus, left precentral gyrus, right supplementary motor area, left anterior cingulate & paracingulate gyri, bilateral inferior parietal gyrus, left supramarginal gyrus, bilateral superior temporal gyrus, and bilateral lobule VI of cerebellum (Fig. S5; Table S6). Taken together, we observed that the supplementary motor area, precentral gyrus, and inferior frontal gyrus were also co-activated with each ROI, apart from the core circuit.

### 3.5. Exploratory analysis

#### 3.5.1 Functional characterization of neutral effect of OT

Five separate functional characterization analyses were conducted to identify the correlations between behavioral terms and ALE maps derived from the meta-analyses for general and non-directional effect of OT as well as MACM-A analyses. Specifically, the OT-affected brain regions revealed by ALE meta-analysis were mainly correlated with reward processing (self-reported, motivation, reward, anticipation, and incentive), emotional processing (nociceptive, anxiety, emotional responses, bipolar disorder, pain, vulnerability, affective, and disgust), memory processing (memory retrieval and recognition memory), social processing (self), and attention processing (salience; Fig. 5A). Likewise, the co-activation networks of left thalamus, pallidum, caudate, and insula were characterized by reward-related, emotion-related and social process-related terms (Figs. 5B -D). Overall, behavioral decoding suggested that OT-influenced brain regions and networks were primarily involved in a variety of core functions headed by reward processing.

**Fig. 5.**
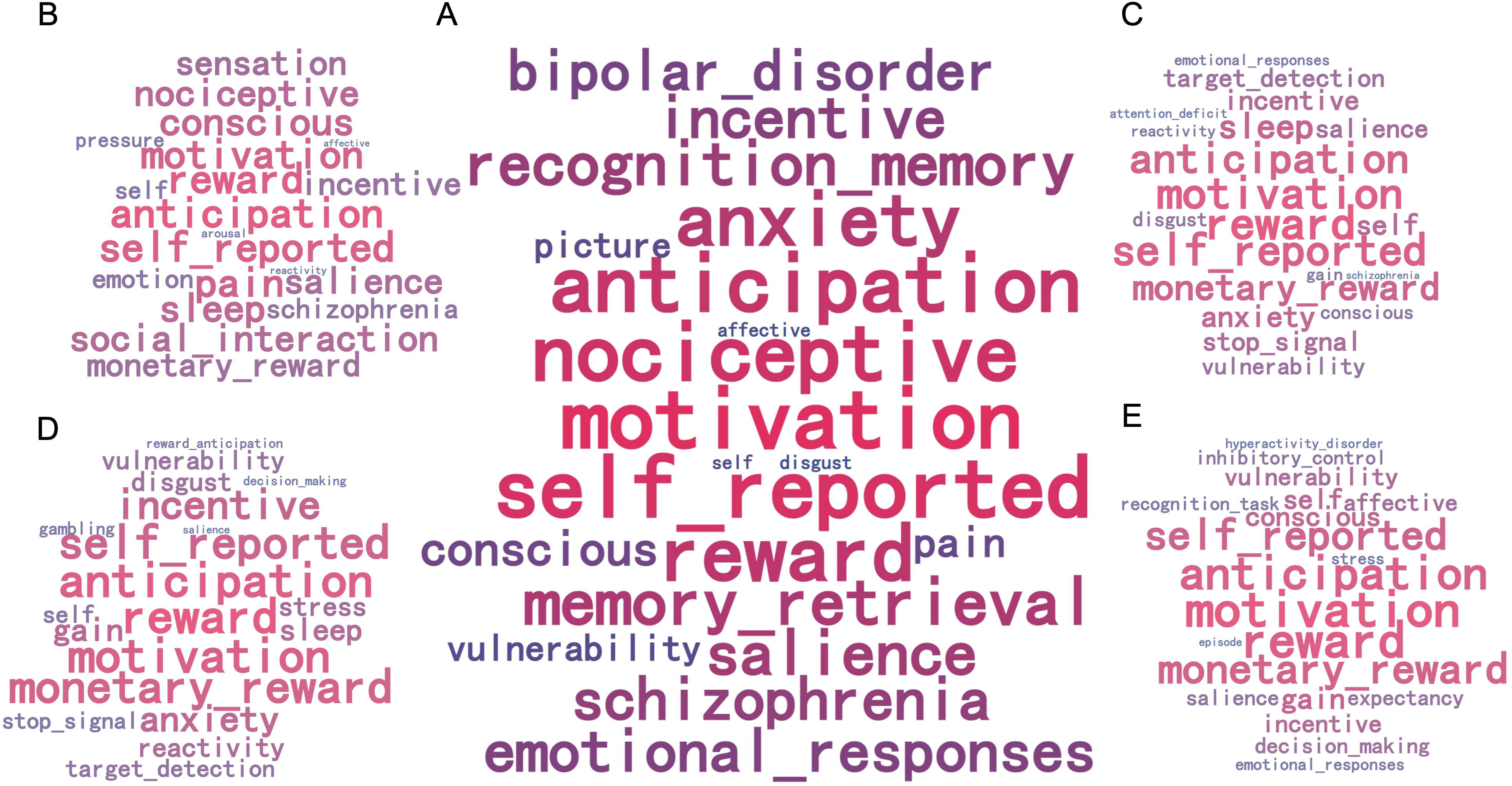
Functional characterization of OT-affected brain regions and networks. Panel A shows the top 20 behavioral terms correlated with the general and non-directional effect of OT. Panels B to D show the top 20 behavioral terms correlated with OT-affected co-activation patterns of the thalamus, pallidum, caudate, and insula, respectively. Behavioral terms are listed in order of statistical significance, thus a term with a larger font means a smaller p-value (higher significance).

#### 3.5.2 Correlation analysis with vivo molecular imaging phenotypes

The ALE map for neural effect of OT was significantly correlated with PET/SPECT map of acetylcholine receptor (M1 [Fisher’s z = 0.37, p_FDR_ = 0.008]), acetylcholine transporter (VAChTmap1 [Fisher’s z = 0.36, p_FDR_ < 0.001], VAChTmap2 [Fisher’s z = 0.35, p_FDR_ = 0.001], and VAChTmap3 [Fisher’s z = 0.33, p_FDR_ < 0.001]), dopamine receptor (D1 [Fisher’s z = 0.33, p_FDR_ = 0.001], D2map1 [Fisher’s z = 0.37, p_FDR_ < 0.001], and D2map2 [Fisher’s z = 0.38, p_FDR_ < 0.001]), dopamine transporter (DAT [Fisher’s z = 0.35, p_FDR_ < 0.001]), vesicular monoamine transporter (VMAT2 [Fisher’s z = 0.33, p_FDR_ = 0.001]), dopamine synthesis capacity (FDOPA [Fisher’s z = 0.31, p_FDR_ = 0.001]), and opioid receptor (KappaOpmap1 [Fisher’s z = 0.41, p_FDR_ = 0.014], KappaOpmap2 [Fisher’s z = 0.37, p_FDR_ = 0.019], and MORmap1 [Fisher’s z = 0.34, p_FDR_ = 0.049]). In the exploratory analysis, the ALE map for neural effect of OT was significantly correlated with PET/SPECT map of 5-hydroxytryptamine receptor (5HT4 [Fisher’s z = 0.32, p_FDR_ = 0.001] and 5HT6 [Fisher’s z = 0.43, p_FDR_ < 0.001]), 5-hydroxytryptamine transporter (SERTmap1 [Fisher’s z = 0.35, p_FDR_ < 0.001], and SERTmap2 [Fisher’s z = 0.38, p_FDR_ < 0.001]), glutamate receptor (mGluR5map3 [Fisher’s z = 0.31, p_FDR_ = 0.044]), noradrenaline transporter (NAT [Fisher’s z = 0.34, p_FDR_ < 0.001]), and N-methyl-D-aspartate receptor (NMDA [Fisher’s z = 0.35, p_FDR_ = 0.008]; Table S2; Fig. S6).

## 4. Discussion

Earlier neuroimaging meta-analyses examining the neuromodulatory effects of OT revealed inconsistent results, and the biological plausibility in terms of an overlap of the robust macroscopic effects of OT and the underlying microscopic architecture of the OT and other neurotransmitter systems remained unclear. Our updated neuroimaging meta-analytic synthesis systematically examined the general neuromodulatory effects of OT across behavioral domains and populations and specific neuromodulatory effects of OT in different behavioral domains and populations, in both an intensity and direction effect framework, and further explored associations with the microscopic architecture of the OT system as well as associations with large scale brain communication and behavioral characterization.

The main findings were as follows: (1) a comprehensive meta-analysis incorporating 75 experiments involving 2,247 participants revealed that OT consistently modulated neural activity in the left thalamus, pallidum, caudate, and insula. Effects were mirrored in the subgroup meta-analysis for healthy participants but not patient populations. In contrast, OT enhanced neural activity in the left superior temporal gyrus in the patient population. In subgroup analyses of positive valence processing, OT significantly modulated neural activity in the left pallidum and nucleus accumbens. In subgroup analyses of social processing, OT enhanced neural activity in the left thalamus and caudate; (2) transcriptomic-neuroimaging analyses underscored the biological plausibility of these results such that the expression of OT pathway genes (especially CD38) was enriched in the thalamic and basal ganglia core regions exhibiting OT modulation, and the distribution of these pathway genes was also spatially correlated with the neural effect of OT. Together with the acetylcholinergic, dopaminergic, and opioidergic genes, OT pathway genes co-predicted the general neural effect of OT, with OT pathway genes as the strongest contributors, and vasopressinergic genes as the weakest contributors; (3) MACM analyses revealed a core functional network directly influenced by OT and a secondary network indirectly influenced by OT. The core network involved thalamus-striatum-insula circuit, whereas the secondary network encompassed extensive connections across the whole brain, with notable involvement of the supplementary motor area, precentral gyrus, and inferior frontal gyrus. Finally, (4) behavioral decoding of the core regions and networks revealed strongest associations of general neural effect of OT with reward and motivational domains (e.g., anticipation, reward, incentive), and negative affective domains (e.g., anxiety, nociception).

First, our meta-analysis of the included studies revealed general and non-directional effect of OT on activity in the left thalamus, pallidum, caudate, and insula across heterogeneous tasks and populations, which is consistent with a resting-state fMRI study reporting OT-induced alterations of the fractional amplitude of intrinsic (task-free) low-frequency fluctuations in the thalamus, striatum and insula (Schneider et al., 2020). While insular involvement has been consistently reported in previous OT meta-analyses, particularly in the context of emotion processing tasks (Rocchetti et al., 2014; Wigton et al., 2015), the present convergence on the striatum and thalamus represents a novel and previously unreported finding. These regions form a core architecture engaged across diverse task states and domains (Cai et al., 2024; Dosenbach et al., 2006; Shine et al., 2019). In contrast to earlier views of the thalamus as a simple relay station for cortical information processing, a recent study has emphasized the central role of the thalamus in ongoing cortical function by acting as a primary and advanced relay (Sherman, 2016). The thalamus has also been considered to play a multifaceted role in high-level cognition (Pergola et al., 2018). The latest evidence has suggested that thalamic hubs may not be functionally (exclusively) dedicated to specific cognitive and behavioral functions, but rather provide more domain-general contributions to brain function via regulating ongoing cortical activity, connectivity, topology, and variability through a variety of mechanisms (Shine et al., 2023). These findings jointly suggest that the thalamus plays a general and multidimensional role in brain function, providing supportive evidence for OT to regulate a wide range of functions and behaviors by acting on the thalamus. Likewise, the pallidum and caudate play key roles in a wide range of functions. The pallidum critically mediates reward and motivational processes (Bhanji & Delgado, 2014; Delgado, 2007), positive emotional experiences (Malezieux et al., 2023), learning (Kaplan et al., 2020; Stephenson-Jones, 2019), and decision-making (Ottenheimer et al., 2020). Together with the caudate (Delgado et al., 2004), they form an integral reward processing hub (Delgado, 2007) in the basal ganglia, which regulate a range of emotional, cognitive and motor functions via frontal cortical-basal ganglia-thalamic circuits (Haber & Calzavara, 2009; Zhuang et al., 2023), and intrinsic communication within these circuits has been shown highly sensitive to OT across species (Bethlehem et al., 2017; Johnson et al., 2017; Zhao et al., 2019). The insula has likewise been proposed to be an integrative hub, responsible for the integration of sensory, cognitive, and affective signals (Ferraro et al., 2022b; Gu et al., 2012; Klugah-Brown et al., 2022; Kurth et al., 2010; Zhang et al., 2024a), and plays a critical role in general salience processing and autonomic activity in response to salient event, introceptive changes and cognitive effort (Ferraro et al., 2022a; Gasquoine, 2014). Behavioral decoding further confirms that the aforementioned activated regions are strongly associated with fundamental behavioral aspects such as reward, emotion, memory, social processing, and salience. Taken together, effects on these core brain regions may reflect that OT regulates basal domains associated with these systems such as salience, motivation, approach-avoidance, emotion, or allostasis (e.g., Harari-Dahan & Bernstein, 2014; Love, 2014; Quintana & Guastella, 2020; Shahrestani et al., 2013; Shamay-Tsoory & Abu-Akel, 2016), and exerts its complex effects via dynamically distributed networks in interaction with environmental and personal factors. This underlying architecture has also been largely replicated in meta-analysis for the healthy population indicating a general effect of OT.

In contrast to our hypothesis, we did not observe enhancing or attenuating effect of OT across all studies, which may reflect the fact that OT does not uniformly up-regulate/down-regulate the activity of a brain region, but rather that the effects evolve in interaction with person and context-specific effects. Regions such as the insula or the striatum have for instance demonstrated opposing effects in the direction of OT effects depending on the sex of the participants (e.g., Ma et al., 2018; Rilling et al., 2014). However, results require cautious interpretation given that the directed effects are based on smaller database of studies.

An exploratory analysis in 10 (sub-)clinical OT studies revealed that OT administration significantly enhanced neural activity in the superior temporal gyrus in the patient population. Previous studies have demonstrated functional and structural alterations of the superior temporal gyrus in a variety of patient populations that we included (De Bellis et al., 2002; Kana et al., 2016; Oliver et al., 2020; Rajarethinam et al., 2000). The superior temporal gyrus plays a crucial role in several of the behavioral domains related to OT, in particular social cognition (Bhaya-Grossman & Chang, 2022; Yi et al., 2019; Zahn et al., 2007). These findings may reflect that OT’s effects depend on the state of the participant and that it may have beneficial effects on social cognition in patient populations (for an overview on the potential therapeutic effects of OT on social cognition, see also Mercedes Perez-Rodriguez et al., 2015; Rigney et al., 2022). However, results should be interpreted with caution and await further verification due to the low number of studies included in the analysis.

The gene expression analysis revealed a dense expression of OT related genes in the thalamus and basal ganglia, which is consistent with a previous transcriptomic study (Quintana et al., 2019). Based on our meta-analytic results, we found that the robust neurofunctional effect of OT was correlated with the gene expression distribution of three OT pathway genes (CD38, OXT, and OXTR). Via their roles in the microscopic biological architecture of the OT system, these three pathway genes may mediate the effects of exogenous OT. CD38 plays a key role in OT secretion by mediating calcium (Ca²_) release (Jin et al., 2007). The OXT gene encodes the OT prepropeptide, which includes both the active nonapeptide OT and its carrier protein, neurophysin-I (Sausville et al., 1985), and the OXTR encodes a seven-transmembrane G protein-coupled receptor that can signal through either Gαi or Gαq protein pathways (Jurek & Neumann, 2018). OT receptors are the primary binding sites for OT and mediate multiple signaling pathways, including s Gαq/PLC/IP_-Ca²_, PKC, and β-arrestin-MAPK, which modulate synaptic release, plasticity, and neural oscillations, and may underlie the macroscopic neural effects of OT in subcortical regions (Gimpl & Fahrenholz, 2001; Jurek & Neumann, 2018). CD38 and OXT - responsible for synthesis and secretion of endogenous OT - have been repeatedly suggested to play a role in the effects of exogenous as well as endogenous OT (Sauer et al., 2012; Sindermann et al., 2020; Zhao et al., 2020). The association between the macroscopic effects of exogenous OT and the microscopic architecture of the OT system underscores biological plausible effects and successful target engagement by exogenous OT administered via the nasal or oral route.

Moreover, exploratory results underscored that OT might exert its complex effects via interactions with other transmitter systems such as the acetylcholinergic, dopaminergic, opioidergic, and vasopressinergic system. The results are consistent with a previous study that found some acetylcholinergic and dopaminergic genes to be highly co-expressed with OT pathway genes (Quintana et al., 2019). Several other studies have shown close interaction between the oxytocinergic system and the acetylcholinergic as well as dopaminergic systems (Paletta et al., 2022; Sauer et al., 2013; Zhang et al., 2024b). For the opioidergic system, Dal Monte and colleagues (2017) found that administering OT under opioid antagonism would boost the therapeutic efficacy of OT for enhancing social cognition. A recent review also highlighted interactions between oxytocin and the opioid system that serve to influence social behavior (Putnam & Chang, 2022). Vasopressin genes contribute comparably less to the prediction of neural effect of OT, however, the poor data quality of the vasopressin gene analyses prevents a clear interpretation of the effects (see detailed discussion in the Supplementary Material I), despite current evidence emphasizing the importance of balance between the OT system and the vasopressin system for social and emotional behavior (Neumann & Landgraf, 2012). The significant correlations between the map of the neural effects of OT and maps of acetylcholine, dopamine, and opioid receptors/transporter from vivo molecular imaging studies provided further evidence for the role of these neurotransmitter systems in effects of OT on human brain function.

On the network level the core brain regions interacted with distributed systems in the brain. The MACM-A and MACM-B analyses revealed a core network directly interacting with the core brain regions and a secondary network which may indirectly contribute to mediating the effects of OT. The MACM-A analysis identified strong inter-communication between the thalamus, striatum, and insula, indicating an integrated circuitry that is structurally promoted by dense white matter fiber bundle between the three systems (Vertes et al., 2015), and involved in functions related to salience detection, motivation, and reward (Krebs et al., 2011; Peters et al., 2016). Notably, abnormalities in this circuit have been identified as a common psychopathological characteristic of mental disorders (Peters et al., 2016), rendering this circuit a promising target for pharmacological and non-pharmacological interventions.

The MACM-B analyses further uncovered a network characterized by extensive coactivations that contained the core nodes revealed in MACM-A as well as several secondary network hubs: the supplementary motor area, precentral gyrus, and inferior frontal gyrus. An OT-modulated intrinsic interaction between these systems has been reported in previous studies (Korann et al., 2022; Zhao et al., 2019). The extensive connections of the core regions may allow OT to exert a dynamic modulation of a wide array of functional domains.

Taken together, our findings indicate that OT may exert direct effects in a core circuit with high expression of OT pathway genes and next shape different behavioral domains via effects on widespread secondary networks.

### Limitations

Compared to several previous meta-analyses, this updated meta-analysis (1) incorporated a larger sample size; (2) added connectivity-level results; (3) adopted an intensity effect model; (4) provided a functional interpretation of the neural effects of OT; and (5) explored the microtranscriptomic basis of OT administration. However, this study still has some limitations and unanswered questions. First, due to sample size constraints, sub-group meta-analyses could only be conducted in social, emotional, and reward domains, and the analyses in the patient population remains highly exploratory. Second, given the differences in mechanism and effect between chronic and acute administration of OT (Kou et al., 2022a; Kou et al., 2022b; Scheele et al., 2019), subsequent meta-analyses are required to examine the neural effects of chronic administration. Third, the current study only focused on a few genes related to several neurotransmitter systems that may interact with OT pathway genes that were included in a previous study (Quintana et al., 2019). However, the strong associations between other neurotransmitter receptors/transporters and the neural effects of OT revealed in the neurotransmitter association analyses may suggest an important role for other neurotransmitter systems in the pharmacodynamics of OT administration (Fig. S6). It is necessary to comprehensively investigate, in a data-driven way, into the key members within the neurotransmitter family that have the potential to modulate the brain effects of OT administration.

## 5. Conclusion

Summarizing, our pre-registered neuroimaging meta-analysis of exogenous OT effect in humans has identified robust effects of exogenous OT on the left thalamus, pallidum, caudate, and insula. These regions are functionally connected and form an OT-regulated “thalamus-striatum-insula” pathway. Such a core pathway represents a fundamental part of the secondary functional network indirectly regulated by OT, and may serve as the underlying framework for OT functionality in vast socio-emotional behaviors. The macroscopic effects of exogenous OT on human brain are aligned with the regional expression levels of OT pathway genes at the microscopic level and are synergistically regulated by acetylcholinergic, dopaminergic, opioidergic, and vasopressinergic systems. Findings suggest that OT may regulate human behavior via effects on a core set of biologically plausible systems with complex effects emerging via interaction with other transmitter systems and interactions with larger scale brain systems.

## Supplementary material

The supplementary materials for this article can be found online.

## Supporting information

Supplementary Material III

## Data Availability

All data produced in the present study are available upon reasonable request to the authors

## Acknowledgment

The present study is supported by the Ministry of Science and Technology of China (STI 2030-Major Projects 2022ZD0208500), National Natural Science Foundation of China (NSFC 82271583), the Hong Kong University Grants Council (GRF 17615525), The University of Hong Kong seed funding and start-up schemes (2407102536). We express our sincere gratitude to the Allen Institute for Brain Science for providing gene expression data.

## Data availability

The raw gene expression data are available at the official website of AHBA database (http://human.brain-map.org). The original codes used in this study are available at: https://osf.io/jp6zs; https://github.com/BMHLab/AHBAprocessing; https://github.com/helab207/Alterations-in-Connectome-Dynamics-in-ASD-main/tree/main/Code.

## References

Abdi, H., & Williams, L. J., 2013. Partial least squares methods: partial least squares correlation and partial least square regression. In B. Reisfeld & A. N. Mayeno (Eds.), Computational toxicology: Volume ii (pp. 549–579). Humana Press. 10.1007/978-1-62703-059-5_23.

ArnatkevicDiūtė, A., Fulcher, B. D., & Fornito, A., 2019. A practical guide to linking brain-wide gene expression and neuroimaging data. Neuroimage 189, 353–367. 10.1016/j.neuroimage.2019.01.011.

Bartz, J. A., Zaki, J., Bolger, N., & Ochsner, K. N., 2011. Social effects of oxytocin in humans: context and person matter. Trends Cogn. Sci. 15(7), 301–309. 10.1016/j.tics.2011.05.002.

Baumgartner, T., Heinrichs, M., Vonlanthen, A., Fischbacher, U., & Fehr, E., 2008. Oxytocin shapes the neural circuitry of trust and trust adaptation in humans. Neuron 58(4), 639–650. 10.1016/j.neuron.2008.04.009.

Benjamini, Y., & Hochberg, Y., 1995. Controlling the false discovery rate: a practical and powerful approach to multiple testing. J. R. Stat. Soc. Series B Stat. Methodol. 57(1), 289–300. 10.1111/j.2517-6161.1995.tb02031.x.

Bethlehem, R. A. I., Lombardo, M. V., Lai, M. C., Auyeung, B., Crockford, S. K., Deakin, J., Soubramanian, S., Sule, A., Kundu, P., Voon, V., & Baron-Cohen, S., 2017. Intranasal oxytocin enhances intrinsic corticostriatal functional connectivity in women. Transl. Psychiatry 7(4), e1099–e1099. 10.1038/tp.2017.72.

Bethlehem, R. A. I., van Honk, J., Auyeung, B., & Baron-Cohen, S., 2013. Oxytocin, brain physiology, and functional connectivity: a review of intranasal oxytocin fMRI studies. Psychoneuroendocrinology 38(7), 962–974. 10.1016/j.psyneuen.2012.10.011.

Bhanji, J. P., & Delgado, M. R., 2014. The social brain and reward: social information processing in the human striatum. WIREs Cogn. Sci. 5(1), 61–73. 10.1002/wcs.1266.

Bhaya-Grossman, I., & Chang, E. F., 2022. Speech computations of the human superior temporal gyrus. Annu. Rev. Psychol. 73(1), 79–102. 10.1146/annurev-psych-022321-035256.

Burt, J. B., Helmer, M., Shinn, M., Anticevic, A., & Murray, J. D., 2020. Generative modeling of brain maps with spatial autocorrelation. Neuroimage 220, 117038. 10.1016/j.neuroimage.2020.117038.

Cai, W., Taghia, J., & Menon, V., 2024. A multi-demand operating system underlying diverse cognitive tasks. Nat. Commun. 15(1), 2185. 10.1038/s41467-024-46511-5.

Cauda, F., Cavanna, A. E., D’agata, F., Sacco, K., Duca, S., & Geminiani, G. C., 2011. Functional connectivity and coactivation of the nucleus accumbens: a combined functional connectivity and structure-based meta-analysis. J. Cogn. Neurosci. 23(10), 2864–2877. 10.1162/jocn.2011.21624.

Cavalli, J., Ruttorf, M., Pahi, M. R., Zidda, F., Flor, H., & Nees, F., 2017. Oxytocin differentially modulates pavlovian cue and context fear acquisition. Soc. Cogn. Affect. Neurosci. 12(6), 976–983. 10.1093/scan/nsx028.

Chen, X., Hackett, P. D., DeMarco, A. C., Feng, C., Stair, S., Haroon, E., Ditzen, B., Pagnoni, G., & Rilling, J. K., 2016. Effects of oxytocin and vasopressin on the neural response to unreciprocated cooperation within brain regions involved in stress and anxiety in men and women. Brain Imaging Behav. 10(2), 581–593. 10.1007/s11682-015-9411-7.

Chen, X., Nishitani, S., Haroon, E., Smith, A. K., & Rilling, J. K., 2020a. OXTR methylation modulates exogenous oxytocin effects on human brain activity during social interaction. Genes, Brain and Behav. 19(1), e12555. 10.1111/gbb.12555.

Chen, Y., Becker, B., Zhang, Y., Cui, H., Du, J., Wernicke, J., Montag, C., Kendrick, K. M., & Yao, S., 2020b. Oxytocin increases the pleasantness of affective touch and orbitofrontal cortex activity independent of valence. Eur. Neuropsychopharmacol. 39, 99–110. 10.1016/j.euroneuro.2020.08.003.

Cole, M. W., Bassett, D. S., Power, J. D., Braver, T. S., & Petersen, S. E., 2014. Intrinsic and task-evoked network architectures of the human brain. Neuron 83(1), 238–251. 10.1016/j.neuron.2014.05.014.

Dal Monte, O., Piva, M., Anderson, K. M., Tringides, M., Holmes, A. J., & Chang, S. W. C., 2017. Oxytocin under opioid antagonism leads to supralinear enhancement of social attention. Proc. Natl. Acad. Sci. U.S.A. 114(20), 5247–5252. 10.1073/pnas.1702725114.

De Bellis, M. D., Keshavan, M. S., Shifflett, H., Iyengar, S., Dahl, R. E., Axelson, D. A., Birmaher, B., Hall, J., Moritz, G., & Ryan, N. D., 2002. Superior temporal gyrus volumes in pediatric generalized anxiety disorder. Biol. Psychiatry 51(7), 553–562. 10.1016/S0006-3223(01)01375-0.

Delgado, M. R., 2007. RewardDrelated responses in the human striatum. Ann. N. Y. Acad. Sci. 1104(1), 70–88. 10.1196/annals.1390.002.

Delgado, M. R., Stenger, V. A., & Fiez, J. A., 2004. Motivation-dependent responses in the human caudate nucleus. Cereb. Cortex 14(9), 1022–1030. 10.1093/cercor/bhh062.

Domes, G., Lischke, A., Berger, C., Grossmann, A., Hauenstein, K., Heinrichs, M., & Herpertz, S. C., 2010. Effects of intranasal oxytocin on emotional face processing in women. Psychoneuroendocrinology 35(1), 83–93. 10.1016/j.psyneuen.2009.06.016.

Dosenbach, N. U. F., Visscher, K. M., Palmer, E. D., Miezin, F. M., Wenger, K. K., Kang, H. C., Burgund, E. D., Grimes, A. L., Schlaggar, B. L., & Petersen, S. E., 2006. A core system for the implementation of task sets. Neuron 50(5), 799–812. 10.1016/j.neuron.2006.04.031.

Dukart, J., Holiga, S., Rullmann, M., Lanzenberger, R., Hawkins, P. C. T., Mehta, M. A., Hesse, S., Barthel, H., Sabri, O., Jech, R., & Eickhoff, S. B., 2021. JuSpace: a tool for spatial correlation analyses of magnetic resonance imaging data with nuclear imaging derived neurotransmitter maps. Hum. Brain Mapp. 42(3), 555–566. 10.1002/hbm.25244.

Eckstein, M., Scheele, D., Patin, A., Preckel, K., Becker, B., Walter, A., Domschke, K., Grinevich, V., Maier, W., & Hurlemann, R., 2016. Oxytocin facilitates pavlovian fear learning in males. Neuropsychopharmacology 41(4), 932–939. 10.1038/npp.2015.245.

Eckstein, M., Scheele, D., Weber, K., Stoffel-Wagner, B., Maier, W., & Hurlemann, R., 2014. Oxytocin facilitates the sensation of social stress. Hum. Brain Mapp. 35(9), 4741–4750. 10.1002/hbm.22508.

Eickhoff, S. B., Bzdok, D., Laird, A. R., Kurth, F., & Fox, P. T., 2012. Activation likelihood estimation meta-analysis revisited. Neuroimage 59(3), 2349–2361. 10.1016/j.neuroimage.2011.09.017.

Eickhoff, S. B., Jbabdi, S., Caspers, S., Laird, A. R., Fox, P. T., Zilles, K., & Behrens, T. E. J., 2010. Anatomical and functional connectivity of cytoarchitectonic areas within the human parietal operculum. J. Neurosci. 30(18), 6409–6421. 10.1523/jneurosci.5664-09.2010.

Eickhoff, S. B., Laird, A. R., Fox, P. M., Lancaster, J. L., & Fox, P. T., 2017. Implementation errors in the GingerALE software: description and recommendations. Hum. Brain Mapp. 38(1), 7–11. 10.1002/hbm.23342.

Eickhoff, S. B., Laird, A. R., Grefkes, C., Wang, L. E., Zilles, K., & Fox, P. T., 2009. Coordinate-based activation likelihood estimation meta-analysis of neuroimaging data: a random-effects approach based on empirical estimates of spatial uncertainty. Hum. Brain Mapp. 30(9), 2907–2926. 10.1002/hbm.20718.

Eickhoff, S. B., Nichols, T. E., Laird, A. R., Hoffstaedter, F., Amunts, K., Fox, P. T., Bzdok, D., & Eickhoff, C. R., 2016. Behavior, sensitivity, and power of activation likelihood estimation characterized by massive empirical simulation. Neuroimage 137, 70–85. 10.1016/j.neuroimage.2016.04.072.

Fan, S., Zhang, Y., Qian, R., Hu, J., Zheng, H., Dai, W., Ji, Y., Wu, Y., Xie, X., Xu, S., Ji, G.-J., Tian, Y., & Wang, K., 2025. Genetic and molecular basis of abnormal BOLD signaling variability in patients with major depressive disorder after electroconvulsive therapy. Transl. Psychiatry 15(1), 117. 10.1038/s41398-025-03330-6.

Fascher, M., Nowaczynski, S., & Muehlhan, M., 2024. Substance use disorders are characterised by increased voxel-wise intrinsic measures in sensorimotor cortices: an ALE meta-analysis. Neurosci. Biobehav. Rev. 162, 105712. 10.1016/j.neubiorev.2024.105712.

Feifel, D., Macdonald, K., Nguyen, A., Cobb, P., Warlan, H., Galangue, B., Minassian, A., Becker, O., Cooper, J., Perry, W., Lefebvre, M., Gonzales, J., & Hadley, A., 2010. Adjunctive intranasal oxytocin reduces symptoms in schizophrenia patients. Biol. Psychiatry 68(7), 678–680. 10.1016/j.biopsych.2010.04.039.

Feldman, R., Monakhov, M., Pratt, M., & Ebstein, R. P., 2016. Oxytocin pathway genes: evolutionary ancient system impacting on human affiliation, sociality, and psychopathology. Biol. Psychiatry 79(3), 174–184. 10.1016/j.biopsych.2015.08.008.

Feng, C., Eickhoff, S. B., Li, T., Wang, L., Becker, B., Camilleri, J. A., Hétu, S., & Luo, Y., 2021. Common brain networks underlying human social interactions: evidence from large-scale neuroimaging meta-analysis. Neurosci. Biobehav. Rev. 126, 289–303. 10.1016/j.neubiorev.2021.03.025.

Ferraro, S., Klugah-Brown, B., Tench, C. R., Bazinet, V., Bore, M. C., Nigri, A., Demichelis, G., Bruzzone, M. G., Palermo, S., Zhao, W., Yao, S., Jiang, X., Kendrick, K. M., & Becker, B., 2022a. The central autonomic system revisited – convergent evidence for a regulatory role of the insular and midcingulate cortex from neuroimaging meta-analyses. Neurosci. Biobehav. Rev. 142, 104915. 10.1016/j.neubiorev.2022.104915.

Ferraro, S., Klugah-Brown, B., Tench, C. R., Yao, S., Nigri, A., Demichelis, G., Pinardi, C., Bruzzone, M. G., & Becker, B., 2022b. Dysregulated anterior insula reactivity as robust functional biomarker for chronic pain—meta-analytic evidence from neuroimaging studies. Hum. Brain Mapp. 43(3), 998–1010. 10.1002/hbm.25702.

Froemke, R. C., & Young, L. J., 2021. Oxytocin, neural plasticity, and social behavior. Annu. Rev. Neurosci. 44(1), 359–381. 10.1146/annurev-neuro-102320-102847.

Fu, K., Xu, S., Zhang, Z., Liu, D., Xu, T., Zhang, Y., Zhou, F., Zhang, X., Lan, C., Wang, J., Wang, L., He, J., Kendrick, K. M., Biswal, B., Yao, D., Liang, Z., Zhao, W., & Becker, B., 2025. Oxytocin reduces subjective fear in naturalistic social contexts via enhancing top-down middle cingulate amygdala regulation and brain-wide fear representations. Adv. Sci., e03251. 10.1002/advs.202503251.

Gan, R., Qiu, Y., Liao, J., Zhang, Y., Wu, J., Peng, X., Lee, T. M.-c., & Huang, R., 2024. Mapping the mentalizing brain: an ALE meta-analysis to differentiate the representation of social scenes and ages on theory of mind. Neurosci. Biobehav. Rev. 167, 105918. 10.1016/j.neubiorev.2024.105918.

Gan, X., Zhou, X., Li, J., Jiao, G., Jiang, X., Biswal, B., Yao, S., Klugah-Brown, B., & Becker, B., 2022. Common and distinct neurofunctional representations of core and social disgust in the brain: coordinate-based and network meta-analyses. Neurosci. Biobehav. Rev. 135, 104553. 10.1016/j.neubiorev.2022.104553.

Gasquoine, P. G., 2014. Contributions of the insula to cognition and emotion. Neuropsychol. Rev. 24(2), 77–87. 10.1007/s11065-014-9246-9.

Geladi, P., & Kowalski, B. R., 1986. Partial least-squares regression: a tutorial. Anal. Chim. Acta 185, 1–17. 10.1016/0003-2670(86)80028-9.

Gimpl, G., & Fahrenholz, F., 2001. The oxytocin receptor system: structure, function, and regulation. Physiol. Rev. 81(2), 629–683. 10.1152/physrev.2001.81.2.629.

Giovanna, G., Damiani, S., Fusar-Poli, L., Rocchetti, M., Brondino, N., de Cagna, F., Mori, A., & Politi, P., 2020. Intranasal oxytocin as a potential therapeutic strategy in post-traumatic stress disorder: a systematic review. Psychoneuroendocrinology 115, 104605. 10.1016/j.psyneuen.2020.104605.

Grace, S. A., Rossell, S. L., Heinrichs, M., Kordsachia, C., & Labuschagne, I., 2018. Oxytocin and brain activity in humans: a systematic review and coordinate-based meta-analysis of functional MRI studies. Psychoneuroendocrinology 96, 6–24. 10.1016/j.psyneuen.2018.05.031.

Gronroos, M., Kivikoski, A., & Kyostila, J., 1962. On the effect of intranasally administered oxytocin (syntocin) on lactation. Ann. Chir. Gynaecol. Fenn. 51, 377–381.

Gu, X., Liu, X., Van Dam, N. T., Hof, P. R., & Fan, J., 2012. Cognition–emotion integration in the anterior insular cortex. Cereb. Cortex 23(1), 20–27. 10.1093/cercor/bhr367.

Gunasekera, B., Davies, C., Blest-Hopley, G., Veronese, M., Ramsey, N. F., Bossong, M. G., Radua, J., Bhattacharyya, S., Pretzsch, C., McAlonan, G., Walter, C., Lötsch, J., Freeman, T., Curran, V., Battistella, G., Fornari, E., Filho, G. B., Crippa, J. A., Duran, F., & Zuardi, A. W., 2022. Task-independent acute effects of delta-9-tetrahydrocannabinol on human brain function and its relationship with cannabinoid receptor gene expression: a neuroimaging meta-regression analysis. Neurosci. Biobehav. Rev. 140, 104801. 10.1016/j.neubiorev.2022.104801.

Haber, S. N., & Calzavara, R., 2009. The cortico-basal ganglia integrative network: the role of the thalamus. Brain Res. Bull. 78(2), 69–74. 10.1016/j.brainresbull.2008.09.013.

Habets, P. C., Mclain, C., & Meijer, O. C., 2021. Brain areas affected by intranasal oxytocin show higher oxytocin receptor expression. Eur. J. Neurosci. 54(7), 6374–6381. 10.1111/ejn.15447.

Harari-Dahan, O., & Bernstein, A., 2014. A general approach-avoidance hypothesis of oxytocin: accounting for social and non-social effects of oxytocin. Neurosci. Biobehav. Rev. 47, 506–519. 10.1016/j.neubiorev.2014.10.007.

Hawrylycz, M. J., Lein, E. S., Guillozet-Bongaarts, A. L., Shen, E. H., Ng, L., Miller, J. A., van de Lagemaat, L. N., Smith, K. A., Ebbert, A., Riley, Z. L., Abajian, C., Beckmann, C. F., Bernard, A., Bertagnolli, D., Boe, A. F., Cartagena, P. M., Chakravarty, M. M., Chapin, M., Chong, J., Dalley, R. A., Daly, B. D., Dang, C., Datta, S., Dee, N., Dolbeare, T. A., Faber, V., Feng, D., Fowler, D. R., Goldy, J., Gregor, B. W., Haradon, Z., Haynor, D. R., Hohmann, J. G., Horvath, S., Howard, R. Jeromin, A., Jochim, J. M., Kinnunen, M., Lau, C., Lazarz, E. T., Lee, C., Lemon, T. A., Li, L., Li, Y., Morris, J. A., Overly, C. C., Parker, P. D., Parry, S. E., Reding, M., Royall, J. J., Schulkin, J., Sequeira, P. A., Slaughterbeck, C. R., Smith, S. C., Sodt, A. J., Sunkin, S. M., Swanson, B. E., Vawter, M. P., Williams, D., Wohnoutka, P., Zielke, H. R., Geschwind, D. H., Hof, P. R., Smith, S. M., Koch, C., Grant, S. G. N., & Jones, A. R., 2012. An anatomically comprehensive atlas of the adult human brain transcriptome. Nature 489(7416), 391–399. 10.1038/nature11405.

Hermesch, A. C., Kernberg, A. S., Layoun, V. R., & Caughey, A. B., 2024. Oxytocin: physiology, pharmacology, and clinical application for labor management. Am. J. Obstet. Gynecol. 230(3, Supplement), S729-S739. 10.1016/j.ajog.2023.06.041.

Hikosaka, O., Kim, H. F., Yasuda, M., & Yamamoto, S., 2014. Basal ganglia circuits for reward value– guided behavior. Annu. Rev. Neurosci. 37(1), 289–306. 10.1146/annurev-neuro-071013-013924.

Hu, J., Qi, S., Becker, B., Luo, L., Gao, S., Gong, Q., Hurlemann, R., & Kendrick, K. M., 2015. Oxytocin selectively facilitates learning with social feedback and increases activity and functional connectivity in emotional memory and reward processing regions. Hum. Brain Mapp. 36(6), 2132–2146. 10.1002/hbm.22760.

Ide, J. S., Nedic, S., Wong, K. F., Strey, S. L., Lawson, E. A., Dickerson, B. C., Wald, L. L., La Camera, G., & Mujica-Parodi, L. R., 2018. Oxytocin attenuates trust as a subset of more general reinforcement learning, with altered reward circuit functional connectivity in males. Neuroimage 174, 35–43. 10.1016/j.neuroimage.2018.02.035.

Insel, T., Cuthbert, B., Garvey, M., Heinssen, R., Pine, D. S., Quinn, K., Sanislow, C., & Wang, P., 2010. Research domain criteria (RDoC): toward a new classification framework for research on mental disorders. Am. J. Psychiatry 167(7), 748–751. 10.1176/appi.ajp.2010.09091379.

Jansen, M., Lockwood, P. L., Cutler, J., & de Bruijn, E. R. A., 2023. l-DOPA and oxytocin influence the neurocomputational mechanisms of self-benefitting and prosocial reinforcement learning. Neuroimage 270, 119983. 10.1016/j.neuroimage.2023.119983.

Jiang, X., Ma, X., Geng, Y., Zhao, Z., Zhou, F., Zhao, W., Yao, S., Yang, S., Zhao, Z., Becker, B., & Kendrick, K. M., 2021. Intrinsic, dynamic and effective connectivity among large-scale brain networks modulated by oxytocin. Neuroimage 227, 117668. 10.1016/j.neuroimage.2020.117668.

Jin, D., Liu, H.-X., Hirai, H., Torashima, T., Nagai, T., Lopatina, O., Shnayder, N. A., Yamada, K., Noda, M., Seike, T., Fujita, K., Takasawa, S., Yokoyama, S., Koizumi, K., Shiraishi, Y., Tanaka, S., Hashii, M., Yoshihara, T., Higashida, K., Islam, M. S., Yamada, N., Hayashi, K., Noguchi, N., Kato, I., Okamoto, H., Matsushima, A., Salmina, A., Munesue, T., Shimizu, N., Mochida, S., Asano, M., & Higashida, H., 2007. CD38 is critical for social behaviour by regulating oxytocin secretion. Nature 446(7131), 41–45. 10.1038/nature05526.

Johnson, Z. V., Walum, H., Xiao, Y., Riefkohl, P. C., & Young, L. J., 2017. Oxytocin receptors modulate a social salience neural network in male prairie voles. Horm. Behav. 87, 16–24. 10.1016/j.yhbeh.2016.10.009.

Jurek, B., & Neumann, I. D., 2018. The oxytocin receptor: from intracellular signaling to behavior. Physiol. Rev. 98(3), 1805–1908. 10.1152/physrev.00031.2017.

Kana, R. K., Patriquin, M. A., Black, B. S., Channell, M. M., & Wicker, B., 2016. Altered medial frontal and superior temporal response to implicit processing of emotions in autism. Autism Res. 9(1), 55–66. 10.1002/aur.1496.

Kanat, M., Heinrichs, M., Schwarzwald, R., & Domes, G., 2015. Oxytocin attenuates neural reactivity to masked threat cues from the eyes. Neuropsychopharmacology 40(2), 287–295. 10.1038/npp.2014.183.

Kaplan, A., Mizrahi-Kliger, A. D., Israel, Z., Adler, A., & Bergman, H., 2020. Dissociable roles of ventral pallidum neurons in the basal ganglia reinforcement learning network. Nat. Neurosci. 23(4), 556–564. 10.1038/s41593-020-0605-y.

Kendrick, K. M., Guastella, A. J., & Becker, B., 2018. Overview of human oxytocin research. In R. Hurlemann & V. Grinevich (Eds.), Behavioral pharmacology of neuropeptides: oxytocin (pp. 321–348). Springer International Publishing. 10.1007/7854_2017_19.

Kirsch, P., Esslinger, C., Chen, Q., Mier, D., Lis, S., Siddhanti, S., Gruppe, H., Mattay, V. S., Gallhofer, B., & Meyer-Lindenberg, A., 2005. Oxytocin modulates neural circuitry for social cognition and fear in humans. J. Neurosci. 25(49), 11489–11493. 10.1523/jneurosci.3984-05.2005.

Klugah-Brown, B., Wang, P., Jiang, Y., Becker, B., Hu, P., Uddin, L. Q., & Biswal, B., 2022. Structural–functional connectivity mapping of the insular cortex: a combined data-driven and meta-analytic topic mapping. Cereb. Cortex 33(5), 1726–1738. 10.1093/cercor/bhac168.

Klugah-Brown, B., Zhou, X., Pradhan, B. K., Zweerings, J., Mathiak, K., Biswal, B., & Becker, B., 2021. Common neurofunctional dysregulations characterize obsessive–compulsive, substance use, and gaming disorders—an activation likelihood meta-analysis of functional imaging studies. Addict. Biol. 26(4), e12997. 10.1111/adb.12997.

Korann, V., Jacob, A., Lu, B., Devi, P., Thonse, U., Nagendra, B., Maria Chacko, D., Dey, A., Padmanabha, A., Shivakumar, V., Dawn Bharath, R., Kumar, V., Varambally, S., Venkatasubramanian, G., Deshpande, G., & Rao, N. P., 2022. Effect of intranasal oxytocin on resting-state effective connectivity in schizophrenia. Schizophr. Bull. 48(5), 1115–1124. 10.1093/schbul/sbac066.

Kosfeld, M., Heinrichs, M., Zak, P. J., Fischbacher, U., & Fehr, E., 2005. Oxytocin increases trust in humans. Nature 435(7042), 673–676. 10.1038/nature03701.

Kou, J., Zhang, Y., Zhou, F., Gao, Z., Yao, S., Zhao, W., Li, H., Lei, Y., Gao, S., Kendrick, K. M., & Becker, B., 2022a. Anxiolytic effects of chronic intranasal oxytocin on neural responses to threat are dose-frequency dependent. Psychother. Psychosom. 91(4), 253–264. 10.1159/000521348.

Kou, J., Zhang, Y., Zhou, F., Sindermann, C., Montag, C., Becker, B., & Kendrick, K. M., 2022b. A randomized trial shows dose-frequency and genotype may determine the therapeutic efficacy of intranasal oxytocin. Psychol. Med. 52(10), 1959–1968. 10.1017/S0033291720003803.

Krebs, R. M., Boehler, C. N., Roberts, K. C., Song, A. W., & Woldorff, M. G., 2011. The involvement of the dopaminergic midbrain and cortico-striatal-thalamic circuits in the integration of reward prospect and attentional task demands. Cereb. Cortex 22(3), 607–615. 10.1093/cercor/bhr134.

Krienen, F. M., Yeo, B. T. T., & Buckner, R. L., 2014. Reconfigurable task-dependent functional coupling modes cluster around a core functional architecture. Philos. Trans. R. Soc. Lond. B Biol. Sci. 369(1653), 20130526. 10.1098/rstb.2013.0526.

Kurth, F., Zilles, K., Fox, P. T., Laird, A. R., & Eickhoff, S. B., 2010. A link between the systems: Functional differentiation and integration within the human insula revealed by meta-analysis. Brain Struct. Funct. 214(5), 519–534. 10.1007/s00429-010-0255-z.

Laird, A. R., Eickhoff, S. B., Rottschy, C., Bzdok, D., Ray, K. L., & Fox, P. T., 2013. Networks of task co-activations. Neuroimage 80, 505–514. 10.1016/j.neuroimage.2013.04.073.

Laird, A. R., Robinson, J. L., McMillan, K. M., Tordesillas-Gutiérrez, D., Moran, S. T., Gonzales, S. M., Ray, K. L., Franklin, C., Glahn, D. C., Fox, P. T., & Lancaster, J. L., 2010. Comparison of the disparity between Talairach and MNI coordinates in functional neuroimaging data: Validation of the lancaster transform. Neuroimage 51(2), 677–683. 10.1016/j.neuroimage.2010.02.048.

Lan, C., Chen, Y., Zhang, Y., Kou, J., Huang, L., Xu, T., Yang, X., Xu, D., Yang, W., Kendrick, K. M., & Zhao, W., 2023. Oral oxytocin facilitates responses to emotional faces in reward and emotional-processing networks in females. Neuroendocrinology 113(9), 957–970. 10.1159/000531064.

Lan, C., Kou, J., Liu, Q., Qing, P., Zhang, X., Song, X., Xu, D., Zhang, Y., Chen, Y., Zhou, X., Kendrick, K. M., & Zhao, W., 2024. Oral oxytocin blurs sex differences in amygdala responses to emotional scenes. Biol. Psychiatry Cogn. Neurosci. Neuroimaging 9(10), 1028–1038. 10.1016/j.bpsc.2024.05.010.

Lan, C., Liu, C., Li, K., Zhao, Z., Yang, J., Ma, Y., Scheele, D., Yao, S., Kendrick, K. M., & Becker, B., 2022. Oxytocinergic modulation of stress-associated amygdala-hippocampus pathways in humans is mediated by serotonergic mechanisms. Int. J. Neuropsychopharmacol. 25(10), 807–817. 10.1093/ijnp/pyac037.

Landgraf, R., & Neumann, I. D., 2004. Vasopressin and oxytocin release within the brain: a dynamic concept of multiple and variable modes of neuropeptide communication. Front. Neuroendocrinol. 25(3), 150–176. 10.1016/j.yfrne.2004.05.001.

Langner, R., & Camilleri, J. A., 2021. Meta-analytic connectivity modelling (MACM): a tool for assessing region-specific functional connectivity patterns in task-constrained states. In V. A. Diwadkar & S. B. Eickhoff (Eds.), Brain network dysfunction in neuropsychiatric illness: Methods, applications, and implications (pp. 93–104). Springer International Publishing. 10.1007/978-3-030-59797-9_5.

Le, J., Zhang, L., Zhao, W., Zhu, S., Lan, C., Kou, J., Zhang, Q., Zhang, Y., Li, Q., Chen, Z., Fu, M., Montag, C., Zhang, R., Yang, W., Becker, B., & Kendrick, K. M., 2022. Infrequent intranasal oxytocin followed by positive social interaction improves symptoms in autistic children: a pilot randomized clinical trial. Psychother. Psychosom. 91(5), 335–347. 10.1159/000524543.

Li, J., Long, Z., Sheng, W., Du, L., Qiu, J., Chen, H., & Liao, W., 2024. Transcriptomic similarity informs neuromorphic deviations in depression biotypes. Biol. Psychiatry 95(5), 414–425. 10.1016/j.biopsych.2023.08.003.

Liu, C., Lan, C., Li, K., Zhou, F., Yao, S., Xu, L., Yang, N., Zhou, X., Yang, J., Yong, X., Ma, Y., Scheele, D., Kendrick, K. M., & Becker, B., 2021. Oxytocinergic modulation of threat-specific amygdala sensitization in humans is critically mediated by serotonergic mechanisms. Biol. Psychiatry Cogn. Neurosci. Neuroimaging 6(11), 1081–1089. 10.1016/j.bpsc.2021.04.009.

Liu, C., Zhuang, K., Zeitlen, D. C., Chen, Q., Wang, X., Feng, Q., Beaty, R. E., & Qiu, J., 2024. Neural, genetic, and cognitive signatures of creativity. Commun. Biol. 7(1), 1324. 10.1038/s42003-024-07007-6.

Liu, Y., Li, S., Lin, W., Li, W., Yan, X., Wang, X., Pan, X., Rutledge, R. B., & Ma, Y., 2019a. Oxytocin modulates social value representations in the amygdala. Nat. Neurosci. 22(4), 633–641. 10.1038/s41593-019-0351-1.

Liu, Z., Rolls, E. T., Liu, Z., Zhang, K., Yang, M., Du, J., Gong, W., Cheng, W., Dai, F., Wang, H., Ugurbil, K., Zhang, J., & Feng, J., 2019b. Brain annotation toolbox: exploring the functional and genetic associations of neuroimaging results. Bioinformatics 35(19), 3771–3778. 10.1093/bioinformatics/btz128.

Long, H., Wu, H., Sun, C., Xu, X., Yang, X.-H., Xiao, J., Lv, M., Chen, Q., & Fan, M., 2024. Biological mechanism of sex differences in mental rotation: evidence from multimodal MRI, transcriptomic and receptor/transporter data. Neuroimage 304, 120955. 10.1016/j.neuroimage.2024.120955.

Love, T. M., 2014. Oxytocin, motivation and the role of dopamine. Pharmacol. Biochem. Behav. 119, 49–60. 10.1016/j.pbb.2013.06.011.

Luhman, L. A., 1963. The effect of intranasal oxytocin on lactation. Obstet. Gynecol. 21(6), 713–717.

Luo, Y., Dong, D., Huang, H., Zhou, J., Zuo, X., Hu, J., He, H., Jiang, S., Duan, M., Yao, D., & Luo, C., 2023. associating multimodal neuroimaging abnormalities with the transcriptome and neurotransmitter signatures in schizophrenia. Schizophr. Bull. 49(6), 1554–1567. 10.1093/schbul/sbad047.

Ma, X., Zhao, W., Luo, R., Zhou, F., Geng, Y., Xu, L., Gao, Z., Zheng, X., Becker, B., & Kendrick, K. M., 2018. Sex- and context-dependent effects of oxytocin on social sharing. Neuroimage 183, 62–72. 10.1016/j.neuroimage.2018.08.004.

Ma, Y., 2015. Neuropsychological mechanism underlying antidepressant effect: a systematic meta-analysis. Mol. Psychiatry 20(3), 311–319. 10.1038/mp.2014.24.

Malezieux, M., Klein, A. S., & Gogolla, N., 2023. Neural circuits for emotion. Annu. Rev. Neurosci. 46(1), 211–231. 10.1146/annurev-neuro-111020-103314.

Maliske, L. Z., Schurz, M., & Kanske, P., 2023. Interactions within the social brain: co-activation and connectivity among networks enabling empathy and theory of mind. Neurosci. Biobehav. Rev. 147, 105080. 10.1016/j.neubiorev.2023.105080.

McTeague, L. M., Rosenberg, B. M., Lopez, J. W., Carreon, D. M., Huemer, J., Jiang, Y., Chick, C. F., Eickhoff, S. B., & Etkin, A., 2020. Identification of common neural circuit disruptions in emotional processing across psychiatric disorders. Am. J. Psychiatry 177(5), 411–421. 10.1176/appi.ajp.2019.18111271.

Mellentin, A. I., Finn, S. W., Skøt, L., Thaysen-Petersen, D., Mistarz, N., Fink-Jensen, A., & Nielsen, D. G., 2023. The effectiveness of oxytocin for treating substance use disorders: a systematic review of randomized placebo-controlled trials. Neurosci. Biobehav. Rev. 151, 105185. 10.1016/j.neubiorev.2023.105185.

Menon, R., & Neumann, I. D., 2023. Detection, processing and reinforcement of social cues: regulation by the oxytocin system. Nat. Rev. Neurosci. 24(12), 761–777. 10.1038/s41583-023-00759-w.

Mercedes Perez-Rodriguez, M., Mahon, K., Russo, M., Ungar, A. K., & Burdick, K. E., 2015. Oxytocin and social cognition in affective and psychotic disorders. Eur. Neuropsychopharmacol. 25(2), 265-282. 10.1016/j.euroneuro.2014.07.012.

Meyer-Lindenberg, A., Domes, G., Kirsch, P., & Heinrichs, M., 2011. Oxytocin and vasopressin in the human brain: social neuropeptides for translational medicine. Nat. Rev. Neurosci. 12(9), 524–538. 10.1038/nrn3044.

Müller, V. I., Cieslik, E. C., Laird, A. R., Fox, P. T., Radua, J., Mataix-Cols, D., Tench, C. R., Yarkoni, T., Nichols, T. E., Turkeltaub, P. E., Wager, T. D., & Eickhoff, S. B., 2018. Ten simple rules for neuroimaging meta-analysis. Neurosci. Biobehav. Rev. 84, 151–161. 10.1016/j.neubiorev.2017.11.012.

Müller, V. I., Cieslik, E. C., Serbanescu, I., Laird, A. R., Fox, P. T., & Eickhoff, S. B., 2017. Altered brain activity in unipolar depression revisited: meta-analyses of neuroimaging studies. JAMA Psychiatry 74(1), 47–55. 10.1001/jamapsychiatry.2016.2783.

Namkung, H., Kim, S.-H., & Sawa, A., 2017. The insula: an underestimated brain area in clinical neuroscience, psychiatry, and neurology. Trends Neurosci. 40(4), 200–207. 10.1016/j.tins.2017.02.002.

Nelson, A. B., & Kreitzer, A. C., 2014. Reassessing models of basal ganglia function and dysfunction. Annu. Rev. Neurosci. 37(1), 117–135. 10.1146/annurev-neuro-071013-013916.

Neumann, I. D., & Landgraf, R., 2012. Balance of brain oxytocin and vasopressin: implications for anxiety, depression, and social behaviors. Trends Neurosci. 35(11), 649–659. 10.1016/j.tins.2012.08.004.

Oliver, L. D., Stewart, C., Coleman, K., Kryklywy, J. H., Bartha, R., Mitchell, D. G. V., & Finger, E. C., 2020. Neural effects of oxytocin and mimicry in frontotemporal dementia. Neurology 95(19), e2635–e2647. 10.1212/WNL.0000000000010933.

Ottenheimer, D. J., Wang, K., Tong, X., Fraser, K. M., Richard, J. M., & Janak, P. H., 2020. Reward activity in ventral pallidum tracks satiety-sensitive preference and drives choice behavior. Sci. Adv. 6(45), eabc9321. 10.1126/sciadv.abc9321.

Page, M. J., Moher, D., Bossuyt, P. M., Boutron, I., Hoffmann, T. C., Mulrow, C. D., Shamseer, L., Tetzlaff, J. M., Akl, E. A., Brennan, S. E., Chou, R., Glanville, J., Grimshaw, J. M., Hróbjartsson, A., Lalu, M. M., Li, T., Loder, E. W., Mayo-Wilson, E., McDonald, S., McGuinness, L. A., Stewart, L. A., Thomas, J., Tricco, A. C., Welch, V. A., Whiting, P., & McKenzie, J. E., 2021. PRISMA 2020 explanation and elaboration: updated guidance and exemplars for reporting systematic reviews. BMJ 372, n160. 10.1136/bmj.n160.

Paletta, P., Bass, N., Kavaliers, M., & Choleris, E., 2022. The role of oxytocin in shaping complex social behaviours: possible interactions with other neuromodulators. Philos. Trans. R. Soc. Lond. B Biol. Sci. 377(1858), 20210058. 10.1098/rstb.2021.0058.

Pergola, G., Danet, L., Pitel, A.-L., Carlesimo, G. A., Segobin, S., Pariente, J., Suchan, B., Mitchell, A. S., & Barbeau, E. J., 2018. The regulatory role of the human mediodorsal thalamus. Trends Cogn. Sci. 22(11), 1011–1025. 10.1016/j.tics.2018.08.006.

Peters, S. K., Dunlop, K., & Downar, J., 2016. Cortico-striatal-thalamic loop circuits of the salience network: a central pathway in psychiatric disease and treatment. Front. Syst. Neurosci. 10(104). 10.3389/fnsys.2016.00104.

Pincus, D., Kose, S., Arana, A., Johnson, K., Morgan, P., Borckardt, J., Herbsman, T., Hardaway, F., George, M., Panksepp, J., & Nahas, Z., 2010. Inverse effects of oxytocin on attributing mental activity to others in depressed and healthy subjects: a double-blind placebo-controlled fMRI study. Front. Psychiatry 1(134). 10.3389/fpsyt.2010.00134.

Putnam, P. T., & Chang, S. W. C., 2022. Interplay between the oxytocin and opioid systems in regulating social behaviour. Philos. Trans. R. Soc. Lond. B Biol. Sci. 377(1858), 20210050. 10.1098/rstb.2021.0050.

Quintana, D. S., & Guastella, A. J., 2020. An allostatic theory of oxytocin. Trends Cogn. Sci. 24(7), 515–528. 10.1016/j.tics.2020.03.008.

Quintana, D. S., Guastella, A. J., Westlye, L. T., & Andreassen, O. A., 2016. The promise and pitfalls of intranasally administering psychopharmacological agents for the treatment of psychiatric disorders. Mol. Psychiatry 21(1), 29–38. 10.1038/mp.2015.166.

Quintana, D. S., Lischke, A., Grace, S., Scheele, D., Ma, Y., & Becker, B., 2021. Advances in the field of intranasal oxytocin research: lessons learned and future directions for clinical research. Mol. Psychiatry 26(1), 80–91. 10.1038/s41380-020-00864-7.

Quintana, D. S., Rokicki, J., van der Meer, D., Alnæs, D., Kaufmann, T., Córdova-Palomera, A., Dieset, I., Andreassen, O. A., & Westlye, L. T., 2019. Oxytocin pathway gene networks in the human brain. Nat. Commun. 10(1), 668. 10.1038/s41467-019-08503-8.

Rajarethinam, R. P., DeQuardo, J. R., Nalepa, R., & Tandon, R., 2000. Superior temporal gyrus in schizophrenia: a volumetric magnetic resonance imaging study. Schizophr. Res. 41(2), 303–312. 10.1016/S0920-9964(99)00083-3.

Reimann, G. M., Küppers, V., Camilleri, J. A., Hoffstaedter, F., Langner, R., Laird, A. R., Fox, P. T., Spiegelhalder, K., Eickhoff, S. B., & Tahmasian, M., 2023. Convergent abnormality in the subgenual anterior cingulate cortex in insomnia disorder: a revisited neuroimaging meta-analysis of 39 studies. Sleep Med. Rev. 71, 101821. 10.1016/j.smrv.2023.101821.

Rigney, N., de Vries, G. J., Petrulis, A., & Young, L. J., 2022. Oxytocin, vasopressin, and social behavior: from neural circuits to clinical opportunities. Endocrinology 163(9). 10.1210/endocr/bqac111.

Rikhye, R. V., Wimmer, R. D., & Halassa, M. M., 2018. Toward an integrative theory of thalamic function. Annu. Rev. Neurosci. 41(1), 163–183. 10.1146/annurev-neuro-080317-062144.

Rilling, J. K., DeMarco, A. C., Hackett, P. D., Chen, X., Gautam, P., Stair, S., Haroon, E., Thompson, R., Ditzen, B., Patel, R., & Pagnoni, G., 2014. Sex differences in the neural and behavioral response to intranasal oxytocin and vasopressin during human social interaction. Psychoneuroendocrinology 39, 237–248. 10.1016/j.psyneuen.2013.09.022.

Robinson, J. L., Laird, A. R., Glahn, D. C., Blangero, J., Sanghera, M. K., Pessoa, L., Fox, P. M., Uecker, A., Friehs, G., Young, K. A., Griffin, J. L., Lovallo, W. R., & Fox, P. T., 2012. The functional connectivity of the human caudate: an application of meta-analytic connectivity modeling with behavioral filtering. Neuroimage 60(1), 117–129. 10.1016/j.neuroimage.2011.12.010.

Rocchetti, M., Radua, J., Paloyelis, Y., Xenaki, L.-A., Frascarelli, M., Caverzasi, E., Politi, P., & Fusar-Poli, P., 2014. Neurofunctional maps of the ‘maternal brain’ and the effects of oxytocin: a multimodal voxel-based meta-analysis. Psychiatry Clin. Neurosci. 68(10), 733–751. 10.1111/pcn.12185.

Rokicki, J., Kaufmann, T., de Lange, A.-M. G., van der Meer, D., Bahrami, S., Sartorius, A. M., Haukvik, U. K., Steen, N. E., Schwarz, E., Stein, D. J., Nærland, T., Andreassen, O. A., Westlye, L. T., & Quintana, D. S., 2022. Oxytocin receptor expression patterns in the human brain across development. Neuropsychopharmacology 47(8), 1550–1560. 10.1038/s41386-022-01305-5.

Rolls, E. T., Huang, C.-C., Lin, C.-P., Feng, J., & Joliot, M., 2020. Automated anatomical labelling atlas 3. Neuroimage 206, 116189. 10.1016/j.neuroimage.2019.116189.

Sauer, C., Montag, C., Reuter, M., & Kirsch, P., 2013. Imaging oxytocin × dopamine interactions: an epistasis effect of CD38 and COMT gene variants influences the impact of oxytocin on amygdala activation to social stimuli. Front. Neurosci. 7(45). 10.3389/fnins.2013.00045.

Sauer, C., Montag, C., Wörner, C., Kirsch, P., & Reuter, M., 2012. Effects of a common variant in the CD38 gene on social processing in an oxytocin challenge study: possible links to autism. Neuropsychopharmacology 37(6), 1474–1482. 10.1038/npp.2011.333.

Sausville, E., Carney, D., & Battey, J., 1985. The human vasopressin gene is linked to the oxytocin gene and is selectively expressed in a cultured lung cancer cell line. J Biol Chem 260(18), 10236–10241.

Scheele, D., Lieberz, J., Goertzen-Patin, A., Engels, C., Schneider, L., Stoffel-Wagner, B., Becker, B., & Hurlemann, R., 2019. Trauma disclosure moderates the effects of oxytocin on intrusions and neural responses to fear. Psychother. Psychosom. 88(1), 61–63. 10.1159/000496056.

Schneider, I., Schmitgen, M. M., Boll, S., Roth, C., Nees, F., Usai, K., Herpertz, S. C., & Wolf, R. C., 2020. Oxytocin modulates intrinsic neural activity in patients with chronic low back pain. Eur. J. Pain 24(5), 945–955. 10.1002/ejp.1543.

Seeley, S. H., Chou, Y.-h., & O’Connor, M.-F., 2018. Intranasal oxytocin and OXTR genotype effects on resting state functional connectivity: a systematic review. Neurosci. Biobehav. Rev. 95, 17–32. 10.1016/j.neubiorev.2018.09.011.

Shahrestani, S., Kemp, A. H., & Guastella, A. J., 2013. The impact of a single administration of intranasal oxytocin on the recognition of basic emotions in humans: a meta-analysis. Neuropsychopharmacology 38(10), 1929–1936. 10.1038/npp.2013.86.

Shamay-Tsoory, S. G., & Abu-Akel, A., 2016. The social salience hypothesis of oxytocin. Biol. Psychiatry 79(3), 194–202. 10.1016/j.biopsych.2015.07.020.

Sherman, S. M., 2016. Thalamus plays a central role in ongoing cortical functioning. Nat. Neurosci. 19(4), 533–541. 10.1038/nn.4269.

Shine, J. M., Breakspear, M., Bell, P. T., Ehgoetz Martens, K. A., Shine, R., Koyejo, O., Sporns, O., & Poldrack, R. A., 2019. Human cognition involves the dynamic integration of neural activity and neuromodulatory systems. Nat. Neurosci. 22(2), 289–296. 10.1038/s41593-018-0312-0.

Shine, J. M., Lewis, L. D., Garrett, D. D., & Hwang, K., 2023. The impact of the human thalamus on brain-wide information processing. Nat. Rev. Neurosci. 24(7), 416–430. 10.1038/s41583-023-00701-0.

Sindermann, C., Luo, R., Becker, B., Kendrick, K. M., & Montag, C., 2020. The role of oxytocin on self-serving lying. Brain Behav. 10(2), e01518. 10.1002/brb3.1518.

Spetter, M. S., Feld, G. B., Thienel, M., Preissl, H., Hege, M. A., & Hallschmid, M., 2018. Oxytocin curbs calorie intake via food-specific increases in the activity of brain areas that process reward and establish cognitive control. Sci. Rep. 8(1), 2736. 10.1038/s41598-018-20963-4.

Stephenson-Jones, M., 2019. Pallidal circuits for aversive motivation and learning. Curr. Opin. Behav. Sci. 26, 82–89. 10.1016/j.cobeha.2018.09.015.

Tononi, G., & Edelman, G. M., 1998. Consciousness and complexity. Science 282(5395), 1846–1851. 10.1126/science.282.5395.1846.

Tully, J., Sethi, A., Griem, J., Paloyelis, Y., Craig, M. C., Williams, S. C. R., Murphy, D., Blair, R. J., & Blackwood, N., 2023. Oxytocin normalizes the implicit processing of fearful faces in psychopathy: a randomized crossover study using fMRI. Nat. Ment. Health 1(6), 420–427. 10.1038/s44220-023-00067-3.

Turkeltaub, P. E., Eickhoff, S. B., Laird, A. R., Fox, M., Wiener, M., & Fox, P., 2012. Minimizing within-experiment and within-group effects in activation likelihood estimation meta-analyses. Hum. Brain Mapp. 33(1), 1–13. 10.1002/hbm.21186.

Utter, A. A., & Basso, M. A., 2008. The basal ganglia: an overview of circuits and function. Neurosci. Biobehav. Rev. 32(3), 333–342. 10.1016/j.neubiorev.2006.11.003.

Valtcheva, S., Issa, H. A., Bair-Marshall, C. J., Martin, K. A., Jung, K., Zhang, Y., Kwon, H.-B., & Froemke, R. C., 2023. Neural circuitry for maternal oxytocin release induced by infant cries. Nature 621(7980), 788–795. 10.1038/s41586-023-06540-4.

Vertes, R. P., Linley, S. B., & Hoover, W. B., 2015. Limbic circuitry of the midline thalamus. Neurosci. Biobehav. Rev. 54, 89–107. 10.1016/j.neubiorev.2015.01.014.

Vigneaud, V. d., Ressler, C., Swan, C. J. M., Roberts, C. W., Katsoyannis, P. G., & Gordon, S., 1953. The synthesis of an octapeptide amide with the hormonal activity of oxytocin. J. Am. Chem. Soc. 75(19), 4879–4880.

Wang, D., Yan, X., Li, M., & Ma, Y., 2017. Neural substrates underlying the effects of oxytocin: a quantitative meta-analysis of pharmaco-imaging studies. Soc. Cogn. Affect. Neurosci. 12(10), 1565–1573. 10.1093/scan/nsx085.

Wigton, R., Radua, J., Allen, P., Averbeck, B., Meyer-Lindenberg, A., McGuire, P., Shergill, S. S., & Fusar-Poli, P., 2015. Neurophysiological effects of acute oxytocin administration: systematic review and meta-analysis of placebo-controlled imaging studies. J. Psychiatry Neurosci. 40(1), E1–E22. 10.1503/jpn.130289.

Wu, Q., Huang, Q., Liu, C., & Wu, H., 2022. Oxytocin modulates social brain network correlations in resting and task state. Cereb. Cortex 33(7), 3607–3620. 10.1093/cercor/bhac295.

Xiao, S., Ebner, N. C., Manzouri, A., Li, T.-Q., Cortes, D. S., Månsson, K. N. T., & Fischer, H., 2024. Age-dependent effects of oxytocin in brain regions enriched with oxytocin receptors. Psychoneuroendocrinology 160, 106666. 10.1016/j.psyneuen.2023.106666.

Xie, Y., Xu, Z., Xia, M., Liu, J., Shou, X., Cui, Z., Liao, X., & He, Y., 2022. Alterations in connectome dynamics in autism spectrum disorder: A harmonized mega- and meta-analysis study using the autism brain imaging data exchange dataset. Biol. Psychiatry 91(11), 945–955. 10.1016/j.biopsych.2021.12.004.

Xin, F., Zhou, F., Zhou, X., Ma, X., Geng, Y., Zhao, W., Yao, S., Dong, D., Biswal, B. B., Kendrick, K. M., & Becker, B., 2018. Oxytocin modulates the intrinsic dynamics between attention-related large-scale networks. Cereb. Cortex 31(3), 1848–1860. 10.1093/cercor/bhy295.

Xu, T., Chen, Z., Zhou, X., Wang, L., Zhou, F., Yao, D., Zhou, B., & Becker, B., 2024. The central renin–angiotensin system: a genetic pathway, functional decoding, and selective target engagement characterization in humans. Proc. Natl. Acad. Sci. U.S.A. 121(8), e2306936121. 10.1073/pnas.2306936121.

Xu, X., Liu, C., Zhou, X., Chen, Y., Gao, Z., Zhou, F., Kou, J., Becker, B., & Kendrick, K. M., 2019. Oxytocin facilitates self-serving rather than altruistic tendencies in competitive social interactions via orbitofrontal cortex. Int. J. Neuropsychopharmacol. 22(8), 501–512. 10.1093/ijnp/pyz028.

Yao, S., Becker, B., Zhao, W., Zhao, Z., Kou, J., Ma, X., Geng, Y., Ren, P., & Kendrick, K. M., 2018a. Oxytocin modulates attention switching between interoceptive signals and external social cues. Neuropsychopharmacology 43(2), 294–301. 10.1038/npp.2017.189.

Yao, S., Zhao, W., Geng, Y., Chen, Y., Zhao, Z., Ma, X., Xu, L., Becker, B., & Kendrick, K. M., 2018b. Oxytocin facilitates approach behavior to positive social stimuli via decreasing anterior insula activity. Int. J. Neuropsychopharmacol. 21(10), 918–925. 10.1093/ijnp/pyy068.

Yarkoni, T., Poldrack, R. A., Nichols, T. E., Van Essen, D. C., & Wager, T. D., 2011. Large-scale automated synthesis of human functional neuroimaging data. Nat. Methods 8(8), 665–670. 10.1038/nmeth.1635.

Yi, H. G., Leonard, M. K., & Chang, E. F., 2019. The encoding of speech sounds in the superior temporal gyrus. Neuron 102(6), 1096–1110. 10.1016/j.neuron.2019.04.023.

Zahn, R., Moll, J., Krueger, F., Huey, E. D., Garrido, G., & Grafman, J., 2007. Social concepts are represented in the superior anterior temporal cortex. Proc. Natl. Acad. Sci. U.S.A. 104(15), 6430–6435. 10.1073/pnas.0607061104.

Zhang, R., Deng, H., & Xiao, X., 2024a. The insular cortex: an interface between sensation, emotion and cognition. Neurosci. Bull. 40(11), 1763–1773. 10.1007/s12264-024-01211-4.

Zhang, Y., Karadas, M., Liu, J., Gu, X., Vöröslakos, M., Li, Y., Tsien, R. W., & Buzsáki, G., 2024b. Interaction of acetylcholine and oxytocin neuromodulation in the hippocampus. Neuron 112(11), 1862–1875. 10.1016/j.neuron.2024.02.021.

Zhao, W., Le, J., Liu, Q., Zhu, S., Lan, C., Zhang, Q., Zhang, Y., Li, Q., Kou, J., Yang, W., Zhang, R., Becker, B., Zhang, L., & Kendrick, K. M., 2024. A clustering approach identifies an autism spectrum disorder subtype more responsive to chronic oxytocin treatment. Transl. Psychiatry 14(1), 312. 10.1038/s41398-024-03025-4.

Zhao, W., Luo, R., Sindermann, C., Li, J., Wei, Z., Zhang, Y., Liu, C., Le, J., Quintana, D. S., Montag, C., Becker, B., & Kendrick, K. M., 2020. Oxytocin modulation of self-referential processing is partly replicable and sensitive to oxytocin receptor genotype. Prog. Neuro-Psychopharmacol. Biol. Psychiatry 96, 109734. 10.1016/j.pnpbp.2019.109734.

Zhao, W., Yao, S., Li, Q., Geng, Y., Ma, X., Luo, L., Xu, L., & Kendrick, K. M., 2016. Oxytocin blurs the self-other distinction during trait judgments and reduces medial prefrontal cortex responses. Hum. Brain Mapp. 37(7), 2512–2527. 10.1002/hbm.23190.

Zhao, Z., Ma, X., Geng, Y., Zhao, W., Zhou, F., Wang, J., Markett, S., Biswal, B. B., Ma, Y., Kendrick, K. M., & Becker, B., 2019. Oxytocin differentially modulates specific dorsal and ventral striatal functional connections with frontal and cerebellar regions. Neuroimage 184, 781–789. 10.1016/j.neuroimage.2018.09.067.

Zhuang, Q., Qiao, L., Xu, L., Yao, S., Chen, S., Zheng, X., Li, J., Fu, M., Li, K., Vatansever, D., Ferraro, S., Kendrick, K. M., & Becker, B., 2023. The right inferior frontal gyrus as pivotal node and effective regulator of the basal ganglia-thalamocortical response inhibition circuit. Psychoradiology 3, kkad016. 10.1093/psyrad/kkad016.

