## Supplementary Material I for "Effects of exogenous oxytocin on human brain function are regulated by oxytocin gene expression: a meta-analysis of 20 years of oxytocin neuroimaging and transcriptomic analyses"

1. **Method**
   1.1. **Search formula**
        1.1.1. **EBSCO (n = 1,295)**

TI ((BOLD OR blood oxygen level-dependent OR neuroimaging OR neural imaging OR brain imaging OR pharmaco-imaging OR functional imaging OR MRI OR fMRI OR magnetic resonance imaging)) AND TI ((oxytocin OR syntocinon OR pitocin))

KW ((BOLD OR blood oxygen level-dependent OR neuroimaging OR neural imaging OR brain imaging OR pharmaco-imaging OR functional imaging OR MRI OR fMRI OR magnetic resonance imaging)) AND KW ((oxytocin OR syntocinon OR pitocin))

AB ((BOLD OR blood oxygen level-dependent OR neuroimaging OR neural imaging OR brain imaging OR pharmaco-imaging OR functional imaging OR MRI OR fMRI OR magnetic resonance imaging)) AND AB ((oxytocin OR syntocinon OR pitocin))

1.1.2. **PubMed (n = 479)**

((((((((((BOLD[Title/Abstract]) OR (blood oxygen level-dependent[Title/Abstract])) OR (neuroimaging[Title/Abstract])) OR (neural imaging[Title/Abstract])) OR (brain imaging[Title/Abstract])) OR (pharmaco-imaging[Title/Abstract])) OR (functional imaging[Title/Abstract])) OR (MRI[Title/Abstract])) OR (fMRI[Title/Abstract])) OR (magnetic resonance imaging[Title/Abstract])) AND (((oxytocin[Title/Abstract]) OR (syntocinon[Title/Abstract])) OR (pitocin[Title/Abstract]))

1.1.3. **Scopus (n = 1,218)**

TITLE-ABS-KEY(BOLD OR “blood oxygen level-dependent” OR neuroimaging OR “neural imaging” OR “brain imaging” OR pharmaco-imaging OR “functional imaging” OR MRI OR fMRI OR “magnetic resonance imaging”) AND TITLE-ABS-KEY(oxytocin OR syntocinon OR pitocin) AND PUBYEAR > 2004

1.1.4. **ScienceDirect (n = 362)**

(BOLD OR blood oxygen level-dependent OR neuroimaging OR neural imaging OR brain imaging OR pharmaco-imaging OR functional imaging OR MRI OR fMRI OR magnetic resonance imaging) AND (oxytocin OR syntocinon OR pitocin)

1.1.5. **Web of Science (n = 1,492)**

(TS = (BOLD OR blood oxygen level-dependent OR neuroimaging OR neural imaging OR brain imaging OR pharmaco-imaging OR functional imaging OR MRI OR fMRI OR magnetic resonance imaging)) AND (TS = (oxytocin OR syntocinon OR pitocin))

1.2. **Pipeline of MACM-B**

There were several following steps for MACM-B. (1) Definition of ROIs: we first defined four seeds, i.e., spherical regions in standard MNI space centered on the four peak coordinates activated in the pooled meta-analysis with a radius of 5 mm, which was done in the MarsBaR toolbox (https://marsbar-toolbox.github.io/marsbar/; Brett et al., 2002). It is worth noting that a variation in ROI size would bring about a corresponding change in the results (Fox et al., 2014), however, there has been no evidence yet to shed light on the recommended ROI size at the time of the MACM analysis. Considering that subcortical regions were reported in our primary meta-analysis, a smaller ROI with a 5-mm radius was appropriate because a smaller ROI would be more likely to be localized within the same anatomical region. Toward a more comprehensive exploration of the co-activated patterns associated with the OT effect, however, we further performed MACM analyses of ROIs with a radius of 6 mm and 8 mm as exploratory results. (2) Definition of inclusion and exclusion criteria: “Activations: Activations only”, “Diagnosis: Normals”, “Imaging Modality: fMRI” and “Context: Normal Mapping” were set in Sleuth 3.0.4 (https://brainmap.org/sleuth/), which were commonly used filtering conditions (Fascher et al., 2024). (3) Selection of eligible experiments: following the above conditions, the dataset for MACM-B analysis was identified for each of the 4 coordinates (left Thalamus: 62 experiments, 1035 foci, 1957 subjects; left Pallidum: 36 experiments, 658 foci, 579 subjects; left Caudate: 95 experiments, 1555 foci, 1744 subjects; left Insula: 193 experiments, 3400 foci, 3574 subjects) from the functional neuroimaging sub-database (3,406 papers, 114 paradigm classes, 76,016 subjects, 16,901 experiments, and 131,598 locations) in the BrainMap database. (4) Data extraction: all coordinates identified in step 3 were exported to a txt file with experiment as the organization criteria and MNI as standard space. (5) Implementation of MACM: after removing all replicated experiments, meta-analyses were re-executed in GingerALE to identify co-activated patterns associated with these peak coordinates (Langner & Camilleri, 2021; Robinson et al., 2012). Despite the differences between MACM-A and MACM-B in terms of the datasets included, they both employed the same ALE-based secondary meta-analysis methodology and adopted the same statistical criteria as in the initial meta-analyses (an uncorrected voxel-level threshold of p < 0.001, and an FWE-corrected cluster-level threshold of p < 0.05 with 5,000 permutations) to examine significant co-activated patterns (Gan et al., 2022).

1.3. **Preprocessing of gene expression data**

The full microarray gene expression dataset comprising more than 20,000 genes from 3,702 brain tissue samples covering the whole brain (cortical regions, subcortical regions, cerebellum, and brainstem) and the structural imaging (T1) data from six healthy adult doners were downloaded from the official website of AHBA (http://www.brain-map.org), and then preprocessed with reference to the standard processing pipeline and processing flow in the pioneering transcriptomic study of OT pathway genes (Arnatkevic̆iūtė et al., 2019; Hawrylycz et al., 2012; Quintana et al., 2019). (1) Probe-to-gene re-annotation: a list of all available 60 bp length AHBA probe sequences (n = 58,692) provided by Arnatkevic̆iūtė et al. (2019) was entered in Re-Annotator to update the probe-to-gene re-annotation (Arloth et al., 2015), with the latest available gene information from National Center for Biotechnology Information (NCBI; https://ftp.ncbi.nih.gov/gene/DATA/GENE_INFO/Mammalia/) and genome assembly hg38 (https://hgdownload.cse.ucsc.edu/goldenpath/hg38/bigZips/) as references. Finally, 46,331 probes were uniquely annotated to a gene, with 1,363 probes were re-annotated to new genes, 2,607 probes that were not previously assigned to any gene in the AHBA could be annotated now, and six probes were not given ID in AHBA. (2) Gene filter and selection: in keeping with a previous study (Quintana et al., 2019), whole-brain expression values for three critical genes in the OT pathway (CD38, OXT, and OXTR) and 17 genes that were crucial to vasopressinergic, acetylcholinergic, dopaminergic, opioidergic systems interaction with the oxytocinergic system were further extracted. Specifically, vasopressinergic gene set included arginine vasopressin receptor 1A (AVPR1A) and 1B (AVPR1B); acetylcholinergic gene set included cholinergic receptor muscarinic 1 (CHRM1), muscarinic 2 (CHRM2), muscarinic 3 (CHRM3), muscarinic 4 (CHRM4), and muscarinic 5 (CHRM5); dopaminergic gene set included dopamine receptor D1 (DRD1), D2 (DRD2), D3 (DRD3), D4 (DRD4), D5 (DRD5), catechol-O-methyltransferase (COMT), and solute carrier family 6 member 3 (SLC6A3, dopamine transporter); opioidergic gene included opioid receptor delta 1 (OPRD1), opioid receptor kappa 1(OPRK1), opioid receptor mu 1 (OPRM1). Of note, following the intensity-based filtering step, the AVPR1A and AVPR1B would have been excluded because they could not exceed the background noise in more than 50% of the samples. Moreover, the original probe (A_23_P252892) for detecting the gene expression of DRD5 also pointed to DRD5P2 after the latest gene re-annotation, which was considered to be a pseudogene with high sequence similarity to DRD5 but lacking in complete coding ability. Considering the importance of their roles, however, we still retained them and performed subsequent exploratory analyses with them. (3) Probe selection: in terms of those genes for which multiple mRNA probes were available, the probe (see Table S1 for a complete list of selected probes for candidate genes) with the highest differential stability value was selected (Hawrylycz et al., 2015). (4) Mapping samples to brain regions: based on the MNI coordinates and voxel coordinates of each tissue sample provided by AHBA, the expression data of the candidate genes for each sample in each donor were mapped to the corresponding location in native image space. To reconstruct missing voxel data, we annotated cerebral boundaries using the nearest neighboring sample expression values based on spatial proximity to each boundary coordinate, then performed Delaunay triangulation to partition the inter-sample space into non-overlapping simplices (Quintana et al., 2019). Finally, we applied linear interpolation within each simplex to estimate expression values across the volumetric space with the value of the vertex as a reference, resulting in a continuous volumetric reconstruction. The generated brain map for each donor was registered to MNI space with linear and nonlinear normalization in Advanced Normalization Tools (ANTs; https://github.com/ANTsX/ANTs/releases), and then we created a composite brain map for each gene for subsequent atlas parcellation, which was obtained by averaging the brain maps from the six donors. Individual and composite brain maps were further segmented into 246 brain regions based on the Brainnetome atlas (https://atlas.brainnetome.org/download.html), which were utilized for subsequent one-sample t-tests and correlation analyses (Fan et al., 2016). However, data from the right hemisphere for four out of six donors was not available, hence only left hemispherical data from six donors were utilized, leading to a matrix of 6 donors × 123 brain regions × 20 genes (Xu et al., 2024).

2. **Results**

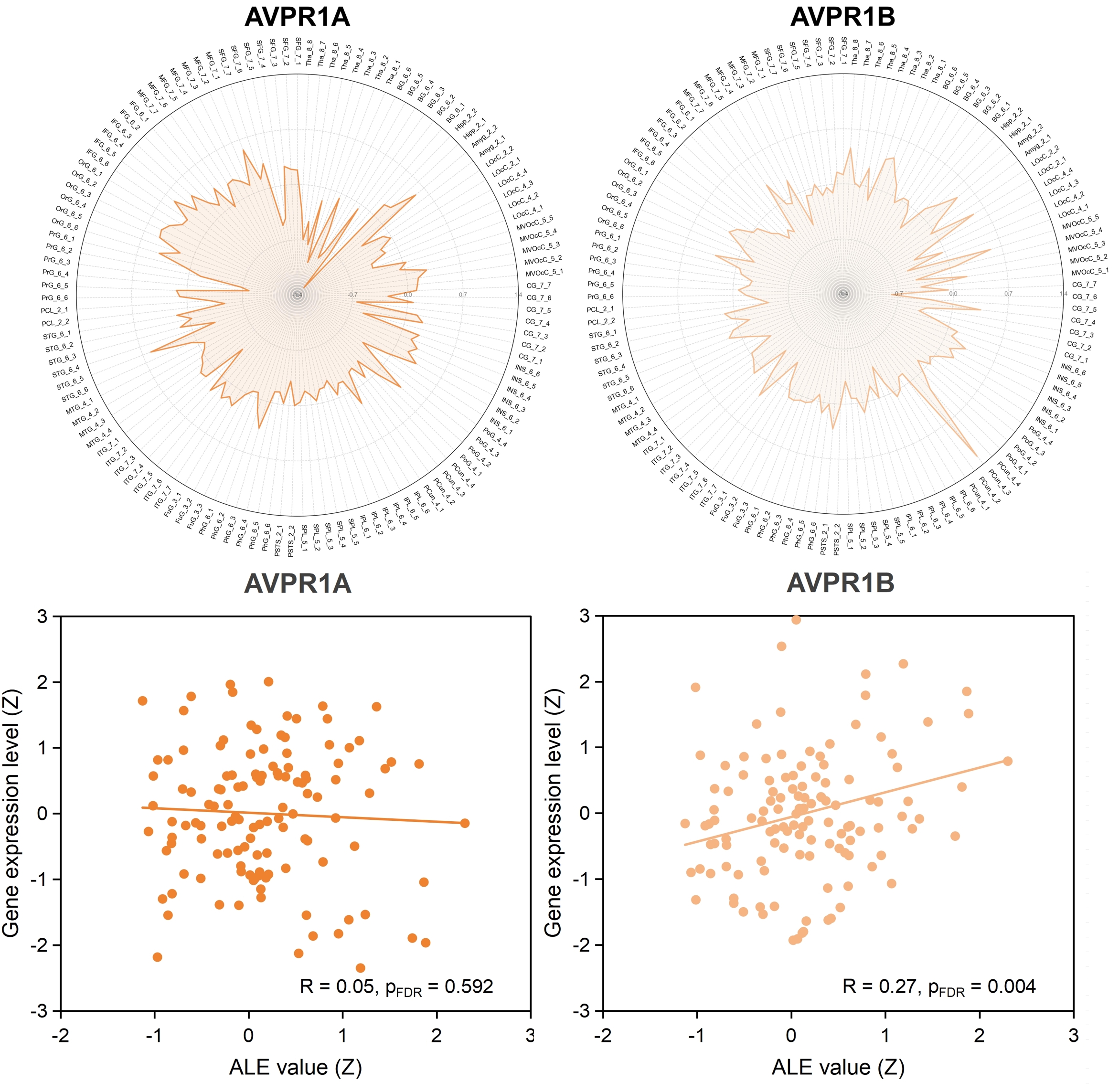

Fig. S1. The expression level of vasopressinergic genes in the whole brain and its correlation with neural effect of OT. The values in the radar maps represent effect sizes (Hedge’s g), indicating the differences in gene expression levels in each brain region compared to the average level of the whole brain. The brain region with significance symbol “*” denotes that the difference in gene expression levels in that region compared to the average level in the whole brain remains significant after FDR correction at 0.05 level. Scatter plots illustrate the Pearson spatial correlation between gene expression levels and ALE values derived from the general and non-directional effect of OT. Abbreviations for brain regions: Amyg = amygdala, BG = basal ganglia, CG = cingulate gyrus, FuG = fusiform gyrus, Hipp = hippocampus, IFG = inferior frontal gyrus, INS = insular gyrus, IPL = inferior parietal lobule, ITG = inferior temporal gyrus, LOcC = lateral occipital cortex, MFG = middle frontal gyrus, MTG = middle temporal gyrus, MVOcC = medioventral occipital cortex, OrG = orbital gyrus, PCL = paracentral lobule, Pcun = precuneus, PhG = parahippocampal gyrus, PoG = postcentral gyrus, PrG = precentral gyrus, pSTS = posterior superior temporal sulcus, STG = superior temporal gyrus, SFG = superior frontal gyrus, SPL = superior parietal lobule, Tha = thalamus; abbreviations for genes: AVPR1A = arginine vasopressin receptor 1A, AVPR1B = arginine vasopressin receptor 1B; other abbreviations: ALE = activation likelihood estimation, FDR = false discovery rate.

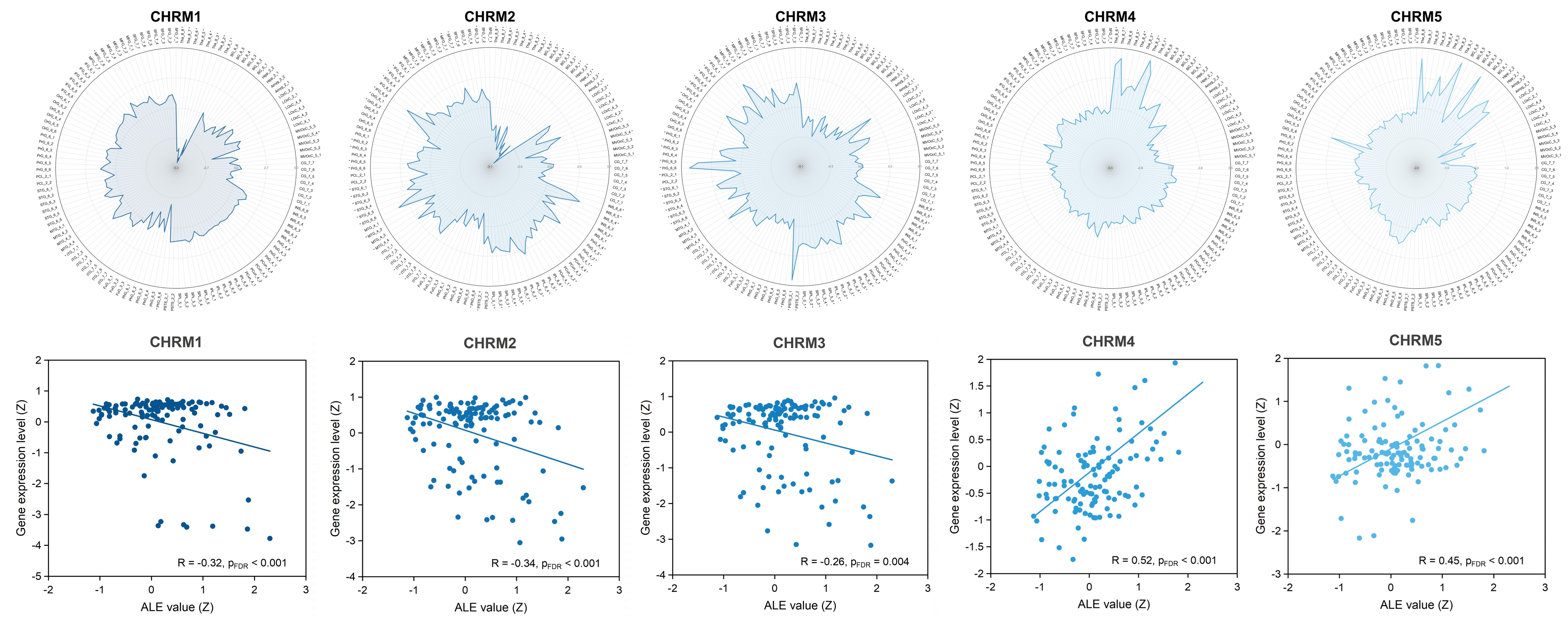

Fig. S2. The expression level of acetylcholinergic gene in the whole brain and its correlation with neural effect of OT. The values in the radar maps represent effect sizes (Hedge’s g), indicating the differences in gene expression levels in each brain region compared to the average level of the whole brain. The brain region with significance symbol “*” denotes that the difference in gene expression levels in that region compared to the average level in the whole brain remains significant after FDR correction at 0.05 level. Scatter plots illustrate the Pearson spatial correlation between gene expression levels and ALE values derived from the general and non-directional effect of OT. Abbreviations for brain regions: Amyg = amygdala, BG = basal ganglia, CG = cingulate gyrus, FuG = fusiform gyrus, Hipp = hippocampus, IFG = inferior frontal gyrus, INS = insular gyrus, IPL = inferior parietal lobule, ITG = inferior temporal gyrus, LOcC = lateral occipital cortex, MFG = middle frontal gyrus, MTG = middle temporal gyrus, MVOcC = medioventral occipital cortex, OrG = orbital gyrus, PCL = paracentral lobule, Pcun = precuneus, PhG = parahippocampal gyrus, PoG = postcentral gyrus, PrG = precentral gyrus, pSTS = posterior superior temporal sulcus, STG = superior temporal gyrus, SFG = superior frontal gyrus, SPL = superior parietal lobule, Tha = thalamus; abbreviations for genes: CHRM1 = cholinergic receptor muscarinic 1, CHRM2 = cholinergic receptor muscarinic 2, CHRM3 = cholinergic receptor muscarinic 3, CHRM4 = cholinergic receptor muscarinic 4, CHRM5 = cholinergic receptor muscarinic 5; other abbreviations: ALE = activation likelihood estimation, FDR = false discovery rate.

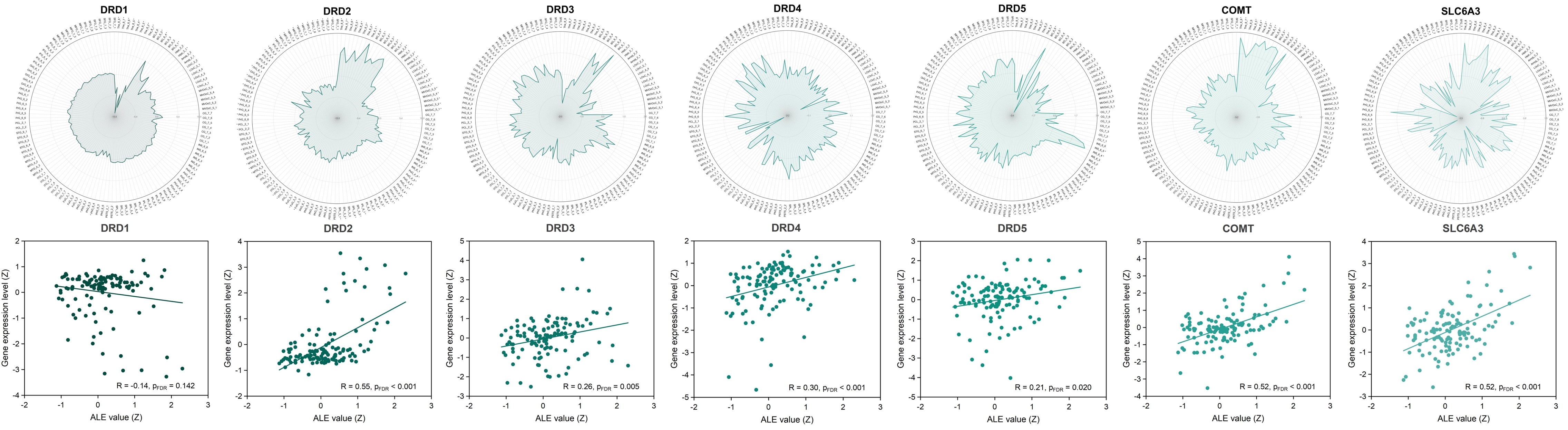

Fig. S3. The expression level of dopaminergic gene in the whole brain and its correlation with neural effect of OT. The values in the radar maps represent effect sizes (Hedge’s g), indicating the differences in gene expression levels in each brain region compared to the average level of the whole brain. The brain region with significance symbol “*” denotes that the difference in gene expression levels in that region compared to the average level in the whole brain remains significant after FDR correction at 0.05 level. Scatter plots illustrate the Pearson spatial correlation between gene expression levels and ALE values derived from the general and non-directional effect of OT. Abbreviations for brain regions: Amyg = amygdala, BG = basal ganglia, CG = cingulate gyrus, FuG = fusiform gyrus, Hipp = hippocampus, IFG = inferior frontal gyrus, INS = insular gyrus, IPL = inferior parietal lobule, ITG = inferior temporal gyrus, LOcC = lateral occipital cortex, MFG = middle frontal gyrus, MTG = middle temporal gyrus, MVOcC = medioventral occipital cortex, OrG = orbital gyrus, PCL = paracentral lobule, Pcun = precuneus, PhG = parahippocampal gyrus, PoG = postcentral gyrus, PrG = precentral gyrus, pSTS = posterior superior temporal sulcus, STG = superior temporal gyrus, SFG = superior frontal gyrus, SPL = superior parietal lobule, Tha = thalamus; abbreviations for genes: DRD1 = dopamine receptor D1, DRD2 = dopamine receptor D2, DRD3 = dopamine receptor D3, DRD4 = dopamine receptor D4, DRD5 = dopamine receptor D5, COMT = catechol-O-methyltransferase, SLC6A3 = solute carrier family 6 member 3; other abbreviations: ALE = activation likelihood estimation, FDR = false discovery rate.

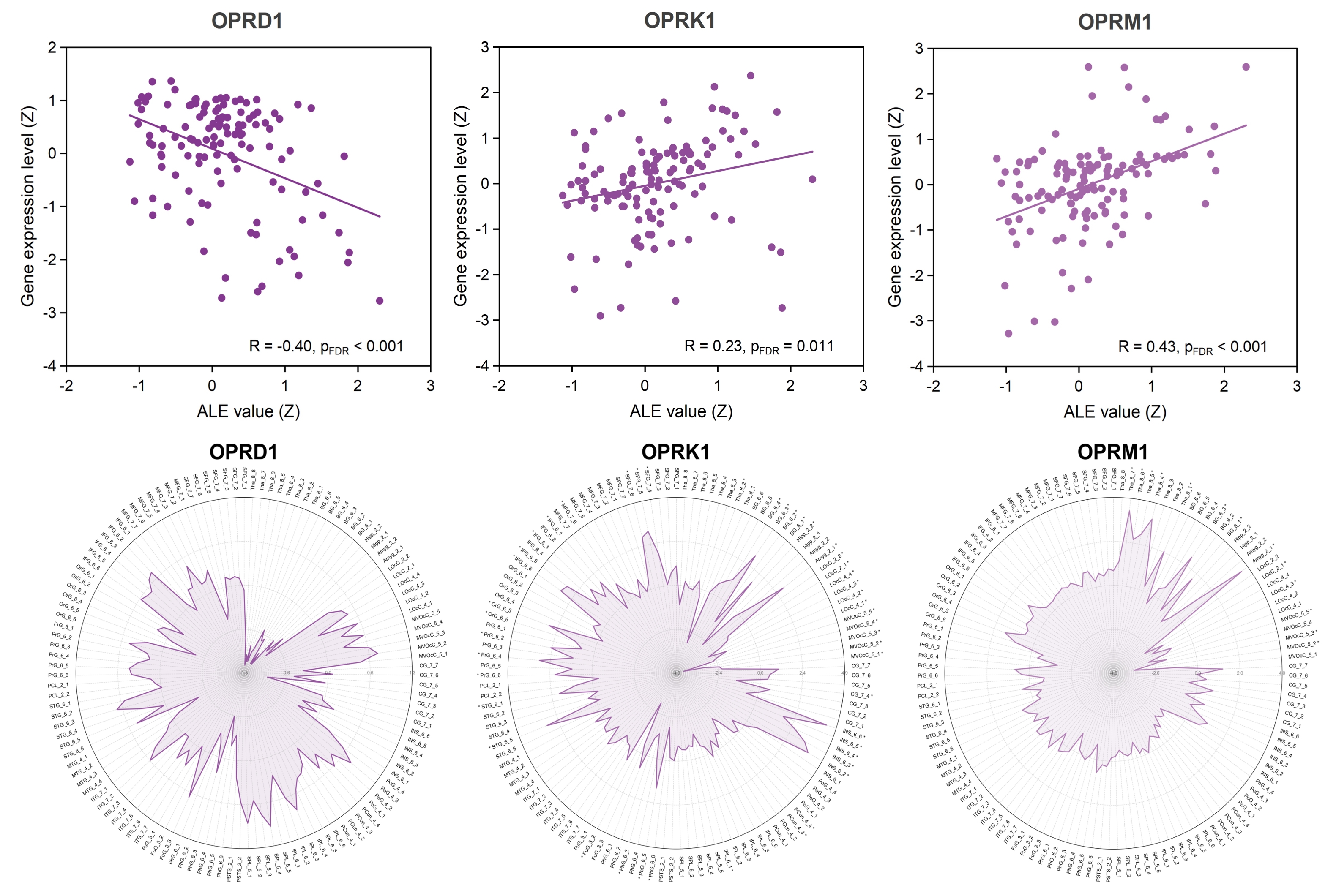

Fig. S4. The expression level of opioidergic gene in the whole brain and its correlation with neural effect of OT. The values in the radar maps represent effect sizes (Hedge’s g), indicating the differences in gene expression levels in each brain region compared to the average level of the whole brain. A brain region with significance symbol “*” signifies that the difference in gene expression levels in that region compared to the average level in the whole brain remains significant after FDR correction at 0.05 level. Scatter plots illustrate the Pearson spatial correlation between gene expression levels and ALE values derived from the general and non-directional effect of OT. Abbreviations for brain regions: Amyg = amygdala, BG = basal ganglia, CG = cingulate gyrus, FuG = fusiform gyrus, Hipp = hippocampus, IFG = inferior frontal gyrus, INS = insular gyrus, IPL = inferior parietal lobule, ITG = inferior temporal gyrus, LOcC = lateral occipital cortex, MFG = middle frontal gyrus, MTG = middle temporal gyrus, MVOcC = medioventral occipital cortex, OrG = orbital gyrus, PCL = paracentral lobule, Pcun = precuneus, PhG = parahippocampal gyrus, PoG = postcentral gyrus, PrG = precentral gyrus, pSTS = posterior superior temporal sulcus, STG = superior temporal gyrus, SFG = superior frontal gyrus, SPL = superior parietal lobule, Tha = thalamus; abbreviations for genes: OPRD1 = opioid receptor delta 1, OPRK1 = opioid receptor kappa 1, OPRM1 = opioid receptor mu 1; other abbreviations: ALE = activation likelihood estimation, FDR = false discovery rate.

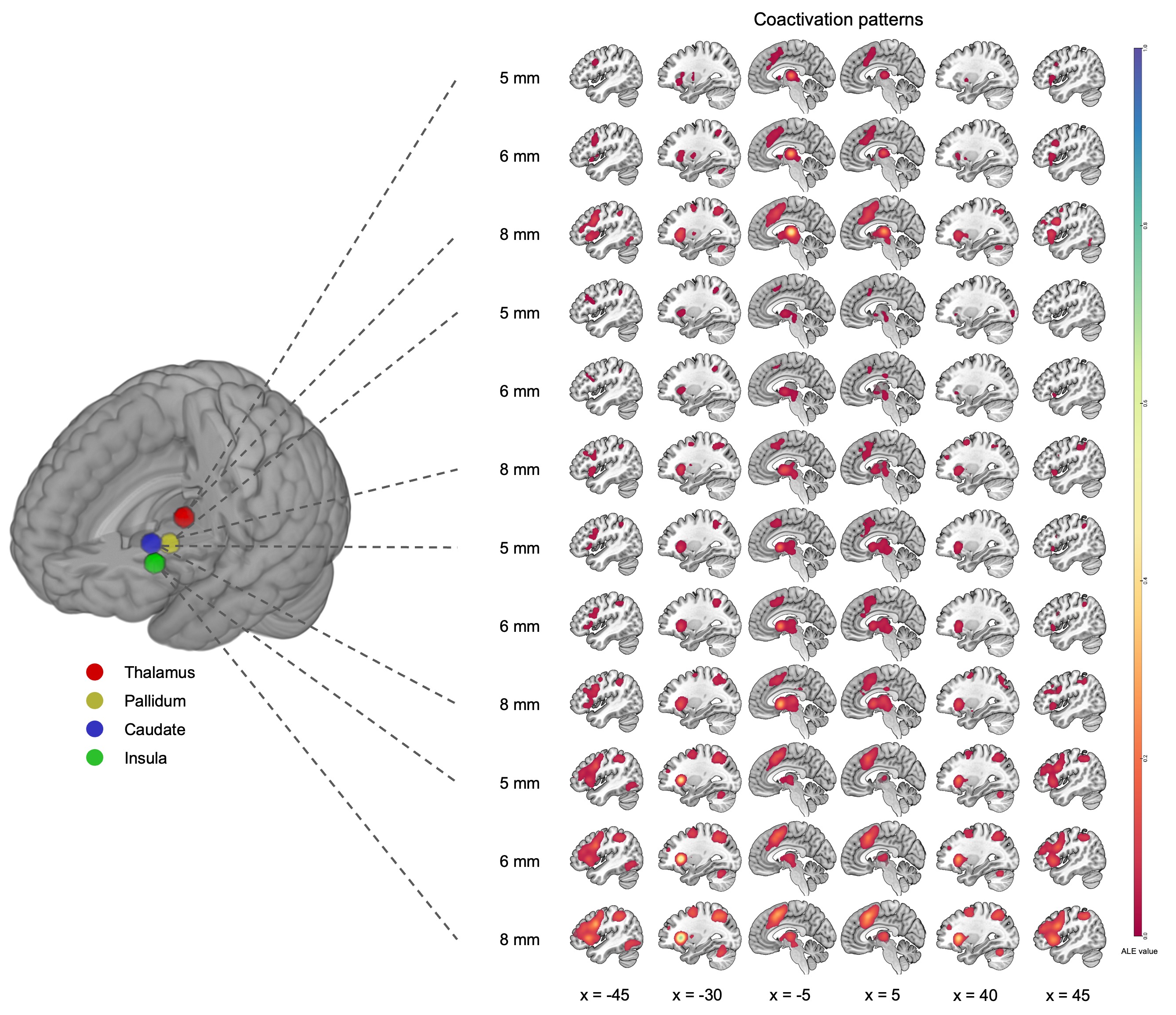

Fig. S5. The coactivation patterns for seed regions with different radii identified from MACM-B analysis. An uncorrected voxel-level threshold of p < 0.001 and an FWE-corrected cluster-level threshold of p < 0.05 are adopted to generate above results.

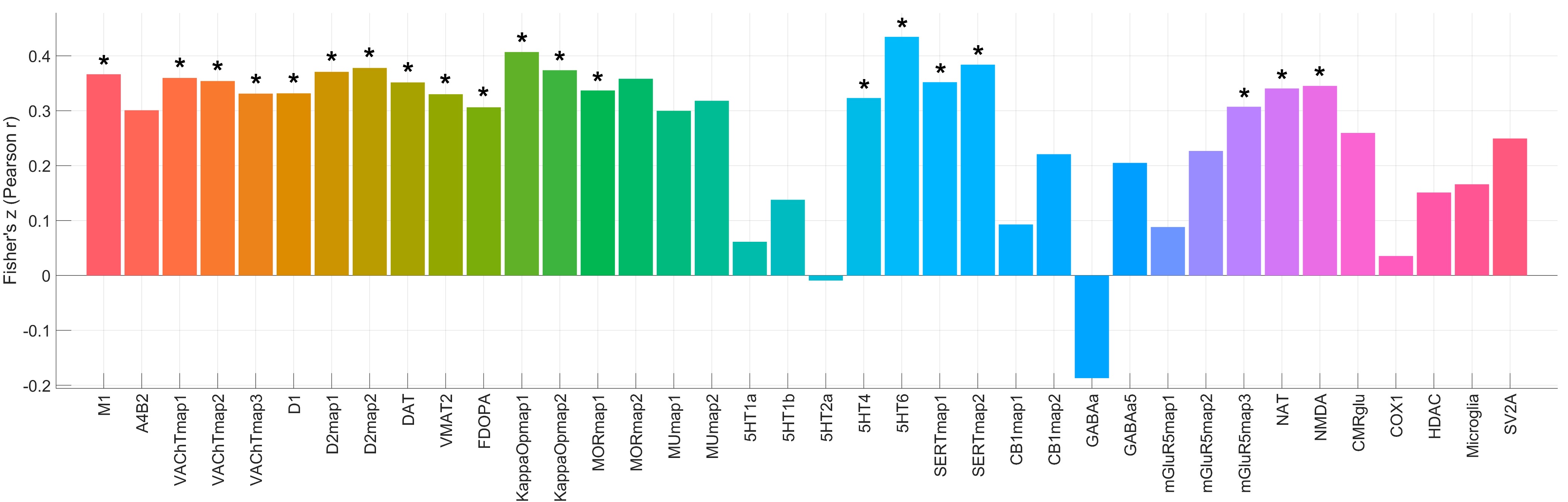

Fig. S6. The spatial correlations between vivo nuclear imaging-derived molecular maps and the ALE map for general and non-directional OT effect from meta-analysis. All results are corrected for spatial auto-correlation, and “*” indicates that the correlation coefficient remains significant after FDR correction at 0.05 level.

Table S1. Probes selected for candidate genes

| **Probe ID** | **Probe name** | **Gene ID** | **Gene symbol** |
| --- | --- | --- | --- |
| 1017111 | A_24_P397613 | 4988 | OPRM1 |
| 1021758 | A_32_P33576 | 4986 | OPRK1 |
| 1022735 | CUST_1385_PI417557136 | 1812 | DRD1 |
| 1023697 | CUST_314_PI417557136 | 1131 | CHRM3 |
| 1024432 | CUST_10874_PI416261804 | 1132 | CHRM4 |
| 1025406 | A_23_P57733 | 1814 | DRD3 |
| 1028943 | A_23_P11752 | 4985 | OPRD1 |
| 1028970 | CUST_1019_PI417557136 | 1312 | COMT |
| 1029133 | A_23_P145606 | 1129 | CHRM2 |
| 1030267 | CUST_240_PI416408490 | 1813 | DRD2 |
| 1051520 | CUST_15799_PI416261804 | 6531 | SLC6A3 |
| 1053417 | A_23_P132619 | 5021 | OXTR |
| 1053419 | A_24_P382579 | 5020 | OXT |
| 1057496 | A_23_P252892 | 1816 | DRD5 |
| 1057541 | A_23_P150162 | 1815 | DRD4 |
| 1058213 | A_23_P117873 | 1133 | CHRM5 |
| 1058221 | CUST_161_PI416408490 | 1128 | CHRM1 |
| 1058446 | CUST_14657_PI416261804 | 952 | CD38 |
| 1058916 | CUST_103_PI416408490 | 553 | AVPR1B |
| 1058917 | A_23_P25246 | 552 | AVPR1A |

Table S2. Vivo nuclear imaging-derived molecular maps used in the analysis

| **Category** | **PET/SPECT map name** | **Measured content** | **File name from the JuSpace toolbox** | **Data source** |
| --- | --- | --- | --- | --- |
| Hypothetical analysis | M1 | muscarinic acetylcholine receptor M1 | M1_lsn3172176_HC24_naganawa2020.nii | Naganawa et al., 2021 |
|  | A4B2 | nicotinic acetylcholine receptor α4β2 | A4B2_flubatine_HC30_hillmer2016.nii | Hillmer et al., 2016 |
|  | VAChTmap1 | acetylcholine transporter | VAChT_feobv_hc18_aghourian.nii | Aghourian et al., 2017 |
|  | VAChTmap2 | acetylcholine transporter | VAChT_feobv_hc4_tuominen.nii | Hansen et al., 2022 |
|  | VAChTmap3 | acetylcholine transporter | VAChT_feobv_hc5_bedard.nii | Bedard et al., 2019 |
|  | D1 | dopamine receptor D1 | D1_SCH23390_c11.nii | Kaller et al., 2017 |
|  | D2map1 | dopamine receptor D2 | D2_RACLOPRIDE_c11.nii | Alakurtti et al., 2015 |
|  | D2map2 | dopamine receptor D2 | D2_fallypride_hc49_jaworska.nii | Jaworska et al., 2020 |
|  | DAT | dopamine transporter | DAT_DATSPECT.nii | Dukart et al., 2018 |
|  | VMAT2 | vesicular monoamine transporter | VMAT2_dtbz_HC76_larsen2020.nii | Larsen et al., 2020 |
|  | FDOPA | dopamine synthesis capacity | FDOPA_f18.nii | Gómez et al., 2018 |
|  | KappaOpmap1 | opioid receptor Kappa | KappaOp_LY2795050_hc10_ShokriKojori.nii | Shokri-Kojori et al., 2022 |
|  | KappaOpmap2 | opioid receptor Kappa | KOR_ly2795050_HC28_vijay2018.nii | Vijay et al., 2018 |
|  | MORmap1 | opioid receptor Mu | MOR_carfentanil_HC204_kantonen2020.nii | Kantonen et al., 2020 |
|  | MORmap2 | opioid receptor Mu | MOR_carfentanil_HC39_turtonen2021.nii | Turtonen et al., 2021 |
|  | MUmap1 | opioid receptor Mu | MU_CARFENTANIL_c11.nii | Kantonen et al., 2020 |
|  | MUmap2 | opioid receptor Mu | MU_carfentanil_hc39_turtonen.nii | Turtonen et al., 2021 |
| Exploratory analysis | 5HT1a | 5-hydroxytryptamine receptor 1A | 5HT1a_WAY_HC36.nii | Savli et al., 2012 |
|  | 5HT1b | 5-hydroxytryptamine receptor 1B | 5HT1b_P943_HC22.nii | Savli et al., 2012 |
|  | 5HT2a | 5-hydroxytryptamine receptor 2A | 5HT2a_ALT_HC19.nii | Savli et al., 2012 |
|  | 5HT4 | 5-hydroxytryptamine receptor 4 | 5HT4_sb20_hc59_beliveau.nii | Beliveau et al., 2017 |
|  | 5HT6 | 5-hydroxytryptamine receptor 6 | 5HT6_gsk215083_HC30_radhakrishnan2018.nii | Radhakrishnan et al., 2018 |
|  | SERTmap1 | serotonin transporter | SERT_DASB_HC30.nii | Savli et al., 2012 |
|  | SERTmap2 | serotonin transporter | SERT_MADAM_c11.nii | Fazio et al., 2016 |
|  | CB1map1 | cannabinoid receptor 1 | CB1_FMPEPd2_hc22_laurikainen.nii | Laurikainen et al., 2019 |
|  | CB1map2 | cannabinoid receptor 1 | CB1_omar_HC77_normandin2015.nii | Normandin et al., 2015 |
|  | GABAa | Gamma-Aminobutyric Acid receptor A | GABAa_FLUMAZENIL_c11.nii | Dukart et al., 2018 |
|  | GABAa5 | Gamma-Aminobutyric Acid receptor A subtype α5 | GABAa5_ro154513_HC10_lukow2022.nii | Lukow et al., 2022 |
|  | mGluR5map1 | glutamate receptor 5 | mGluR5_abp_hc22_rosaneto.nii | Hansen et al., 2022 |
|  | mGluR5map2 | glutamate receptor 5 | mGluR5_abp_hc28_dubois.nii | DuBois et al., 2016 |
|  | mGluR5map3 | glutamate receptor 5 | mGluR5_abp_hc73_smart.nii | Smart et al., 2019 |
|  | NAT | noradrenaline transporter | NAT_MRB_c11.nii | Hesse et al., 2017 |
|  | NMDA | N-methyl-D-aspartate receptor | NMDA_ge179_29hc_galovic2021.nii | Galovic et al., 2021 |
|  | CMRglu | cerebral metabolic rate of glucose | CMRglu_fdg_HC20_castrillon2023.nii | Castrillon et al., 2023 |
|  | COX1 | cyclooxygenase-1 | COX1_ps13_HC11_kim2020.nii | Kim et al., 2020 |
|  | HDAC | histone deacetylase | HDAC_martinostat_HC8_wey2016.nii | Wey et al., 2016 |
|  | Microglia | 18 kDa translocator protein | Microglia_TSPO_pbr28_HC6_lois2018.nii | Lois et al., 2018 |
|  | SV2A | synaptic vesicle glycoprotein 2A | SV2A_ucbj_HC10_finnema2016.nii | Finnema et al., 2018 |

Table S3. Characteristics of the included studies with reference.

| **Study ID** | **Sample size** | **Sex** | **Mean age** | **SD of age** | **Dosage (IU)** | **Administration route** | **Interval (min)** | **Paradigm** | **Multiple comparison correction** |
| --- | --- | --- | --- | --- | --- | --- | --- | --- | --- |
| Andari et al., 2016 | 20 | M&F | 26.37 | 8.45 | 24 | Intranasal | 48-53 | Social ball-tossing game | Corrected |
| Aoki et al., 2014 | 20 | M | 30.8 | 6 | 24 | Intranasal | 40 | First-order false belief task (Modified Sally-Anne Task) | Uncorrected |
| Bach et al., 2021 | 13 | M | 34.5 | 16.7 | 24 | Intranasal | 60 | Face matching task | Corrected |
| Baettig et al., 2020 | 27 | M | 33.5 for OT 31 for PL | 9.8 for OT 7 for PL | 24 | Intranasal | 45 | Emotional processing and empathy task | Corrected |
| Chen et al., 2016_Female | 98 | F | 20.5 | 1.3 | 24 | Intranasal | 42 | Prisoner dilemma task | Corrected |
| Chen et al., 2016_Male | 96 | M | 20.7 | 2.2 | 24 | Intranasal | 42 | Prisoner dilemma task | Corrected |
| Chen et al., 2017 | 29 | F | 20.5 | 1.4 | 24 | Intranasal | 42 | Prisoner dilemma task | Corrected |
| Chen et al., 2020a | 40 | M | 21.19 | 2.62 | 24 | Intranasal | 45 | Affective touch task | Corrected |
| Chen et al., 2020b | 46 | M | 21.22 | 2.77 | 24 | Intranasal | 45 | Massage task | Corrected |
| Cohen et al., 2017 | 19 | M | 26.05 | 3.51 | 24 | Intranasal | 45 | Modified comfortable interpersonal distance task | Corrected |
| Cohen et al., 2018 | 22 | M | 28.02 | 2.69 | 24 | Intranasal | 45 | Implicit personal distance task. | Corrected |
| De Coster et al., 2019 | 23 | M | 35.3 | 10.48 | 40 | Intranasal | 45 | False belief task | Corrected |
| Domes et al., 2007 | 13 | M | 25.7 | 2.9 | 24 | Intranasal | 45 | Implicit facial emotion recognition task | Uncorrected |
| Domes et al., 2010 | 16 | F | 24.2 | 2.5 | 24 | Intranasal | 45-60 | Facial emotion viewing task | Uncorrected |
| Domes et al., 2014_AD | 14 | M | 24 | 6.9 | 24 | Intranasal | 45 | Facial emotion recognition task | Uncorrected |
| Domes et al., 2014_HP | 14 | M | 23.6 | 5.4 | 24 | Intranasal | 45 | Facial emotion recognition task | Uncorrected |
| Eckstein et al., 2014 | 60 | M | 24.67 | 3.89 | 24 | Intranasal | 30 | Montreal imaging stress task | Corrected |
| Eckstein et al., 2015 | 62 | M | 24.61 | 4.28 | 24 | Intranasal | 30 | Fear extinction task | Corrected |
| Eckstein et al., 2016 | 97 | M | 24.45 | 4.02 | 24 | Intranasal | 30 | Fear conditioning task | Corrected |
| Feng et al., 2015a | 196 | M&F | NA | NA | 24 | Intranasal | NA | Prisoner dilemma task | Corrected |
| Feng et al., 2015b | 177 178 | M&F | 19.97 for male 20.21 for female | NA | 24 | Intranasal | NA | Prisoner dilemma task | Corrected |
| Gao et al., 2016 | 74 | M&F | 22.8 | 1.7 | 24 | Intranasal | 45 | First-impression task | Corrected |
| Gao et al., 2022 | 62 | F | 19.68 | 1.53 | 24 | Intranasal | 45 | Compliment task (First-impression task) | Corrected |
| Gozzi et al., 2017 | 21 | M | 26.57 | NA | 24 | Intranasal | 45 | Social feedback task | Uncorrected |
| Grimm et al., 2014 | 31 | M | 28.4 29.5 | 4.5 4.5 | 24 | Intranasal | 45 | Montreal imaging stress task | Uncorrected |
| Groppe et al., 2013 | 28 | F | 26.64 | 5.55 | 26 | Intranasal | 30 | Social incentive delay task | Uncorrected |
| Hansson et al., 2018 | 12 | M | 33 | 16 | 24 | Intranasal | 45 | Alcohol cue-reactivity task | Corrected |
| Holtfrerich et al., 2018 | 57 | F | 24.63 | 3.08 | 24 | Intranasal | 45 | Target detection task | Corrected |
| Hu et al., 2015 | 54 | NA | 19.8 | 1.49 | 24 | Intranasal | 45 | Reinforcement association learning task | Partially corrected |
| Hu et al., 2016 | 22 | M | 25.1 | 3.88 | 24 | Intranasal | 45 | Third-party help/punishment task | Uncorrected |
| Kanat et al., 2015a | 43 | M | 24.32 for OT group 23.9 for PL group | 3.43 for OT group 2.74 for PL group | 24 | Intranasal | 55 | Facial emotion detection task | Uncorrected |
| Kanat et al., 2015b | 46 | M | 23.64 | 2.81 | 24 | Intranasal | 45 | Facial emotion detection task | Uncorrected |
| Kou et al., 2021 | 75 | M | 21.71 for OT group 22 for PL group | 1.87 for OT group 1.93 for PL group | 24 | Oral | 45 | Implicit face emotion processing task | Corrected |
| Labuschagne et al., 2010 | 34 | M | 29.4 for GSAD 29.9 for HC | 9 for GSAD 10.2 for HC | 24 | Intranasal | 45 | Emotional face matching task | Uncorrected |
| Labuschagne et al., 2012_GSAD | 18 | M | 29.4 | 9 | 24 | Intranasal | 50 | Emotional face processing task | Uncorrected |
| Labuschagne et al., 2012_HP | 18 | M | 29.9 | 10.2 | 24 | Intranasal | 50 | Emotional face processing task | Uncorrected |
| Labuschagne et al., 2018 | 19 | M | 48.3 for HD CAG-expansion 42.9 for HC | 11.4 for HD CAG-expansion 10.2 for HC | 24 | Intranasal | 45 | Emotional face matching task | Corrected |
| Lambert et al., 2017 | 29 | F | 24 | 2.8 | 24 | Intranasal | 35 | Social dilemma game | Uncorrected |
| Lan et al., 2023 | 70 | F | 21.7 for OT group 21.2 for PL group | 2 for OT group 2.3 for PL group | 24 | Oral | 53.5 | Implicit face emotion processing task | Corrected |
| Li et al., 2017 | 15 | M | 32.8 | 4.7 | 24 | Intranasal | 51.29 | Emotional face viewing task | Corrected |
| Lieberz et al., 2020 | 79 | F | 24 | 4.3 | 6 & 12 &24 | Intranasal | 45 | Emotional face recognition task | Corrected |
| Lischke et al., 2012 | 14 | F | 23.79 | 2.32 | 24 | Intranasal | 45 | Emotional arousal rating task | Uncorrected |
| Liu et al., 2017_EXP2 | 50 | M | 22.7 | 2.53 | 24 | Intranasal | 45 | Self-referential memory task | Corrected |
| Liu et al., 2017_EXP3 | 52 | M | 21.9 | 2.55 | 24 | Intranasal | 45 | Self-referential memory task | Corrected |
| Liu et al., 2019 | 116 | M | 22.7 for PL-Prosocial 21.8 for OT-Prosocial 23 for PL-Individualist 22.5 for OT-Individualist | 2.61 for PL-Prosocial 2.38 for OT-Prosocial 2.29 for PL-Individualist 3.03 for OT-Individualist | 24 | Intranasal | 35 | Monetary outcome-pair evaluation task | Corrected |
| Ma et al., 2018 | 104 | M&F | 21 | 0.22 | 40 | Intranasal | 45 | Social sharing task | Corrected |
| Ma et al., 2020 | 51 | M&F | NA | NA | 24 | Intranasal | 45 | Implicit emotional task | Corrected |
| Ma et al., 2022 | 65 | F | NA | NA | 24 | Intranasal | 45 | Facial recognition task | Uncorrected |
| Mayer et al., 2021 | 21 | M | 22.91 | 4.72 | 18 or 24, depending on age | Intranasal | 45 | Physical pain viewing task | Corrected |
| Meier et al., 2023 | 22 | F | 20.25 | 1.33 | 24 | Intranasal | 59 | Empathy task | Corrected |
| Mickey et al., 2016 | 18 | M | 22 | 2 | 24 | Intranasal | 49 | Monetary incentive delay task | Uncorrected |
| Oliver et al., 2020 | 23 | M&F | 64.29 | 7.88 | 72 | Intranasal | 45 | Facial expression viewing and imitating task | Corrected |
| Pincus et al., 2010_DD | 8 8 | F F | 35.5 | 10.62 | 40 | Intranasal | 10 | Reading the mind in the eyes task | Corrected |
| Pincus et al., 2010_HP | 9 9 | M & F M & F | 36.4 | 11.4 | 40 | Intranasal | 10 | Reading the mind in the eyes task | Corrected |
| Plessow et al., 2018 | 10 | M | 31.4 | 1.8 | 24 | Intranasal | 60 | Food motivation task | Uncorrected |
| Preckel et al., 2014 | 48 | M | 24.6 | 4.56 | 24 | Intranasal | 45 | Moral dilemma task | Corrected |
| Radke et al., 2020 | 39 | F | 22.8 | 3.1 | 24 | Intranasal | 45-60 | Verbal interaction social threat task | Corrected |
| Riem et al., 2011 | 42 | F | 29.07 | 7.56 | 24 | Intranasal | 36 | Infant cry task | Corrected |
| Rilling et al., 2012 | 60 | M | 20.2 | NA | 24 | Intranasal | 42 | Prisoner dilemma task | Corrected |
| Sauer et al., 2012 | 55 | M | 25.2 for CC genotype 24.6 for CA/AA genotype | 2.76 for CC genotype 2.42 for CA/AA genotype | 25 | Intranasal | 30 44 | Emotional face/social scenes matching task  Gaze processing task | Uncorrected |
| Sauer et al., 2019 | 55 | M | 24.9 | 2.6 | 25 | Intranasal | 30 | Emotional face/social scenes matching task | Uncorrected |
| Scheele et al., 2014a | 40 | M | 25.75 | 3.82 | 24 | Intranasal | 30 | Social touch task | Corrected |
| Scheele et al., 2014b | 23 | M | 25.57 | 3.29 | 24 | Intranasal | 30 | Emotional face viewing task | Corrected |
| Scheele et al., 2015 | 39 21 | F | 24.38 | 3.26 | 24 | Intranasal | 30 | Modified face perception task | Corrected |
| Schmidt et al., 2020 | 27 | M | 22.48 | 4.68 | 40 | Intranasal | NA | Sally-Anne task | Uncorrected |
| Spengler et al., 2017 | 107 | M | 24.7 | 4.4 | 12 & 24 & 48 | Intranasal | 15 & 45 & 75 | Facial emotion recognition task | Corrected |
| Spetter et al., 2018 | 15 | M | 25.7 | 2.6 | 24 | Intranasal | 35 51 | Food picture viewing task Monetary incentive delay task | Corrected |
| Striepens et al., 2012 | 70 | M | 26 | 4 | 24 | Intranasal | 45 | Aversive social stimulus viewing task | Corrected |
| Striepens et al., 2016 | 31 | F | 25.35 | 4.37 | 24 | Intranasal | 45 | Food craving task | Corrected |
| Vogt et al., 2023 | 49 | M | 24.5 | 5.3 | 24 | Intranasal | 40 | Speech production task | Corrected |
| Watanabe et al., 2014 | 33 | M | 28.5 | 5.9 | 24 | Intranasal | 40 | Friend or foe judgment task | Corrected |
| Wigton et al., 2022 | 20 | M | 37.9 | 7.43 | 40 | Intranasal | 90 | Rewarded decision-making task | Corrected |
| Wittfoth-Schardt et al., 2012 | 19 | M | 39.3 | 6.2 | 24 | Intranasal | 30 | Child emotional face viewing task | Uncorrected |
| Xin et al., 2020 | 65 | M | 20.83 | 2.6 | 24 | Intranasal | 70 | Distancing reappraisal task | Corrected |
| Xu et al., 2019 | 74 | M | 21.36 | 0.24 | 24 | Intranasal | 45 | Modified cyberball task | Corrected |

Note: more detailed information is available in Supplementary Material III, and the information provided here is intended only to comply with the PRISMA 2020: citing all studies included in the meta-analysis. AD = Asperger’s disorder, DD = depression disorder, Exp = experiment, F = female, GSAD = generalized social anxiety disorder, HP = healthy population, Interval = interval between administration and task-based fMRI scanning (min), M = male, NA = not available, OT = oxytocin, PL = placebo, SD = standard deviation.

Table S4. Sample characteristics for ALE meta-analyses for the full sample and subgroups

| **Category** | **OT non-directional effect** | **OT enhancing effect** | **OT attenuating effect** |
| --- | --- | --- | --- |
| Full sample | 70 studies  75 experiments  900 foci  151 contrasts  2247 subjects | 42 studies  45 experiments  430 foci  75 contrasts  1235 subjects | 29 studies  31 experiments  260 foci  44 contrasts  787 subjects |
| Social processing subgroup | 57 studies  61 experiments  727 foci  121 contrasts  1885 subjects | 34 studies  37 experiments  341 foci  57 contrasts  1043 subjects | 23 studies  24 experiments  197 foci  36 contrasts  624 subjects |
| Emotional processing subgroup | 44 studies  47 experiments  573 foci  87 contrasts  1517 subjects | 24 studies  26 experiments  251 foci  37 contrasts  784 subjects | 19 studies  21 experiments  181 foci  31 contrasts  575 subjects |
| Positive valence subgroup | 20 studies  21 experiments  225 foci  32 contrasts  691 subjects | 11 studies  12 experiments  123 foci  15 contrasts  369 subjects | 9 studies  10 experiments  52 foci  10 contrasts  172 subjects |
| Negative valence subgroup | 26 studies  28 experiments  280 foci  46 contrasts  729 subjects | 14 studies  15 experiments  83 foci  17 contrasts  377 subjects | 13 studies  15 experiments  129 foci  20 contrasts  368 subjects |
| Healthy population subgroup | 58 studies  60 experiments  687 foci  124 contrasts  1954 subjects | 36 studies  36 experiments  333 foci  63 contrasts  1058 subjects | 24 studies  25 experiments  187 foci  36 contrasts  674 subjects |
| Patient population subgroup | 13 studies  13 experiments  171 foci  21 contrasts  240 subjects | 10 studies  10 experiments  98 foci  13 contrasts  195 subjects | Insufficient experiments |
| Female subgroup | 17 studies  17 experiments  138 foci  33 contrasts  477 subjects | 13 studies  13 experiments  106 foci  21 contrasts  347 subjects | Insufficient experiments |
| Male subgroup | 47 studies  50 experiments  663 foci  107 contrasts  1439 subjects | 27 studies  29 experiments  290 foci  50 contrasts  848 subjects | 22 studies  23 experiments  207 foci  34 contrasts  589 subjects |

Note: OT = oxytocin.

Table S5. The brain regions affected by OT from uncorrected ALE meta-analyses

| **OT effect type** | **Volume (mm^3^)** | **Side** | **Brain region** | **MNI coordinates (x, y, z)** | **ALE value** | **Z** | **P** |
| --- | --- | --- | --- | --- | --- | --- | --- |
| OT > PL | | | | | | | |
|  | 456 | Right | Anterior cingulate cortex, supracallosal | 6, 34, 26 | 0.021 | 4.126 | 0.0000185 |
|  | 344 | Left | Temporal pole: superior temporal gyrus | -50, 10, -20 | 0.021 | 4.085 | 0.0000221 |
|  | 312 | Right | Temporal pole: superior temporal gyrus | 56, 18, -6 | 0.021 | 4.106 | 0.0000201 |
|  | 304 | Left | Inferior frontal gyrus, opercular part | -52, 6, 10 | 0.019 | 3.856 | 0.0000577 |
|  | 272 | Right | Substantia nigra, pars reticulata | 14, -18, -14 | 0.019 | 3.898 | 0.0000486 |
|  | 272 | Left | Ventral anterior thalamus | -10, -8, 2 | 0.019 | 3.898 | 0.0000486 |
|  | 240 | Left | Superior temporal gyrus | -58, -18, 10 | 0.019 | 3.834 | 0.0000631 |
|  | 216 | Left | Inferior frontal gyrus, pars orbitalis | -36, 24, 4 | 0.019 | 3.809 | 0.0000698 |
|  | 208 | Right | Mediodorsal lateral parvocellular thalamus | 6, -14, 6 | 0.018 | 3.732 | 0.0000949 |
|  | 184 | Left | Mediodorsal medial magnocellular thalamus | -6, -16, 6 | 0.018 | 3.758 | 0.0000858 |
|  | 184 | Right | Posterior cingulate gyrus | 12, -40, 26 | 0.018 | 3.750 | 0.0000884 |
|  | 168 | Right | Supplementary motor area | 12, 10, 56 | 0.019 | 3.789 | 0.0000757 |
|  | 120 | Left | Supramarginal gyrus | -48, -38, 28 | 0.017 | 3.580 | 0.0001720 |
|  | 112 | Left | Superior temporal gyrus | -52, -8, -2 | 0.017 | 3.614 | 0.0001509 |
|  | 104 | Right | Caudate | 14, 16, 0 | 0.017 | 3.614 | 0.0001509 |
| PL > OT | | | | | | | |
|  | 256 | Left | Anterior cingulate & paracingulate gyri | -8, 50, -2 | 0.016 | 3.961 | 0.0000372 |
|  | 216 | Left | Putamen | -22, -2, 10 | 0.014 | 3.553 | 0.0001903 |
|  |  | Left | Pallidum | -16, 2, 4 | 0.014 | 3.484 | 0.0002472 |
|  | 160 | Left | Middle frontal gyrus | -52, 18, 40 | 0.015 | 3.643 | 0.0001350 |
|  | 120 | Left | Superior frontal gyrus, medial | 0, 48, 36 | 0.015 | 3.627 | 0.0001431 |
|  | 112 | Right | Middle temporal gyrus | 62, -6, -20 | 0.013 | 3.362 | 0.0003862 |

Note: ALE = activation likelihood estimation, MNI = Montreal neurological institute, OT = oxytocin, PL = placebo. An uncorrected voxel-level threshold of p < 0.001 is adopted to generate above results.

Table S6. The coactivation network potentially affected by OT revealed by MACM-B analyses

| **Seed region (radius, x, y, z)** | **Volume (mm^3^)** | **Side** | **Brain region** | **MNI coordinates (x, y, z)** | **ALE value** | **Z** | **P** |
| --- | --- | --- | --- | --- | --- | --- | --- |
| Thalamus (5, -4, -16, 6) | | | | | | | |
| 1 | 13896 | Left | Mediodorsal medial magnocellular thalamus | -4, -16, 6 | 0.233 | 21.097 | 0.00E+00 |
| 1 |  | Right | Caudate | 12, 8, 4 | 0.034 | 4.983 | 3.13E-07 |
| 1 |  | Right | Putamen | 24, 4, -4 | 0.034 | 4.967 | 3.40E-07 |
| 1 |  | Right | Ventral anterior thalamus | 14, -2, 4 | 0.027 | 4.176 | 1.48E-05 |
| 2 | 9520 | Right | Middle cingulate & paracingulate gyri | 2, 14, 42 | 0.052 | 6.948 | 1.86E-12 |
| 2 |  | Right | Middle cingulate & paracingulate gyri | 2, 18, 36 | 0.048 | 6.574 | 2.45E-11 |
| 2 |  | Right | Supplementary motor area | 2, 8, 56 | 0.048 | 6.538 | 3.12E-11 |
| 2 |  | Left | Anterior cingulate & paracingulate gyri | -4, 30, 22 | 0.030 | 4.519 | 3.11E-06 |
| 3 | 6704 | Left | Pallidum | -14, 8, -4 | 0.039 | 5.556 | 1.38E-08 |
| 3 |  | Left | Putamen | -20, -4, 12 | 0.030 | 4.510 | 3.24E-06 |
| 3 |  | Left | Pallidum | -22, 0, 2 | 0.029 | 4.431 | 4.69E-06 |
| 3 |  | Left | Pallidum | -24, -4, -2 | 0.028 | 4.289 | 8.99E-06 |
| 3 |  | Left | Insula | -34, -6, 12 | 0.028 | 4.247 | 1.08E-05 |
| 4 | 3896 | Right | Insula | 38, 22, -6 | 0.048 | 6.517 | 3.58E-11 |
| 5 | 3512 | Left | Precentral gyrus | -44, 6, 34 | 0.037 | 5.319 | 5.22E-08 |
| 6 | 1784 | Left | Insula | -30, 18, 8 | 0.031 | 4.614 | 1.97E-06 |
| 6 |  | Left | Insula | -30, 22, -8 | 0.029 | 4.460 | 4.11E-06 |
| 7 | 1224 | Right | Inferior frontal gyrus, opercular part | 50, 10, 28 | 0.041 | 5.824 | 2.88E-09 |
| Pallidum (5, -12, -2, -2) | | | | | | | |
| 1 | 17296 | Left | Pallidum | -12, -2, -2 | 0.152 | 16.496 | 0.00E+00 |
| 1 |  | Right | Pallidum | 14, 2, -2 | 0.042 | 6.475 | 4.74E-11 |
| 1 |  | Right | Caudate | 14, 4, 6 | 0.039 | 6.147 | 3.94E-10 |
| 1 |  | Right | Ventral lateral thalamus | 12, -16, -2 | 0.034 | 5.612 | 1.00E-08 |
| 1 |  | Left | Red nucleus | -4, -22, -12 | 0.027 | 4.687 | 1.39E-06 |
| 1 |  | Right | Ventral lateral thalamus | 12, -8, 10 | 0.025 | 4.393 | 5.58E-06 |
| 1 |  | Right | Red nucleus | 0, -22, -18 | 0.024 | 4.263 | 1.01E-05 |
| 2 | 2872 | Left | Inferior frontal gyrus, opercular part | -48, 22, 32 | 0.025 | 4.359 | 6.52E-06 |
| 2 |  | Left | Precentral gyrus | -52, 4, 26 | 0.024 | 4.298 | 8.63E-06 |
| 2 |  | Left | Inferior frontal gyrus, opercular part | -44, 12, 22 | 0.023 | 4.163 | 1.57E-05 |
| 2 |  | Left | Precentral gyrus | -48, 4, 36 | 0.019 | 3.605 | 1.56E-04 |
| 3 | 2424 | Left | Insula | -32, 18, 2 | 0.040 | 6.228 | 2.36E-10 |
| 4 | 2160 | Right | Supplementary motor area | 2, 12, 48 | 0.025 | 4.378 | 5.98E-06 |
| 4 |  | Right | Middle cingulate & paracingulate gyri | 8, 14, 38 | 0.024 | 4.218 | 1.23E-05 |
| 5 | 1872 | Left | Inferior parietal gyrus | -34, -50, 48 | 0.028 | 4.798 | 8.03E-07 |
| 6 | 1016 | Right | Insula | 36, 22, -2 | 0.024 | 4.313 | 8.06E-06 |
| 7 | 896 | Right | Inferior frontal gyrus, triangular part | 50, 26, 30 | 0.022 | 3.991 | 3.29E-05 |
| 7 |  | Right | Inferior frontal gyrus, triangular part | 52, 24, 22 | 0.021 | 3.915 | 4.53E-05 |
| 8 | 824 | Right | Middle occipital gyrus | 32, -90, 2 | 0.027 | 4.639 | 1.75E-06 |
| 8 |  | Right | Inferior occipital gyrus | 32, -90, -6 | 0.021 | 3.894 | 4.92E-05 |
| Caudate (5, -10, 8, 0) | | | | | | | |
| 1 | 52744 | Left | Pallidum | -10, 8, -2 | 0.382 | 28.880 | 0.00E+00 |
| 1 |  | Right | Caudate | 12, 8, -2 | 0.209 | 17.633 | 0.00E+00 |
| 1 |  | Right | Insula | 34, 22, 0 | 0.110 | 10.937 | 3.91E-28 |
| 1 |  | Left | Insula | -32, 22, -2 | 0.092 | 9.567 | 5.54E-22 |
| 1 |  | Left | Ventral lateral thalamus | -10, -16, 4 | 0.064 | 7.222 | 2.57E-13 |
| 1 |  | Right | Ventral lateral thalamus | 14, -10, 8 | 0.061 | 6.973 | 1.56E-12 |
| 1 |  | Left | Precentral gyrus | -44, 6, 32 | 0.053 | 6.167 | 3.49E-10 |
| 1 |  | Right | Red nucleus | 4, -26, -6 | 0.046 | 5.513 | 1.76E-08 |
| 1 |  | Left | Substantia nigra | -6, -14, -10 | 0.044 | 5.361 | 4.14E-08 |
| 1 |  | Left | Inferior frontal gyrus, opercular part | -46, 8, 20 | 0.040 | 4.935 | 4.02E-07 |
| 1 |  | Right | Amygdala | 18, -2, -14 | 0.038 | 4.702 | 1.29E-06 |
| 1 |  | Left | Inferior frontal gyrus, opercular part | -50, 14, 8 | 0.035 | 4.407 | 5.24E-06 |
| 1 |  | Left | Inferior frontal gyrus, triangular part | -40, 28, 28 | 0.028 | 3.572 | 1.77E-04 |
| 1 |  | Left | Thalamus | -20, -8, 18 | 0.027 | 3.397 | 3.41E-04 |
| 2 | 7928 | Right | Supplementary motor area | 0, 10, 48 | 0.065 | 7.262 | 1.92E-13 |
| 2 |  | Right | Middle cingulate & paracingulate gyri | 6, 18, 36 | 0.038 | 4.737 | 1.08E-06 |
| 2 |  | Right | Anterior cingulate & paracingulate gyri | 8, 26, 20 | 0.032 | 4.011 | 3.03E-05 |
| 3 | 4400 | Left | Inferior parietal gyrus | -42, -48, 44 | 0.041 | 4.995 | 2.94E-07 |
| 3 |  | Left | Inferior parietal gyrus | -32, -50, 50 | 0.039 | 4.859 | 5.90E-07 |
| 3 |  | Left | Inferior parietal gyrus | -32, -50, 40 | 0.035 | 4.326 | 7.58E-06 |
| 3 |  | Left | Inferior parietal gyrus | -26, -66, 44 | 0.033 | 4.191 | 1.39E-05 |
| 3 |  | Left | Superior parietal gyrus | -24, -56, 54 | 0.031 | 3.882 | 5.19E-05 |
| 4 | 1440 | Right | Inferior parietal gyrus | 42, -44, 42 | 0.041 | 5.030 | 2.45E-07 |
| 4 |  | Right | Superior parietal gyrus | 38, -52, 58 | 0.028 | 3.521 | 2.15E-04 |
| 5 | 872 | Right | Inferior frontal gyrus, opercular part | 50, 12, 24 | 0.040 | 4.931 | 4.08E-07 |
| Insula (5, -36, 22, 4) | | | | | | | |
| 1 | 43296 | Right | Insula | 34, 22, 2 | 0.312 | 21.180 | 0.00E+00 |
| 1 |  | Right | Inferior frontal gyrus, opercular part | 48, 8, 28 | 0.144 | 11.335 | 4.51E-30 |
| 1 |  | Right | Mediodorsal lateral parvocellular thalamus | 12, -14, 6 | 0.089 | 7.553 | 2.14E-14 |
| 1 |  | Right | Inferior frontal gyrus, triangular part | 48, 26, 24 | 0.088 | 7.457 | 4.45E-14 |
| 1 |  | Right | Middle frontal gyrus | 40, 40, 26 | 0.080 | 6.847 | 3.78E-12 |
| 1 |  | Right | Putamen | 22, 8, 4 | 0.079 | 6.794 | 5.47E-12 |
| 1 |  | Right | Precentral gyrus | 52, 2, 44 | 0.074 | 6.402 | 7.66E-11 |
| 1 |  | Right | Pallidum | 14, 6, 0 | 0.073 | 6.353 | 1.06E-10 |
| 1 |  | Right | Middle frontal gyrus | 42, 0, 54 | 0.060 | 5.236 | 8.21E-08 |
| 1 |  | Right | Superior frontal gyrus, dorsolateral | 30, 4, 58 | 0.050 | 4.407 | 5.24E-06 |
| 1 |  | Right | Insula | 38, 2, 2 | 0.049 | 4.257 | 1.04E-05 |
| 1 |  | Right | Middle frontal gyrus | 36, 48, 18 | 0.049 | 4.253 | 1.05E-05 |
| 1 |  | Right | Superior frontal gyrus, dorsolateral | 30, 4, 48 | 0.047 | 4.086 | 2.19E-05 |
| 1 |  | Right | Middle frontal gyrus | 40, 48, 10 | 0.046 | 4.012 | 3.01E-05 |
| 1 |  | Right | Red nucleus | 10, -20, -6 | 0.042 | 3.606 | 1.55E-04 |
| 2 | 37448 | Left | Insula | -34, 22, 2 | 0.617 | Inf | 0.00E+00 |
| 2 |  | Left | Precentral gyrus | -46, 6, 32 | 0.157 | 12.160 | 2.61E-34 |
| 2 |  | Left | Inferior frontal gyrus, opercular part | -52, 10, 20 | 0.115 | 9.403 | 2.68E-21 |
| 2 |  | Left | Precentral gyrus | -30, -2, 54 | 0.080 | 6.897 | 2.66E-12 |
| 2 |  | Left | Inferior frontal gyrus, opercular part | -52, 12, 0 | 0.074 | 6.394 | 8.11E-11 |
| 2 |  | Left | Inferior frontal gyrus, triangular part | -46, 32, 18 | 0.070 | 6.063 | 6.68E-10 |
| 2 |  | Left | Precentral gyrus | -46, -4, 50 | 0.067 | 5.861 | 2.30E-09 |
| 2 |  | Left | Middle frontal gyrus | -36, 46, 22 | 0.062 | 5.398 | 3.37E-08 |
| 2 |  | Left | Middle frontal gyrus | -38, 52, 8 | 0.059 | 5.215 | 9.21E-08 |
| 3 | 21496 | Right | Supplementary motor area | 2, 18, 48 | 0.179 | 13.493 | 8.94E-42 |
| 3 |  | Left | Anterior cingulate & paracingulate gyri | -4, 34, 24 | 0.042 | 3.614 | 1.51E-04 |
| 4 | 13336 | Left | Inferior parietal gyrus | -34, -50, 46 | 0.142 | 11.218 | 1.71E-29 |
| 5 | 9376 | Left | Ventral lateral thalamus | -10, -12, 4 | 0.092 | 7.768 | 4.00E-15 |
| 5 |  | Left | Putamen | -20, 4, 4 | 0.083 | 7.134 | 4.89E-13 |
| 6 | 8936 | Right | Inferior parietal gyrus | 40, -50, 46 | 0.111 | 9.141 | 3.11E-20 |
| 6 |  | Right | Inferior parietal gyrus | 32, -56, 50 | 0.089 | 7.551 | 2.16E-14 |
| 6 |  | Right | Inferior parietal gyrus | 46, -38, 48 | 0.080 | 6.865 | 3.34E-12 |
| 7 | 3832 | Left | Supramarginal gyrus | -60, -38, 26 | 0.058 | 5.067 | 2.02E-07 |
| 7 |  | Left | Superior temporal gyrus | -64, -30, 8 | 0.056 | 4.886 | 5.14E-07 |
| 7 |  | Left | Superior temporal gyrus | -52, -42, 12 | 0.050 | 4.427 | 4.77E-06 |
| 7 |  | Left | Superior temporal gyrus | -60, -20, 6 | 0.047 | 4.101 | 2.05E-05 |
| 8 | 3176 | Left | Inferior parietal gyrus | -44, -68, -8 | 0.067 | 5.877 | 2.10E-09 |
| 9 | 2256 | Right | Superior temporal gyrus | 64, -22, 8 | 0.058 | 5.126 | 1.48E-07 |
| 9 |  | Right | Superior temporal gyrus | 54, -32, 2 | 0.049 | 4.312 | 8.10E-06 |
| 10 | 2192 | Right | Lobule VI of cerebellar hemisphere | 30, -62, -28 | 0.087 | 7.381 | 7.87E-14 |
| 11 | 2056 | Left | Lobule VI of cerebellar hemisphere | -28, -62, -26 | 0.081 | 6.925 | 2.19E-12 |

Note: ALE = activation likelihood estimation, MNI = Montreal neurological institute. An uncorrected voxel-level threshold of p < 0.001 and an FWE-corrected cluster-level threshold of p < 0.05 are adopted to generate above result.

3. **Reference**

Aghourian, M., Legault-Denis, C., Soucy, J. P., Rosa-Neto, P., Gauthier, S., Kostikov, A., Gravel, P., & Bédard, M. A., 2017. Quantification of brain cholinergic denervation in Alzheimer’s disease using PET imaging with [18f]-FEOBV. Mol. Psychiatry 22(11), 1531-1538. <https://doi.org/10.1038/mp.2017.183>.

Alakurtti, K., Johansson, J. J., Joutsa, J., Laine, M., Bäckman, L., Nyberg, L., & Rinne, J. O., 2015. Long-term test–retest reliability of striatal and extrastriatal dopamine D2/3 receptor binding: study with [11c] raclopride and high-resolution PET. J. Cereb. Blood Flow Metab. 35(7), 1199-1205. <https://doi.org/10.1038/jcbfm.2015.53>.

Andari, E., Richard, N., Leboyer, M., & Sirigu, A., 2016. Adaptive coding of the value of social cues with oxytocin, an fMRI study in autism spectrum disorder. Cortex 76, 79-88. <https://doi.org/10.1016/j.cortex.2015.12.010>.

Aoki, Y., Yahata, N., Watanabe, T., Takano, Y., Kawakubo, Y., Kuwabara, H., Iwashiro, N., Natsubori, T., Inoue, H., Suga, M., Takao, H., Sasaki, H., Gonoi, W., Kunimatsu, A., Kasai, K., & Yamasue, H., 2014. Oxytocin improves behavioural and neural deficits in inferring others’ social emotions in autism. Brain 137(11), 3073-3086. <https://doi.org/10.1093/brain/awu231>.

Arloth, J., Bader, D. M., Röh, S., & Altmann, A., 2015. Re-annotator: annotation pipeline for microarray probe sequences. Plos One 10(10), e0139516. <https://doi.org/10.1371/journal.pone.0139516>.

Arnatkevic̆iūtė, A., Fulcher, B. D., & Fornito, A., 2019. A practical guide to linking brain-wide gene expression and neuroimaging data. Neuroimage 189, 353-367. <https://doi.org/10.1016/j.neuroimage.2019.01.011>.

Bach, P., Koopmann, A., Bumb, J. M., Zimmermann, S., Bühler, S., Reinhard, I., Witt, S. H., Rietschel, M., Vollstädt-Klein, S., & Kiefer, F., 2021. Oxytocin attenuates neural response to emotional faces in social drinkers: an fMRI study. Eur. Arch. Psychiatry Clin. Neurosci. 271(5), 873-882. <https://doi.org/10.1007/s00406-020-01115-0>.

Baettig, L., Baeumelt, A., Ernst, J., Boeker, H., Grimm, S., & Richter, A., 2020. The awareness of the scared - context dependent influence of oxytocin on brain function. Brain Imaging Behav. 14(6), 2073-2083. <https://doi.org/10.1007/s11682-019-00143-2>.

Bedard, M.-A., Aghourian, M., Legault-Denis, C., Postuma, R. B., Soucy, J.-P., Gagnon, J.-F., Pelletier, A., & Montplaisir, J., 2019. Brain cholinergic alterations in idiopathic REM sleep behaviour disorder: a PET imaging study with 18F-FEOBV. Sleep Med. 58, 35-41. <https://doi.org/10.1016/j.sleep.2018.12.020>.

Beliveau, V., Ganz, M., Feng, L., Ozenne, B., Højgaard, L., Fisher, P. M., Svarer, C., Greve, D. N., & Knudsen, G. M., 2017. A high-resolution in vivo atlas of the human brain's serotonin system. J. Neurosci. 37(1), 120-128. <https://doi.org/10.1523/jneurosci.2830-16.2016>.

Brett, M., Anton, J.-L., Valabregue, R., & Poline, J.-B., 2002. Region of interest analysis using the MarsBar toolbox for SPM 99. Neuroimage 16(2), S497.

Castrillon, G., Epp, S., Bose, A., Fraticelli, L., Hechler, A., Belenya, R., Ranft, A., Yakushev, I., Utz, L., Sundar, L., Rauschecker, J. P., Preibisch, C., Kurcyus, K., & Riedl, V., 2023. An energy costly architecture of neuromodulators for human brain evolution and cognition. Sci. Adv. 9(50), eadi7632. <https://doi.org/10.1126/sciadv.adi7632>.

Chen, X., Gautam, P., Haroon, E., & Rilling, J. K., 2017. Within vs. between-subject effects of intranasal oxytocin on the neural response to cooperative and non-cooperative social interactions. Psychoneuroendocrinology 78, 22-30. <https://doi.org/10.1016/j.psyneuen.2017.01.006>.

Chen, X., Hackett, P. D., DeMarco, A. C., Feng, C., Stair, S., Haroon, E., Ditzen, B., Pagnoni, G., & Rilling, J. K., 2016. Effects of oxytocin and vasopressin on the neural response to unreciprocated cooperation within brain regions involved in stress and anxiety in men and women. Brain Imaging Behav. 10(2), 581-593. <https://doi.org/10.1007/s11682-015-9411-7>.

Chen, Y., Becker, B., Zhang, Y., Cui, H., Du, J., Wernicke, J., Montag, C., Kendrick, K. M., & Yao, S., 2020a. Oxytocin increases the pleasantness of affective touch and orbitofrontal cortex activity independent of valence. Eur. Neuropsychopharmacol. 39, 99-110. <https://doi.org/10.1016/j.euroneuro.2020.08.003>.

Chen, Y., Li, Q., Zhang, Q., Kou, J., Zhang, Y., Cui, H., Wernicke, J., Montag, C., Becker, B., Kendrick, K. M., & Yao, S., 2020b. The effects of intranasal oxytocin on neural and behavioral responses to social touch in the form of massage. Front. Neurosci. 14, 589878. <https://doi.org/10.3389/fnins.2020.589878>.

Cohen, D., Perry, A., Gilam, G., Mayseless, N., Gonen, T., Hendler, T., & Shamay-Tsoory, S. G., 2017. The role of oxytocin in modulating interpersonal space: a pharmacological fMRI study. Psychoneuroendocrinology 76, 77-83. <https://doi.org/10.1016/j.psyneuen.2016.10.021>.

Cohen, D., Perry, A., Mayseless, N., Kleinmintz, O., & Shamay-Tsoory, S. G., 2018. The role of oxytocin in implicit personal space regulation: an fMRI study. Psychoneuroendocrinology 91, 206-215. <https://doi.org/10.1016/j.psyneuen.2018.02.036>.

De Coster, L., Lin, L., Mathalon, D. H., & Woolley, J. D., 2019. Neural and behavioral effects of oxytocin administration during theory of mind in schizophrenia and controls: a randomized control trial. Neuropsychopharmacology 44(11), 1925-1931. <https://doi.org/10.1038/s41386-019-0417-5>.

Domes, G., Heinrichs, M., Gläscher, J., Büchel, C., Braus, D. F., & Herpertz, S. C., 2007. Oxytocin attenuates amygdala responses to emotional faces regardless of valence. Biol. Psychiatry 62(10), 1187-1190. <https://doi.org/10.1016/j.biopsych.2007.03.025>.

Domes, G., Kumbier, E., Heinrichs, M., & Herpertz, S. C., 2014. Oxytocin promotes facial emotion recognition and amygdala reactivity in adults with Asperger syndrome. Neuropsychopharmacology 39(3), 698-706. <https://doi.org/10.1038/npp.2013.254>.

Domes, G., Lischke, A., Berger, C., Grossmann, A., Hauenstein, K., Heinrichs, M., & Herpertz, S. C., 2010. Effects of intranasal oxytocin on emotional face processing in women. Psychoneuroendocrinology 35(1), 83-93. <https://doi.org/10.1016/j.psyneuen.2009.06.016>.

DuBois, J. M., Rousset, O. G., Rowley, J., Porras-Betancourt, M., Reader, A. J., Labbe, A., Massarweh, G., Soucy, J.-P., Rosa-Neto, P., & Kobayashi, E., 2016. Characterization of age/sex and the regional distribution of mGluR5 availability in the healthy human brain measured by high-resolution [11c]ABP688 PET. Eur. J. Nucl. Med. Mol. Imaging 43(1), 152-162. <https://doi.org/10.1007/s00259-015-3167-6>.

Dukart, J., Holiga, Š., Chatham, C., Hawkins, P., Forsyth, A., McMillan, R., Myers, J., Lingford-Hughes, A. R., Nutt, D. J., Merlo-Pich, E., Risterucci, C., Boak, L., Umbricht, D., Schobel, S., Liu, T., Mehta, M. A., Zelaya, F. O., Williams, S. C., Brown, G., Paulus, M., Honey, G. D., Muthukumaraswamy, S., Hipp, J., Bertolino, A., & Sambataro, F., 2018. Cerebral blood flow predicts differential neurotransmitter activity. Sci. Rep. 8(1), 4074. <https://doi.org/10.1038/s41598-018-22444-0>.

Eckstein, M., Becker, B., Scheele, D., Scholz, C., Preckel, K., Schlaepfer, T. E., Grinevich, V., Kendrick, K. M., Maier, W., & Hurlemann, R., 2015. Oxytocin facilitates the extinction of conditioned fear in humans. Biol. Psychiatry 78(3), 194-202. <https://doi.org/10.1016/j.biopsych.2014.10.015>.

Eckstein, M., Scheele, D., Patin, A., Preckel, K., Becker, B., Walter, A., Domschke, K., Grinevich, V., Maier, W., & Hurlemann, R., 2016. Oxytocin facilitates pavlovian fear learning in males. Neuropsychopharmacology 41(4), 932-939. <https://doi.org/10.1038/npp.2015.245>.

Eckstein, M., Scheele, D., Weber, K., Stoffel-Wagner, B., Maier, W., & Hurlemann, R., 2014. Oxytocin facilitates the sensation of social stress. Hum. Brain Mapp. 35(9), 4741-4750. <https://doi.org/10.1002/hbm.22508>.

Fan, L., Li, H., Zhuo, J., Zhang, Y., Wang, J., Chen, L., Yang, Z., Chu, C., Xie, S., Laird, A. R., Fox, P. T., Eickhoff, S. B., Yu, C., & Jiang, T., 2016. The human brainnetome atlas: a new brain atlas based on connectional architecture. Cereb. Cortex 26(8), 3508-3526. <https://doi.org/10.1093/cercor/bhw157>.

Fascher, M., Nowaczynski, S., & Muehlhan, M., 2024. Substance use disorders are characterised by increased voxel-wise intrinsic measures in sensorimotor cortices: an ALE meta-analysis. Neurosci. Biobehav. Rev. 162, 105712. <https://doi.org/10.1016/j.neubiorev.2024.105712>.

Fazio, P., Schain, M., Varnäs, K., Halldin, C., Farde, L., & Varrone, A., 2016. Mapping the distribution of serotonin transporter in the human brainstem with high-resolution PET: Validation using postmortem autoradiography data. Neuroimage 133, 313-320. <https://doi.org/10.1016/j.neuroimage.2016.03.019>.

Feng, C., Hackett, P. D., DeMarco, A. C., Chen, X., Stair, S., Haroon, E., Ditzen, B., Pagnoni, G., & Rilling, J. K., 2015a. Oxytocin and vasopressin effects on the neural response to social cooperation are modulated by sex in humans. Brain Imaging Behav. 9(4), 754-764. <https://doi.org/10.1007/s11682-014-9333-9>.

Feng, C., Lori, A., Waldman, I. D., Binder, E. B., Haroon, E., & Rilling, J. K., 2015b. A common oxytocin receptor gene (OXTR) polymorphism modulates intranasal oxytocin effects on the neural response to social cooperation in humans. Genes, Brain and Behav. 14(7), 516-525. <https://doi.org/10.1111/gbb.12234>.

Finnema, S. J., Nabulsi, N. B., Mercier, J., Lin, S.-f., Chen, M.-K., Matuskey, D., Gallezot, J.-D., Henry, S., Hannestad, J., Huang, Y., & Carson, R. E., 2018. Kinetic evaluation and test–retest reproducibility of [11^c^]UCB-J, a novel radioligand for positron emission tomography imaging of synaptic vesicle glycoprotein 2A in humans. J. Cereb. Blood Flow Metab. 38(11), 2041-2052. <https://doi.org/10.1177/0271678X17724947>.

Fox, P. T., Lancaster, J. L., Laird, A. R., & Eickhoff, S. B., 2014. Meta-analysis in human neuroimaging: computational modeling of large-scale databases. Annu. Rev. Neurosci. 37(1), 409-434. <https://doi.org/10.1146/annurev-neuro-062012-170320>.

Galovic, M., Erlandsson, K., Fryer, T. D., Hong, Y. T., Manavaki, R., Sari, H., Chetcuti, S., Thomas, B. A., Fisher, M., Sephton, S., Canales, R., Russell, J. J., Sander, K., Årstad, E., Aigbirhio, F. I., Groves, A. M., Duncan, J. S., Thielemans, K., Hutton, B. F., Coles, J. P., & Koepp, M. J., 2021. Validation of a combined image derived input function and venous sampling approach for the quantification of [18f]GE-179 PET binding in the brain. Neuroimage 237, 118194. <https://doi.org/10.1016/j.neuroimage.2021.118194>.

Gan, X., Zhou, X., Li, J., Jiao, G., Jiang, X., Biswal, B., Yao, S., Klugah-Brown, B., & Becker, B., 2022. Common and distinct neurofunctional representations of core and social disgust in the brain: coordinate-based and network meta-analyses. Neurosci. Biobehav. Rev. 135, 104553. <https://doi.org/10.1016/j.neubiorev.2022.104553>.

Gao, S., Becker, B., Luo, L., Geng, Y., Zhao, W., Yin, Y., Hu, J., Gao, Z., Gong, Q., Hurlemann, R., Yao, D., & Kendrick, K. M., 2016. Oxytocin, the peptide that bonds the sexes also divides them. Proc. Natl. Acad. Sci. U.S.A. 113(27), 7650-7654. <https://doi.org/10.1073/pnas.1602620113>.

Gao, Z., Ma, X., Zhou, X., Xin, F., Gao, S., Kou, J., Becker, B., & Kendrick, K. M., 2022. Oxytocin reduces the attractiveness of silver-tongued men for women during mid-cycle. Front. Neurosci. 16, 760695. <https://doi.org/10.3389/fnins.2022.760695>.

Gómez, F. J. G., Huertas, I., Ramírez, J. A. L., & Solís, D. G., 2018. Elaboración de una plantilla de SPM para la normalización de imágenes de PET con 18F-DOPA. Imagen Diagnóstica 9(01), 23-25. <https://doi.org/10.33588/imagendiagnostica.901.2>.

Gozzi, M., Dashow, E. M., Thurm, A., Swedo, S. E., & Zink, C. F., 2017. Effects of oxytocin and vasopressin on preferential brain responses to negative social feedback. Neuropsychopharmacology 42(7), 1409-1419. <https://doi.org/10.1038/npp.2016.248>.

Grimm, S., Pestke, K., Feeser, M., Aust, S., Weigand, A., Wang, J., Wingenfeld, K., Pruessner, J. C., La Marca, R., Böker, H., & Bajbouj, M., 2014. Early life stress modulates oxytocin effects on limbic system during acute psychosocial stress. Soc. Cogn. Affect. Neurosci. 9(11), 1828-1835. <https://doi.org/10.1093/scan/nsu020>.

Groppe, S. E., Gossen, A., Rademacher, L., Hahn, A., Westphal, L., Gründer, G., & Spreckelmeyer, K. N., 2013. Oxytocin influences processing of socially relevant cues in the ventral tegmental area of the human brain. Biol. Psychiatry 74(3), 172-179. <https://doi.org/10.1016/j.biopsych.2012.12.023>.

Hansen, J. Y., Shafiei, G., Markello, R. D., Smart, K., Cox, S. M. L., Nørgaard, M., Beliveau, V., Wu, Y., Gallezot, J.-D., Aumont, É., Servaes, S., Scala, S. G., DuBois, J. M., Wainstein, G., Bezgin, G., Funck, T., Schmitz, T. W., Spreng, R. N., Galovic, M., Koepp, M. J., Duncan, J. S., Coles, J. P., Fryer, T. D., Aigbirhio, F. I., McGinnity, C. J., Hammers, A., Soucy, J.-P., Baillet, S., Guimond, S., Hietala, J., Bedard, M.-A., Leyton, M., Kobayashi, E., Rosa-Neto, P., Ganz, M., Knudsen, G. M., Palomero-Gallagher, N., Shine, J. M., Carson, R. E., Tuominen, L., Dagher, A., & Misic, B., 2022. Mapping neurotransmitter systems to the structural and functional organization of the human neocortex. Nat. Neurosci. 25(11), 1569-1581. <https://doi.org/10.1038/s41593-022-01186-3>.

Hansson, A. C., Koopmann, A., Uhrig, S., Bühler, S., Domi, E., Kiessling, E., Ciccocioppo, R., Froemke, R. C., Grinevich, V., Kiefer, F., Sommer, W. H., Vollstädt-Klein, S., & Spanagel, R., 2018. Oxytocin reduces alcohol cue-reactivity in alcohol-dependent rats and humans. Neuropsychopharmacology 43(6), 1235-1246. <https://doi.org/10.1038/npp.2017.257>.

Hawrylycz, M., Miller, J. A., Menon, V., Feng, D., Dolbeare, T., Guillozet-Bongaarts, A. L., Jegga, A. G., Aronow, B. J., Lee, C.-K., Bernard, A., Glasser, M. F., Dierker, D. L., Menche, J., Szafer, A., Collman, F., Grange, P., Berman, K. A., Mihalas, S., Yao, Z., Stewart, L., Barabási, A.-L., Schulkin, J., Phillips, J., Ng, L., Dang, C., Haynor, D. R., Jones, A., Van Essen, D. C., Koch, C., & Lein, E., 2015. Canonical genetic signatures of the adult human brain. Nat. Neurosci. 18(12), 1832-1844. <https://doi.org/10.1038/nn.4171>.

Hawrylycz, M. J., Lein, E. S., Guillozet-Bongaarts, A. L., Shen, E. H., Ng, L., Miller, J. A., van de Lagemaat, L. N., Smith, K. A., Ebbert, A., Riley, Z. L., Abajian, C., Beckmann, C. F., Bernard, A., Bertagnolli, D., Boe, A. F., Cartagena, P. M., Chakravarty, M. M., Chapin, M., Chong, J., Dalley, R. A., Daly, B. D., Dang, C., Datta, S., Dee, N., Dolbeare, T. A., Faber, V., Feng, D., Fowler, D. R., Goldy, J., Gregor, B. W., Haradon, Z., Haynor, D. R., Hohmann, J. G., Horvath, S., Howard, R. E., Jeromin, A., Jochim, J. M., Kinnunen, M., Lau, C., Lazarz, E. T., Lee, C., Lemon, T. A., Li, L., Li, Y., Morris, J. A., Overly, C. C., Parker, P. D., Parry, S. E., Reding, M., Royall, J. J., Schulkin, J., Sequeira, P. A., Slaughterbeck, C. R., Smith, S. C., Sodt, A. J., Sunkin, S. M., Swanson, B. E., Vawter, M. P., Williams, D., Wohnoutka, P., Zielke, H. R., Geschwind, D. H., Hof, P. R., Smith, S. M., Koch, C., Grant, S. G. N., & Jones, A. R., 2012. An anatomically comprehensive atlas of the adult human brain transcriptome. Nature 489(7416), 391-399. <https://doi.org/10.1038/nature11405>.

Hesse, S., Becker, G.-A., Rullmann, M., Bresch, A., Luthardt, J., Hankir, M. K., Zientek, F., Reißig, G., Patt, M., Arelin, K., Lobsien, D., Müller, U., Baldofski, S., Meyer, P. M., Blüher, M., Fasshauer, M., Fenske, W. K., Stumvoll, M., Hilbert, A., Ding, Y.-S., & Sabri, O., 2017. Central noradrenaline transporter availability in highly obese, non-depressed individuals. Eur. J. Nucl. Med. Mol. Imaging 44(6), 1056-1064. <https://doi.org/10.1007/s00259-016-3590-3>.

Hillmer, A. T., Esterlis, I., Gallezot, J. D., Bois, F., Zheng, M. Q., Nabulsi, N., Lin, S. F., Papke, R. L., Huang, Y., Sabri, O., Carson, R. E., & Cosgrove, K. P., 2016. Imaging of cerebral α4β2* nicotinic acetylcholine receptors with (−)-[18f]Flubatine PET: implementation of bolus plus constant infusion and sensitivity to acetylcholine in human brain. Neuroimage 141, 71-80. <https://doi.org/10.1016/j.neuroimage.2016.07.026>.

Holtfrerich, S. K. C., Pfister, R., El Gammal, A. T., Bellon, E., & Diekhof, E. K., 2018. Endogenous testosterone and exogenous oxytocin influence the response to baby schema in the female brain. Sci. Rep. 8(1), 7672. <https://doi.org/10.1038/s41598-018-26020-4>.

Hu, J., Qi, S., Becker, B., Luo, L., Gao, S., Gong, Q., Hurlemann, R., & Kendrick, K. M., 2015. Oxytocin selectively facilitates learning with social feedback and increases activity and functional connectivity in emotional memory and reward processing regions. Hum. Brain Mapp. 36(6), 2132-2146. <https://doi.org/10.1002/hbm.22760>.

Hu, Y., Scheele, D., Becker, B., Voos, G., David, B., Hurlemann, R., & Weber, B., 2016. The effect of oxytocin on third-party altruistic decisions in unfair situations: an fMRI study. Sci. Rep. 6(1), 20236. <https://doi.org/10.1038/srep20236>.

Jaworska, N., Cox, S. M. L., Tippler, M., Castellanos-Ryan, N., Benkelfat, C., Parent, S., Dagher, A., Vitaro, F., Boivin, M., Pihl, R. O., Côté, S. M., Tremblay, R. E., Séguin, J. R., & Leyton, M., 2020. Extra-striatal D2/3 receptor availability in youth at risk for addiction. Neuropsychopharmacology 45(9), 1498-1505. <https://doi.org/10.1038/s41386-020-0662-7>.

Kaller, S., Rullmann, M., Patt, M., Becker, G.-A., Luthardt, J., Girbardt, J., Meyer, P. M., Werner, P., Barthel, H., Bresch, A., Fritz, T. H., Hesse, S., & Sabri, O., 2017. Test–retest measurements of dopamine D1-type receptors using simultaneous PET/MRI imaging. Eur. J. Nucl. Med. Mol. Imaging 44(6), 1025-1032. <https://doi.org/10.1007/s00259-017-3645-0>.

Kanat, M., Heinrichs, M., Mader, I., van Elst, L. T., & Domes, G., 2015a. Oxytocin modulates amygdala reactivity to masked fearful eyes. Neuropsychopharmacology 40(11), 2632-2638. <https://doi.org/10.1038/npp.2015.111>.

Kanat, M., Heinrichs, M., Schwarzwald, R., & Domes, G., 2015b. Oxytocin attenuates neural reactivity to masked threat cues from the eyes. Neuropsychopharmacology 40(2), 287-295. <https://doi.org/10.1038/npp.2014.183>.

Kantonen, T., Karjalainen, T., Isojärvi, J., Nuutila, P., Tuisku, J., Rinne, J., Hietala, J., Kaasinen, V., Kalliokoski, K., Scheinin, H., Hirvonen, J., Vehtari, A., & Nummenmaa, L., 2020. Interindividual variability and lateralization of μ-opioid receptors in the human brain. Neuroimage 217, 116922. <https://doi.org/10.1016/j.neuroimage.2020.116922>.

Kim, M.-J., Lee, J.-H., Juarez Anaya, F., Hong, J., Miller, W., Telu, S., Singh, P., Cortes, M. Y., Henry, K., Tye, G. L., Frankland, M. P., Montero Santamaria, J. A., Liow, J.-S., Zoghbi, S. S., Fujita, M., Pike, V. W., & Innis, R. B., 2020. First-in-human evaluation of [11c]PS13, a novel PET radioligand, to quantify cyclooxygenase-1 in the brain. Eur. J. Nucl. Med. Mol. Imaging 47(13), 3143-3151. <https://doi.org/10.1007/s00259-020-04855-2>.

Kou, J., Lan, C., Zhang, Y., Wang, Q., Zhou, F., Zhao, Z., Montag, C., Yao, S., Becker, B., & Kendrick, K. M., 2021. In the nose or on the tongue? Contrasting motivational effects of oral and intranasal oxytocin on arousal and reward during social processing. Transl. Psychiatry 11(1), 94. <https://doi.org/10.1038/s41398-021-01241-w>.

Labuschagne, I., Phan, K. L., Wood, A., Angstadt, M., Chua, P., Heinrichs, M., Stout, J. C., & Nathan, P. J., 2010. Oxytocin attenuates amygdala reactivity to fear in generalized social anxiety disorder. Neuropsychopharmacology 35(12), 2403-2413. <https://doi.org/10.1038/npp.2010.123>.

Labuschagne, I., Phan, K. L., Wood, A., Angstadt, M., Chua, P., Heinrichs, M., Stout, J. C., & Nathan, P. J., 2012. Medial frontal hyperactivity to sad faces in generalized social anxiety disorder and modulation by oxytocin. Int. J. Neuropsychopharmacol. 15(7), 883-896. <https://doi.org/10.1017/s1461145711001489>.

Labuschagne, I., Poudel, G., Kordsachia, C., Wu, Q., Thomson, H., Georgiou-Karistianis, N., & Stout, J. C., 2018. Oxytocin selectively modulates brain processing of disgust in Huntington's disease gene carriers. Prog. Neuro-Psychopharmacol. Biol. Psychiatry 81, 11-16. <https://doi.org/10.1016/j.pnpbp.2017.09.023>.

Lambert, B., Declerck, C. H., Boone, C., & Parizel, P. M., 2017. A functional MRI study on how oxytocin affects decision making in social dilemmas: cooperate as long as it pays off, aggress only when you think you can win. Horm. Behav. 94, 145-152. <https://doi.org/10.1016/j.yhbeh.2017.06.011>.

Lan, C., Chen, Y., Zhang, Y., Kou, J., Huang, L., Xu, T., Yang, X., Xu, D., Yang, W., Kendrick, K. M., & Zhao, W., 2023. Oral oxytocin facilitates responses to emotional faces in reward and emotional-processing networks in females. Neuroendocrinology 113(9), 957-970. <https://doi.org/10.1159/000531064>.

Langner, R., & Camilleri, J. A., 2021. Meta-Analytic Connectivity Modelling (MACM): A tool for assessing region-specific functional connectivity patterns in task-constrained states. In V. A. Diwadkar & S. B. Eickhoff (Eds.), Brain network dysfunction in neuropsychiatric illness: methods, applications, and implications (pp. 93-104). Springer International Publishing. <https://doi.org/10.1007/978-3-030-59797-9_5>.

Larsen, B., Olafsson, V., Calabro, F., Laymon, C., Tervo-Clemmens, B., Campbell, E., Minhas, D., Montez, D., Price, J., & Luna, B., 2020. Maturation of the human striatal dopamine system revealed by PET and quantitative MRI. Nat. Commun. 11(1), 846. <https://doi.org/10.1038/s41467-020-14693-3>.

Laurikainen, H., Tuominen, L., Tikka, M., Merisaari, H., Armio, R.-L., Sormunen, E., Borgan, F., Veronese, M., Howes, O., Haaparanta-Solin, M., Solin, O., & Hietala, J., 2019. Sex difference in brain CB1 receptor availability in man. Neuroimage 184, 834-842. <https://doi.org/10.1016/j.neuroimage.2018.10.013>.

Li, T., Chen, X., Mascaro, J., Haroon, E., & Rilling, J. K., 2017. Intranasal oxytocin, but not vasopressin, augments neural responses to toddlers in human fathers. Horm. Behav. 93, 193-202. <https://doi.org/10.1016/j.yhbeh.2017.01.006>.

Lieberz, J., Scheele, D., Spengler, F. B., Matheisen, T., Schneider, L., Stoffel-Wagner, B., Kinfe, T. M., & Hurlemann, R., 2020. Kinetics of oxytocin effects on amygdala and striatal reactivity vary between women and men. Neuropsychopharmacology 45(7), 1134-1140. <https://doi.org/10.1038/s41386-019-0582-6>.

Lischke, A., Gamer, M., Berger, C., Grossmann, A., Hauenstein, K., Heinrichs, M., Herpertz, S. C., & Domes, G., 2012. Oxytocin increases amygdala reactivity to threatening scenes in females. Psychoneuroendocrinology 37(9), 1431-1438. <https://doi.org/10.1016/j.psyneuen.2012.01.011>.

Liu, Y., Li, S., Lin, W., Li, W., Yan, X., Wang, X., Pan, X., Rutledge, R. B., & Ma, Y., 2019. Oxytocin modulates social value representations in the amygdala. Nat. Neurosci. 22(4), 633-641. <https://doi.org/10.1038/s41593-019-0351-1>.

Liu, Y., Wu, B., Wang, X., Li, W., Zhang, T., Wu, X., & Han, S., 2017. Oxytocin effects on self-referential processing: behavioral and neuroimaging evidence. Soc. Cogn. Affect. Neurosci. 12(12), 1845-1858. <https://doi.org/10.1093/scan/nsx116>.

Lois, C., González, I., Izquierdo-García, D., Zürcher, N. R., Wilkens, P., Loggia, M. L., Hooker, J. M., & Rosas, H. D., 2018. Neuroinflammation in Huntington’s disease: new insights with 11C-PBR28 PET/MRI. Acs Chem. Neurosci. 9(11), 2563-2571. <https://doi.org/10.1021/acschemneuro.8b00072>.

Lukow, P. B., Martins, D., Veronese, M., Vernon, A. C., McGuire, P., Turkheimer, F. E., & Modinos, G., 2022. Cellular and molecular signatures of in vivo imaging measures of GABAergic neurotransmission in the human brain. Commun. Biol. 5(1), 372. <https://doi.org/10.1038/s42003-022-03268-1>.

Ma, X., Zhao, W., Luo, R., Zhou, F., Geng, Y., Xu, L., Gao, Z., Zheng, X., Becker, B., & Kendrick, K. M., 2018. Sex- and context-dependent effects of oxytocin on social sharing. Neuroimage 183, 62-72. <https://doi.org/10.1016/j.neuroimage.2018.08.004>.

Ma, Y., Liu, G., Hu, Y., & Long, W., 2022. Adult attachment style moderates the effect of oxytocin on neural responses to infant emotional faces. Int. J. Psychophysiol. 171, 38-47. <https://doi.org/10.1016/j.ijpsycho.2021.12.003>.

Ma, Y., Ran, G., Hu, N., Hu, Y., Long, W., & Chen, X., 2020. Intranasal oxytocin attenuates insula activity in response to dynamic angry faces. Biol. Psychol. 157, 107976. <https://doi.org/10.1016/j.biopsycho.2020.107976>.

Mayer, A. V., Wermter, A.-K., Stroth, S., Alter, P., Haberhausen, M., Stehr, T., Paulus, F. M., Krach, S., & Kamp-Becker, I., 2021. Randomized clinical trial shows no substantial modulation of empathy-related neural activation by intranasal oxytocin in autism. Sci. Rep. 11(1), 15056. <https://doi.org/10.1038/s41598-021-94407-x>.

Meier, I. M., Montoya, E. R., Spencer, H., Orellana, S. C., van Buuren, M., van Honk, J., & Bos, P. A., 2023. Preliminary data on oxytocin modulation of neural reactivity in women to emotional stimuli of children depending on childhood emotional neglect. Dev. Psychobiol. 65(1), e22349. <https://doi.org/10.1002/dev.22349>.

Mickey, B. J., Heffernan, J., Heisel, C., Peciña, M., Hsu, D. T., Zubieta, J.-K., & Love, T. M., 2016. Oxytocin modulates hemodynamic responses to monetary incentives in humans. Psychopharmacology Berl. 233(23), 3905-3919. <https://doi.org/10.1007/s00213-016-4423-6>.

Naganawa, M., Nabulsi, N., Henry, S., Matuskey, D., Lin, S.-F., Slieker, L., Schwarz, A. J., Kant, N., Jesudason, C., Ruley, K., Navarro, A., Gao, H., Ropchan, J., Labaree, D., Carson, R. E., & Huang, Y., 2021. First-in-human assessment of 11^c^-LSN3172176, an M1 muscarinic acetylcholine receptor PET radiotracer. J. Nucl. Med. 62(4), 553-560. <https://doi.org/10.2967/jnumed.120.246967>.

Normandin, M. D., Zheng, M.-Q., Lin, K.-S., Mason, N. S., Lin, S.-F., Ropchan, J., Labaree, D., Henry, S., Williams, W. A., & Carson, R. E., 2015. Imaging the cannabinoid cb1 receptor in humans with [11c] omar: assessment of kinetic analysis methods, test–retest reproducibility, and gender differences. J. Cereb. Blood Flow Metab. 35(8), 1313-1322. <https://doi.org/10.1038/jcbfm.2015.46>.

Oliver, L. D., Stewart, C., Coleman, K., Kryklywy, J. H., Bartha, R., Mitchell, D. G. V., & Finger, E. C., 2020. Neural effects of oxytocin and mimicry in frontotemporal dementia. Neurology 95(19), e2635-e2647. <https://doi.org/10.1212/WNL.0000000000010933>.

Pincus, D., Kose, S., Arana, A., Johnson, K., Morgan, P., Borckardt, J., Herbsman, T., Hardaway, F., George, M., Panksepp, J., & Nahas, Z., 2010. Inverse effects of oxytocin on attributing mental activity to others in depressed and healthy subjects: a double-blind placebo-controlled fMRI study. Front. Psychiatry 1(134). <https://doi.org/10.3389/fpsyt.2010.00134>.

Plessow, F., Marengi, D. A., Perry, S. K., Felicione, J. M., Franklin, R., Holmes, T. M., Holsen, L. M., Makris, N., Deckersbach, T., & Lawson, E. A., 2018. Effects of intranasal oxytocin on the Blood Oxygenation Level-Dependent signal in food motivation and cognitive control pathways in overweight and obese men. Neuropsychopharmacology 43(3), 638-645. <https://doi.org/10.1038/npp.2017.226>.

Preckel, K., Scheele, D., Eckstein, M., Maier, W., & Hurlemann, R., 2014. The influence of oxytocin on volitional and emotional ambivalence. Soc. Cogn. Affect. Neurosci. 10(7), 987-993. <https://doi.org/10.1093/scan/nsu147>.

Quintana, D. S., Rokicki, J., van der Meer, D., Alnæs, D., Kaufmann, T., Córdova-Palomera, A., Dieset, I., Andreassen, O. A., & Westlye, L. T., 2019. Oxytocin pathway gene networks in the human brain. Nat. Commun. 10(1), 668. <https://doi.org/10.1038/s41467-019-08503-8>.

Radhakrishnan, R., Nabulsi, N., Gaiser, E., Gallezot, J.-D., Henry, S., Planeta, B., Lin, S.-f., Ropchan, J., Williams, W., Morris, E., D’Souza, D. C., Huang, Y., Carson, R. E., & Matuskey, D., 2018. Age-related change in 5-HT_6_ receptor availability in healthy male volunteers measured with 11^c^-GSK215083 PET. J. Nucl. Med. 59(9), 1445-1450. <https://doi.org/10.2967/jnumed.117.206516>.

Radke, S., Jankowiak, K., Tops, S., Abel, T., Habel, U., & Derntl, B., 2020. Neurobiobehavioral responses to virtual social rejection in females—exploring the influence of oxytocin. Soc. Cogn. Affect. Neurosci. 16(3), 326-333. <https://doi.org/10.1093/scan/nsaa168>.

Riem, M. M. E., Bakermans-Kranenburg, M. J., Pieper, S., Tops, M., Boksem, M. A. S., Vermeiren, R. R. J. M., van Ijzendoorn, M. H., & Rombouts, S. A. R. B., 2011. Oxytocin modulates amygdala, insula, and inferior frontal gyrus responses to infant crying: a randomized controlled trial. Biol. Psychiatry 70(3), 291-297. <https://doi.org/10.1016/j.biopsych.2011.02.006>.

Rilling, J. K., DeMarco, A. C., Hackett, P. D., Thompson, R., Ditzen, B., Patel, R., & Pagnoni, G., 2012. Effects of intranasal oxytocin and vasopressin on cooperative behavior and associated brain activity in men. Psychoneuroendocrinology 37(4), 447-461. <https://doi.org/10.1016/j.psyneuen.2011.07.013>.

Robinson, J. L., Laird, A. R., Glahn, D. C., Blangero, J., Sanghera, M. K., Pessoa, L., Fox, P. M., Uecker, A., Friehs, G., Young, K. A., Griffin, J. L., Lovallo, W. R., & Fox, P. T., 2012. The functional connectivity of the human caudate: an application of meta-analytic connectivity modeling with behavioral filtering. Neuroimage 60(1), 117-129. <https://doi.org/10.1016/j.neuroimage.2011.12.010>.

Sauer, C., Montag, C., Reuter, M., & Kirsch, P., 2019. Oxytocinergic modulation of brain activation to cues related to reproduction and attachment: Differences and commonalities during the perception of erotic and fearful social scenes. Int. J. Psychophysiol. 136, 87-96. <https://doi.org/10.1016/j.ijpsycho.2018.06.005>.

Sauer, C., Montag, C., Wörner, C., Kirsch, P., & Reuter, M., 2012. Effects of a common variant in the CD38 gene on social processing in an oxytocin challenge study: possible links to autism. Neuropsychopharmacology 37(6), 1474-1482. <https://doi.org/10.1038/npp.2011.333>.

Savli, M., Bauer, A., Mitterhauser, M., Ding, Y.-S., Hahn, A., Kroll, T., Neumeister, A., Haeusler, D., Ungersboeck, J., Henry, S., Isfahani, S. A., Rattay, F., Wadsak, W., Kasper, S., & Lanzenberger, R., 2012. Normative database of the serotonergic system in healthy subjects using multi-tracer PET. Neuroimage 63(1), 447-459. <https://doi.org/10.1016/j.neuroimage.2012.07.001>.

Scheele, D., Kendrick, K. M., Khouri, C., Kretzer, E., Schläpfer, T. E., Stoffel-Wagner, B., Güntürkün, O., Maier, W., & Hurlemann, R., 2014a. An oxytocin-induced facilitation of neural and emotional responses to social touch correlates inversely with autism traits. Neuropsychopharmacology 39(9), 2078-2085. <https://doi.org/10.1038/npp.2014.78>.

Scheele, D., Plota, J., Stoffel-Wagner, B., Maier, W., & Hurlemann, R., 2015. Hormonal contraceptives suppress oxytocin-induced brain reward responses to the partner’s face. Soc. Cogn. Affect. Neurosci. 11(5), 767-774. <https://doi.org/10.1093/scan/nsv157>.

Scheele, D., Striepens, N., Kendrick, K. M., Schwering, C., Noelle, J., Wille, A., Schläpfer, T. E., Maier, W., & Hurlemann, R., 2014b. Opposing effects of oxytocin on moral judgment in males and females. Hum. Brain Mapp. 35(12), 6067-6076. <https://doi.org/10.1002/hbm.22605>.

Schmidt, A., Davies, C., Paloyelis, Y., Meyer, N., De Micheli, A., Ramella-Cravaro, V., Provenzani, U., Aoki, Y., Rutigliano, G., Cappucciati, M., Oliver, D., Murguia, S., Zelaya, F., Allen, P., Shergill, S., Morrison, P., Williams, S., Taylor, D., Borgwardt, S., Yamasue, H., McGuire, P., & Fusar-Poli, P., 2020. Acute oxytocin effects in inferring others’ beliefs and social emotions in people at clinical high risk for psychosis. Transl. Psychiatry 10(1), 203. <https://doi.org/10.1038/s41398-020-00885-4>.

Shokri-Kojori, E., Naganawa, M., Ramchandani, V. A., Wong, D. F., Wang, G.-J., & Volkow, N. D., 2022. Brain opioid segments and striatal patterns of dopamine release induced by naloxone and morphine. Hum. Brain Mapp. 43(4), 1419-1430. <https://doi.org/10.1002/hbm.25733>.

Smart, K., Cox, S. M. L., Scala, S. G., Tippler, M., Jaworska, N., Boivin, M., Séguin, J. R., Benkelfat, C., & Leyton, M., 2019. Sex differences in [11c]ABP688 binding: a positron emission tomography study of mGlu5 receptors. Eur. J. Nucl. Med. Mol. Imaging 46(5), 1179-1183. <https://doi.org/10.1007/s00259-018-4252-4>.

Spengler, F. B., Schultz, J., Scheele, D., Essel, M., Maier, W., Heinrichs, M., & Hurlemann, R., 2017. Kinetics and dose dependency of intranasal oxytocin effects on amygdala reactivity. Biol. Psychiatry 82(12), 885-894. <https://doi.org/10.1016/j.biopsych.2017.04.015>.

Spetter, M. S., Feld, G. B., Thienel, M., Preissl, H., Hege, M. A., & Hallschmid, M., 2018. Oxytocin curbs calorie intake via food-specific increases in the activity of brain areas that process reward and establish cognitive control. Sci. Rep. 8(1), 2736. <https://doi.org/10.1038/s41598-018-20963-4>.

Striepens, N., Scheele, D., Kendrick, K. M., Becker, B., Schäfer, L., Schwalba, K., Reul, J., Maier, W., & Hurlemann, R., 2012. Oxytocin facilitates protective responses to aversive social stimuli in males. Proc. Natl. Acad. Sci. U.S.A. 109(44), 18144-18149. <https://doi.org/10.1073/pnas.1208852109>.

Striepens, N., Schröter, F., Stoffel-Wagner, B., Maier, W., Hurlemann, R., & Scheele, D., 2016. Oxytocin enhances cognitive control of food craving in women. Hum. Brain Mapp. 37(12), 4276-4285. <https://doi.org/10.1002/hbm.23308>.

Turtonen, O., Saarinen, A., Nummenmaa, L., Tuominen, L., Tikka, M., Armio, R.-L., Hautamäki, A., Laurikainen, H., Raitakari, O., Keltikangas-Järvinen, L., & Hietala, J., 2021. Adult attachment system links with brain mu opioid receptor availability in vivo. Biol. Psychiatry Cogn. Neurosci. Neuroimaging 6(3), 360-369. <https://doi.org/10.1016/j.bpsc.2020.10.013>.

Vijay, A., Cavallo, D., Goldberg, A., de Laat, B., Nabulsi, N., Huang, Y., Krishnan-Sarin, S., & Morris, E. D., 2018. PET imaging reveals lower kappa opioid receptor availability in alcoholics but no effect of age. Neuropsychopharmacology 43(13), 2539-2547. <https://doi.org/10.1038/s41386-018-0199-1>.

Vogt, C., Floegel, M., Kasper, J., Gispert-Sánchez, S., & Kell, C. A., 2023. Oxytocinergic modulation of speech production—a double-blind placebo-controlled fMRI study. Soc. Cogn. Affect. Neurosci. 18(1), nsad035. <https://doi.org/10.1093/scan/nsad035>.

Watanabe, T., Abe, O., Kuwabara, H., Yahata, N., Takano, Y., Iwashiro, N., Natsubori, T., Aoki, Y., Takao, H., Kawakubo, Y., Kamio, Y., Kato, N., Miyashita, Y., Kasai, K., & Yamasue, H., 2014. Mitigation of sociocommunicational deficits of autism through oxytocin-induced recovery of medial prefrontal activity: a randomized trial. JAMA Psychiatry 71(2), 166-175. <https://doi.org/10.1001/jamapsychiatry.2013.3181>.

Wey, H.-Y., Gilbert, T. M., Zürcher, N. R., She, A., Bhanot, A., Taillon, B. D., Schroeder, F. A., Wang, C., Haggarty, S. J., & Hooker, J. M., 2016. Insights into neuroepigenetics through human histone deacetylase PET imaging. Sci. Transl. Med. 8(351), 351ra106-351ra106. <https://doi.org/10.1126/scitranslmed.aaf7551>.

Wigton, R., Tracy, D. K., Verneuil, T. M., Johns, M., White, T., Michalopoulou, P. G., Averbeck, B., & Shergill, S., 2022. The importance of pro-social processing, and ameliorating dysfunction in schizophrenia: an fMRI study of oxytocin. Schizophr. Res. Cogn. 27, 100221. <https://doi.org/10.1016/j.scog.2021.100221>.

Wittfoth-Schardt, D., Gründing, J., Wittfoth, M., Lanfermann, H., Heinrichs, M., Domes, G., Buchheim, A., Gündel, H., & Waller, C., 2012. Oxytocin modulates neural reactivity to children's faces as a function of social salience. Neuropsychopharmacology 37(8), 1799-1807. <https://doi.org/10.1038/npp.2012.47>.

Xin, F., Zhou, X., Dong, D., Zhao, Z., Yang, X., Wang, Q., Gu, Y., Kendrick, K. M., Chen, A., & Becker, B., 2020. Oxytocin differentially modulates amygdala responses during top-down and bottom-up aversive anticipation. Adv. Sci. 7(16), 2001077. <https://doi.org/10.1002/advs.202001077>.

Xu, T., Chen, Z., Zhou, X., Wang, L., Zhou, F., Yao, D., Zhou, B., & Becker, B., 2024. The central renin–angiotensin system: A genetic pathway, functional decoding, and selective target engagement characterization in humans. Proc. Natl. Acad. Sci. U.S.A. 121(8), e2306936121. <https://doi.org/10.1073/pnas.2306936121>.

Xu, X., Liu, C., Zhou, X., Chen, Y., Gao, Z., Zhou, F., Kou, J., Becker, B., & Kendrick, K. M., 2019. Oxytocin facilitates self-serving rather than altruistic tendencies in competitive social interactions via orbitofrontal cortex. Int. J. Neuropsychopharmacol. 22(8), 501-512. <https://doi.org/10.1093/ijnp/pyz028>.
